# Genetic variation associated with human longevity and Alzheimer’s disease risk act through microglia and oligodendrocyte cross-talk

**DOI:** 10.1101/2023.03.30.23287975

**Authors:** Andrew C. Graham, Eftychia Bellou, Janet C. Harwood, Umran Yaman, Meral Celikag, Naciye Magusali, Naiomi Rambarack, Juan A. Botia, Carlo Sala Frigerio, John Hardy, Valentina Escott-Price, Dervis A. Salih

## Abstract

Ageing is the greatest global healthcare challenge, as it underlies age-related functional decline and is the primary risk factor for a range of common diseases, including neurodegenerative conditions such as Alzheimer’s disease (AD). However, the molecular mechanisms defining chronological age versus biological age, and how these underlie AD pathogenesis, are not well understood. The objective of this study was to integrate common human genetic variation associated with human lifespan or AD from Genome-Wide Association Studies (GWAS) with co-expression networks altered with age in the central nervous system, to gain insights into the biological processes which connect ageing with AD and lifespan. Initially, we identified common genetic variation in the human population associated with lifespan and AD by performing a gene-based association study using GWAS data. We also identified preserved co-expression networks associated with age in the brains of C57BL/6J mice from bulk and single-cell RNA-sequencing (RNA-seq) data, and in the brains of humans from bulk RNA-seq data. We then intersected the human gene-level common variation with these co-expression networks, representing the different cell types and processes of the brain. We found that genetic variation associated with AD was enriched in both microglial and oligodendrocytic bulk RNA-seq gene networks, which show increased expression with ageing in the human hippocampus, in contrast to synaptic networks which decreased with age. Further, longevity-associated genetic variation was modestly enriched in a single-cell gene network expressed by homeostatic microglia. Finally, we performed a transcriptome-wide association study (TWAS), to identify and confirm new risk genes associated with ageing that show variant-dependent changes in gene expression. In addition to validating known ageing-related genes such as *APOE* and *FOXO3*, we found that Caspase 8 (*CASP8*) and *APOC1* show genetic variation associated with longevity. We observed that variants contributing to ageing and AD balance different aspects of microglial function suggesting that ageing-related processes affect multiple cell types in the brain. Specifically, changes in homeostatic microglia are associated with lifespan, and allele-dependent expression changes in age-related genes control microglial activation and myelination influencing the risk of developing AD. We identified putative molecular drivers of these genetic networks, as well as module genes whose expression in relevant human tissues are significantly associated with AD-risk or longevity, and may drive “inflammageing.” Our study also shows allele-dependent expression changes with ageing for genes classically involved in neurodegeneration, including *MAPT* and *HTT*, and demonstrates that *PSEN1* is a prominent member/hub of an age-dependent expression network. In conclusion, this work provides new insights into cellular processes associated with ageing in the brain, and how these may contribute to the resilience of the brain against ageing or AD-risk. Our findings have important implications for developing markers indicating the physiological age and pre-pathological state of the brain, and provide new targets for therapeutic intervention.

## Introduction

Biological ageing is the decline of adult tissue function following development. The mammalian central nervous system (CNS), shows various impairments in function with age, such as impaired spatial memory, episodic memory, and executive function^1–3^. This decline in function has been linked to cellular changes in different brain regions, including modification of synaptic connections, and inflammatory changes^3, 4^. Ageing is considered the primary risk factor for a number of neurodegenerative conditions including Alzheimer’s disease (AD), however the molecular mechanisms linking ageing and AD are not well understood^1, 5, 6^. Therefore, understanding the mechanisms of biological ageing may not only facilitate combatting the widespread and growing morbidity caused by age-related CNS decline, but also provide insights into the onset and progression of age-dependent diseases, such as AD.

Model organisms have revealed important insights into genetic pathways which can dramatically alter organismal lifespan and healthspan when manipulated. Manipulating these pathways in humans is not trivial, but understanding the causes of ageing in specific tissues, such as the CNS, may enable more directed treatments with less side-effects^3, 5, 7–9^. Gene expression changes with ageing have been studied comprehensively in different tissues, including the brain. While these studies shed light on age-associated transcriptional changes, it is not clear which of the many altered pathways drive age-associated changes, and thus which pathways should be targeted by anti-ageing therapies that may also be capable of slowing age-dependent CNS-disorders. The use of genome-wide association studies (GWAS) has advanced our ability to identify genetic variation associated with human ageing and age-related diseases, with the identification of genes such as *APOE* and *FOXO3* which are associated with ageing in some populations^10, 11^. Genetic variation in these genes may underlie some of the differences in ageing between people that are long-lived versus those that live an average lifespan. However, these genes are often multi-functional, so how genetic variation modulates biological pathways of ageing are not clear.

Ageing affects all cell types in the brain from neurons to microglia and the supporting vasculature. Emerging data are also revealing prominent disruption of myelin occurs with ageing. Myelin is a layered lipid-rich membrane produced by oligodendrocytes which ensheaths neuronal axons, providing metabolic support and enabling fast propagation of action potentials. In mammals, significant age-related changes are observed in highly myelinated areas of the brain, known as white matter, starting from around 50 years of age in humans. These include hyperintensities seen by magnetic resonance imaging (MRI), as well as myelin outfolding and accumulation of multilamellar fragments seen by electron microscopy^12–14^. Myelin regeneration or remyelination occurs during ageing and injury. Indeed, axonal function can be restored via the proliferation and differentiation of oligodendrocyte precursor cells into mature oligodendrocytes that lay down new myelin at sites of lesions^14, 15^. However, remyelination becomes impaired in the aged brain. Microglia are thought to play an important protective role in the remyelination process, by clearing degenerated myelin that accumulates during ageing and disease through phagocytosis, and by secretion of growth factors and cytokines to support oligodendrocyte maturation, and to remodel the extracellular matrix^13, 14, 16, 17^. Recent work has shown that myelin disruption with ageing in rodents can drive accumulation of amyloid plaques, by entangling microglia, potentially contributing to AD pathogenesis^18^. However, little is known about the genetic and molecular mechanisms leading to disruption of myelin with ageing, or its importance in determining healthspan of the brain and AD risk.

To better understand the age-dependent processes within the CNS that contribute to lifespan and AD pathogenesis, we used a four-step approach to identify biological pathways that are dysregulated with age, and are enriched for genes associated with longevity or AD risk. Firstly, we identified common human genetic variation associated with ageing or AD at the gene-based level by aggregating individual SNPs’ significance and by testing the joint association of all SNPs in the gene accounting for linkage disequilibrium (LD) and number of SNPs per gene. Secondly, we overlaid this human genetic variation onto age-associated gene co-expression networks, generated from bulk and single-cell RNA-sequencing (scRNA-seq)-based transcriptomic profiling of the mouse hippocampus, a region which is thought to be central to both age-related cognitive decline and AD^3, 19^. Thirdly, we tested for an enrichment of gene variants associated with ageing and AD in different co-expression networks which are dysregulated with age in the mouse and human hippocampus. We found a significant enrichment of gene variants associated with AD in human microglial and oligodendrocytic networks, whose expression was also significantly increased with age. Assessment of changes in co-expression modules with ageing at the single-cell level in microglia identified stark similarities to changes in young mice treated with cuprizone to induce demyelination, indicating that age-related demyelination may provide a link between these two AD-associated processes. Furthermore, we saw a modest but significant enrichment of genes associated with ageing in humans in a single-cell co-expression network, highly expressed by homeostatic microglia, which is lost during ageing and demyelination. Finally, we performed a transcriptome-wide association study (TWAS), to identify and confirm new risk genes associated with longevity, and compared the results to the genes associated with AD in the recently published AD TWAS^20^. Our data suggest a hypothesis whereby potentially linked microglial and oligodendrocytic responses to ageing may modify AD risk, while the degree of age-dependent loss of homeostatic microglial functions may influence longevity. These findings may allow earlier tracking of disease progression, and provide new targets for AD prevention, as well as facilitation of healthy CNS ageing.

## Results

### Identification of genes harbouring common genetic variation associated with longevity and AD

We initially identified genes harbouring common genetic variants associated with lifespan to capture each gene’s collective SNPs, even if a single SNP does not reach genome-wide significance, by performing a gene-based analysis with the summary statistics from a meta-analysis combining three different GWAS associated with lifespan^10^. Our gene-based analysis showed *APOC1* (P_FDR_ = 2.49e-63), and *APOE* (P_FDR_ = 1.95e-46), were the strongest genes associated with the measures of lifespan (top genes associated with lifespan are given in Supplementary Table 1, P_FDR_ < 0.01). The results of a similar gene-based analysis on the data from the International Genomics of Alzheimer’s Projects (IGAP)^21^ to identify genes harbouring common genetic variation associated with AD risk are given in Supplementary Table 2 (top genes associated with AD, P_FDR_ < 0.01).

### Transcriptional networks of oligodendrocytic and microglial genes are associated with age in the mouse hippocampus

We then sought to gain insights into which biological pathways contained the genes associated with longevity and AD (Supplementary Tables 1 and 2), while determining how these pathways are affected by ageing. To determine transcriptomic changes associated with ageing at the systems level, we performed weighted gene co-expression analysis (WGCNA)^22^, with an optimization for constructing more biologically meaningful co-expression networks^23^, to group genes which were co-expressed in the hippocampi of C57BL/6J wild-type mice at 2-18 months-of-age^24^. We identified eight co-expression modules whose expression was significantly associated with age (Pearson’s product-moment correlation with age, here and thereafter, R^2^ > 0.4, p < 0.01; Fig. 1a), from more than 20 modules. Using cell-type enrichment analysis, we determined that the module with strongest correlation to age (R^2^ = 0.93, p < 0.001), was heavily enriched for microglial genes (microglial module). A second module which significantly correlated with age (R^2^ = 0.54, p < 0.01), was heavily enriched for oligodendrocytic genes (mouse oligodendrocytic module) (Fig. 1b). Corresponding with their cell-type annotations, the mouse microglial module was significantly enriched for biological annotations associated with innate immunity, while the oligodendrocytic module was enriched for myelination and axon ensheathment (Fig. 1c, d).

**Figure 1.**
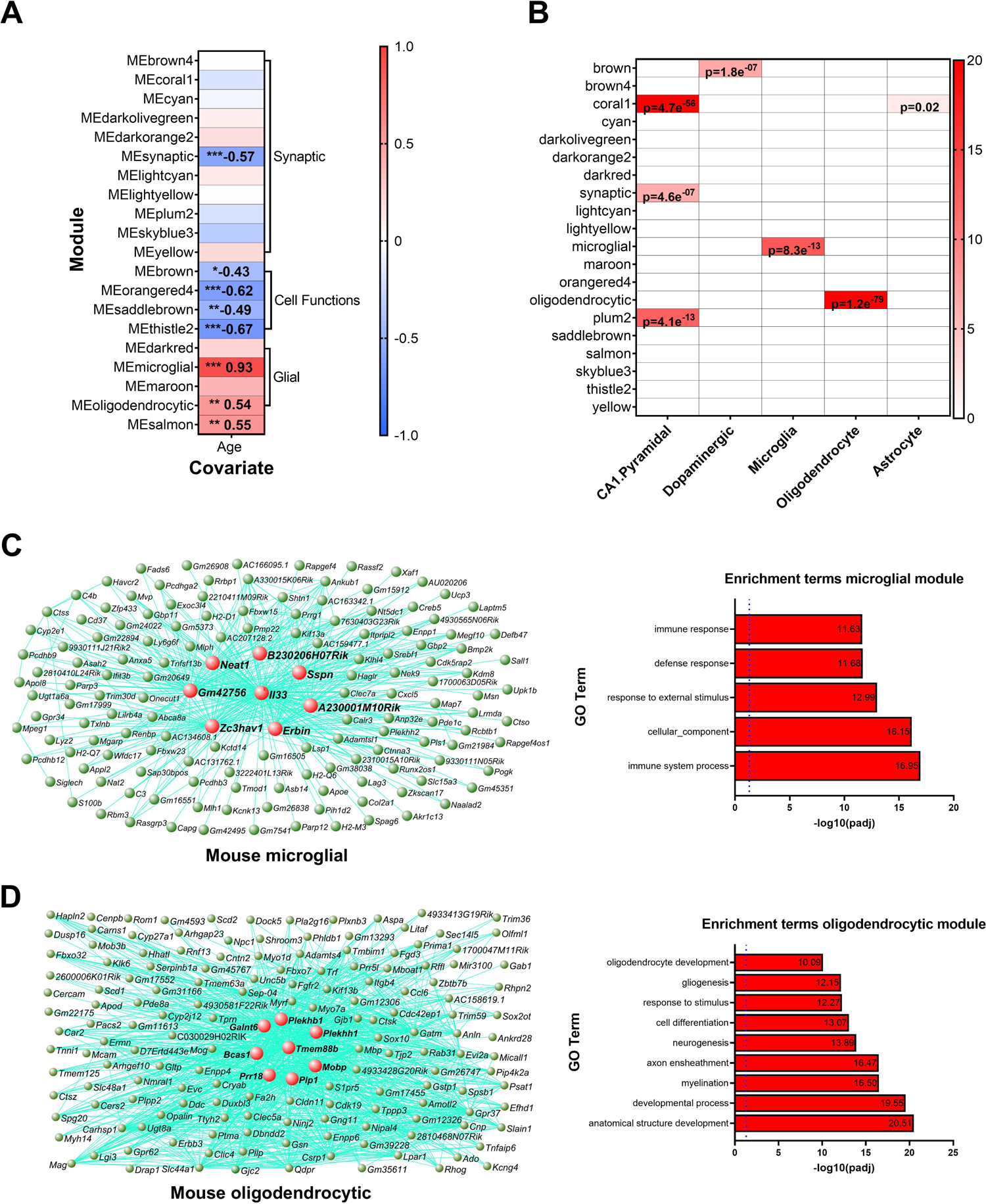
Gene co-expression modules enriched for microglial and oligodendrocytic genes are significantly associated with age in the mouse hippocampus. A) Co-expression analysis of bulk RNA-seq of the mouse hippocampus produced eight gene modules with significant correlation to age using our Mouseac data^24^. Pearson’s product-moment correlation with age, value given in each cell where R^2^>0.4, ** p <0.01, *** p < 0.001. No module has a significant association with either sequencing batch or lane, validating our batch correction strategy. B) Cell-type enrichment analysis reveals a significant enrichment of microglial genes in the microglial module, oligodendrocyte genes in the oligodendrocytic module, and dopaminergic neuron genes in the brown module. * p<0.05, ** p <0.01, *** p < 0.001. C) Network plot of the 151 most connected genes in the microglial module (left). Hub genes are shown in red. Biological annotations (right). D) Network plot of the 152 most connected genes in the oligodendrocytic module (left). Hub genes are shown in red. Biological annotations (right). Full networks given in Supplementary Table 10 and 11.

Genes with the highest connectivity to other genes within a co-expression module are known as hub genes, and are postulated to drive the response of the entire network^25, 26^. The mouse microglial module contained as hub genes, *Il33, Erbin, Sspn, Neat1,* and *Zc3hav1,* and also contained the canonical microglial genes *Trem2* and *Apoe* (Fig. 1c). The oligodendrocytic module included the hub genes: *Mobp*, *Tmem88b*, *Plp1*, *Plekhh1*, *Plekhb1*, *Galnt6*, *Bcas1* and *Prr18* (Fig. 1d). Interestingly, while analysis of single-cell RNA-seq data determined that 14 of the 15 most-connected genes in the bulk microglial module were also detected by scRNA-seq of the mouse hippocampus at 3 and 24 months-of-age^27^, 8 of these genes were predominantly detected in oligodendrocytes, where they all showed elevated expression with age (Supplementary Fig. 1). Overall, these scRNA-seq findings indicate that while the microglial module is significantly enriched for microglial genes such as *Apoe* and *Trem2*, its hub genes are contributed by other cell types, disproportionately oligodendrocytes, including genes coding for signalling molecules known to affect microglial function such as the interleukin *Il33* and complement component *C4b*^28–30^. This suggests age-related signalling from oligodendrocytes may trigger microglial activation in the aged hippocampus.

Since transcriptional changes in mice can be affected by many co-variates, such as mouse strain, husbandry conditions, diet, and technical variation due to RNA preparation, RNA-seq protocols and analysis pipeline, we then tested whether the age-dependent co-expression networks were preserved in other bulk RNA-seq datasets of wild-type mouse hippocampus or cortex at various ages from 13-126-weeks of age from three other labs, processed via a variety of pipelines^31–33^. We saw strong preservation of both the microglial and oligodendrocytic modules, with respect to gene expression connectivity and correlation to age in the different datasets (strong preservation, z.summary > 10 for co-expression patterns between module genes; microglial module R^2^ with age >0.93 and oligodendrocytic module R^2^ > 0.58 against two further query datasets; Supplementary Fig. 2). This provides evidence that the age-dependent co-expression networks are preserved between different mice, labs, and sequencing experiments.

### Single-cell RNA-seq changes of age-dependent gene expression

Given that recent scRNA-seq studies have revealed that microglia respond heterogeneously to various challenges, we sought to determine age-dependent co-expression networks at the single-cell level, using scRNA-seq data generated from microglia isolated from male and female wild-type mice hippocampi at 3, 6, 12, and 21 months-of-age (2 mice of each sex/age, pooled prior to sequencing) (data from Sala Frigerio *et al.*, 2019^34^). Previous analysis of this data identified 6 microglial subpopulations: homeostatic microglia 1 and 2 (HM1 and HM2), which highly express genes associated with microglial homeostatic functions; cycling proliferating microglia (CPM), which express genes involved in the cell cycle; activated response microglia (ARM); interferon response microglia (IRM); and transiting microglia (TRM) which may be an intermediate state between homeostatic microglia and ARM^34^. We identified eight co-expression modules expressed by distinct microglial sub-populations in the mouse hippocampus (Fig. 2a). To reliably assess the age-related changes in expression of these modules in more replicates than those available in scRNA-seq data, we assessed these modules’ dysregulation (using the mean expression of the core 100 genes with the highest connectivity) between independent replicates of microglia isolated from wild-type mice hippocampi at 2 and 16-18 months-of-age, and profiled by bulk RNA-seq (data from O’Neil *et al.*, 2018^35^).

**Figure 2.**
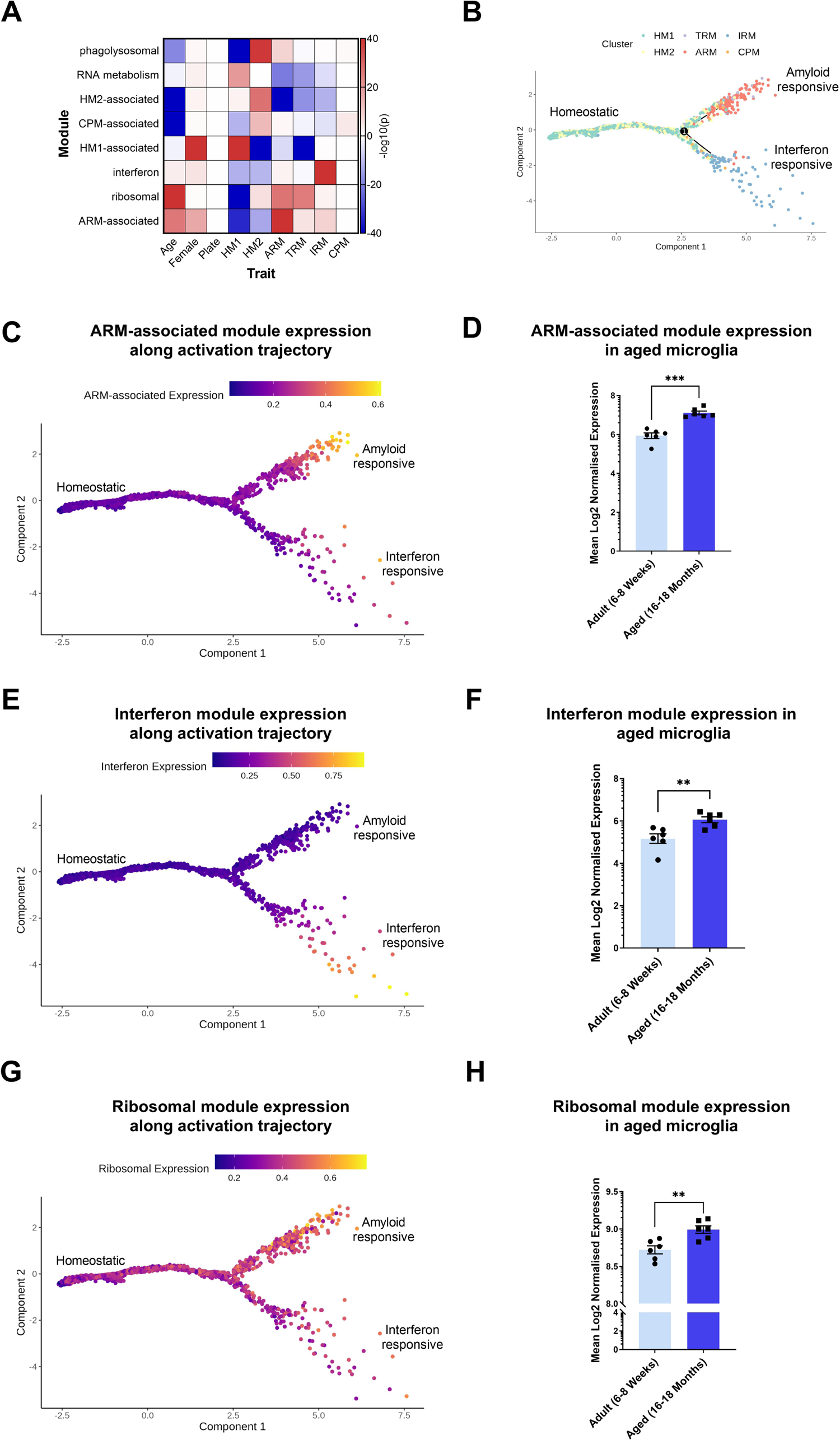
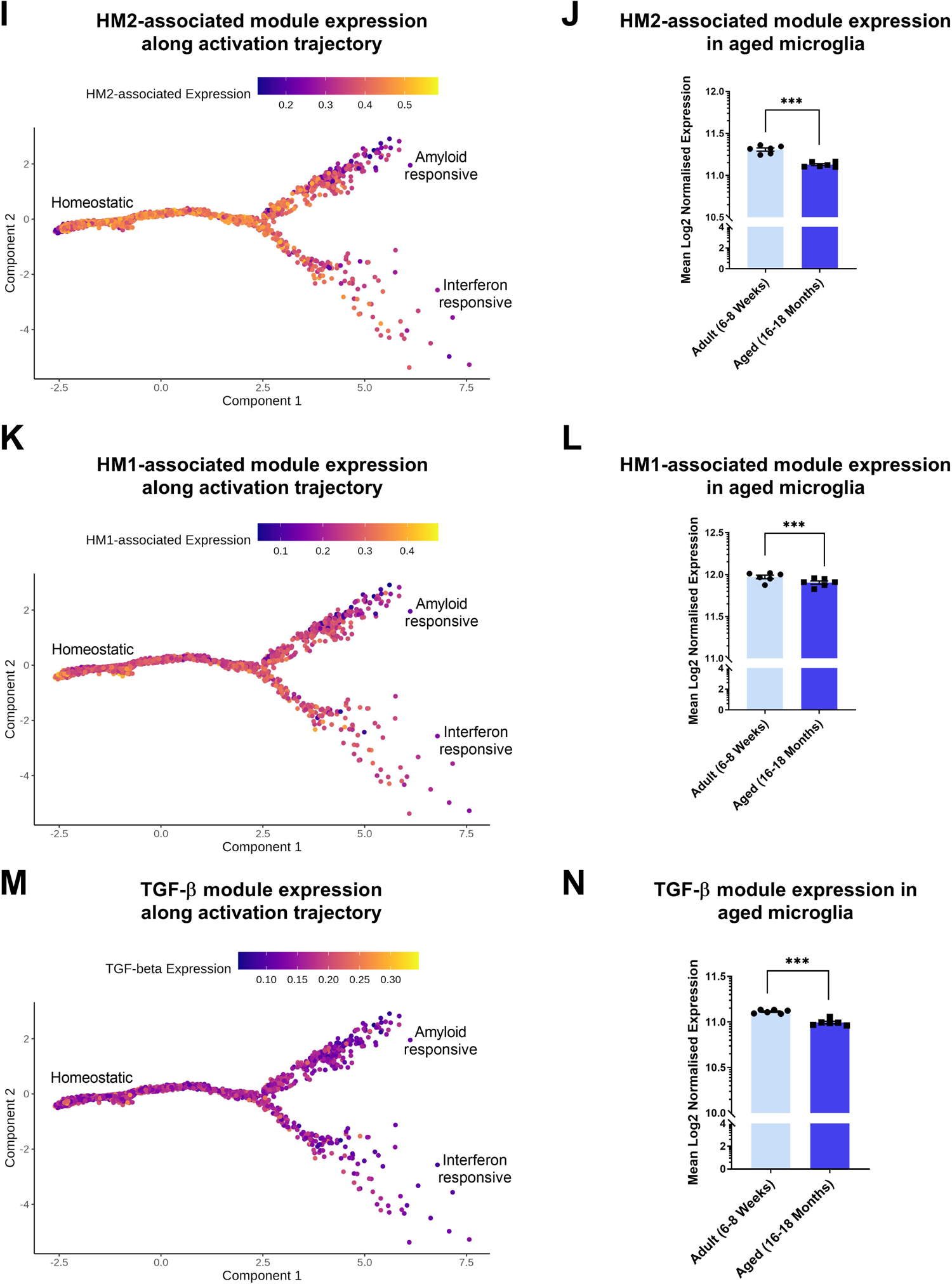
Five gene co-expression modules expressed by distinct microglial subpopulations are dysregulated with age in the mouse hippocampus. A) Association, p(-log10), between the expression of co-expression modules formed by co-expression analysis of data generated by scRNA-seq of microglia (Mg) isolated from wild-type mice at 3, 6, 12, and 21 months-of-age, and female sex, sequencing plate, and six microglial subpopulations identified by the original authors^34^. Subpopulations: HM1 – Homeostatic Microglia 1, HM2 – Homeostatic Microglia 2, TRM –Transiting Microglia, ARM – Activated Response Microglia, IRM – Interferon Response Microglia. Negative p(-log10) indicates a negative association. B) Pseudotime analysis with Monocle 2 identified a bifurcating trajectory, with HM1 as a root state and ARM and IRM as two terminal states. C,E,G,I,K,M). Expression of the differentially expressed genes in the ARM-associated (C), interferon (E), Ribosomal (G), HM2-associated (I), HM1-associated (K), and TGF-β (M) co-expression modules shown along this Pseudotime trajectory. D,F,H,J,L,N) Comparison of expression of the 100 most central genes (as a proxy for module expression) from the ARM-associated (D), interferon (F), ribosomal (H), HM2-associated (J), HM1-associated (L), and TGF-β (N) modules in microglia isolated from the mouse hippocampus at 6-8 and 16-18 months-of-age and profiled by RNA-seq^35^.

Six gene modules exhibited altered expression with age. A module of genes representing activated microglia exhibited strikingly increased expression with age (ARM-associated module, p = 1.2e-3, Student’s *t*-test; Fig. 2a, b, c, d). This module was predominantly expressed by the ARM subpopulation of activated microglia, which are synonymous with the disease-associated microglia (DAM) subpopulations seen in AD mouse models^34, 36^. Interestingly, this module’s hub genes (*Spp1, Clec7a, Lilrb4a*; Fig. 3a) have been reported to mark microglia scavenging fragmented myelin in the corpus callosum, which also become more numerous with ageing^13^. This module is significantly enriched for biological annotations related to phagocytosis, cytokine production, and TYROBP signalling (Supplementary Fig. 3a). Another gene module consisting of interferon-stimulated genes was also increased with ageing (Interferon module, p = 8.0e-3, Student’s *t*-test; Fig. 2f, and Fig. 3b), and was associated with a distinct subpopulation of interferon responsive microglia, IRM, so was named the interferon module. This module is significantly enriched for biological annotations related to the interferon response (Supplementary Fig. 3b). A further module containing ribosomal genes, was more modestly increased with ageing (ribosomal module; p = 3.0e-2, Student’s *t*-test, Fig. 2h and Supplementary Fig. 3). In contrast, a module strongly associated with a homeostatic subcluster, HM2, was decreased during ageing (HM2-associated module, p = 1.4e-5, Student’s *t*-test; Fig. 2j and Fig. 3c). While a module associated with a different homeostatic subpopulation of microglia, HM1 (HM1-associated module, p = 8.7e-3, Student’s *t*-test; Fig. 2l), and a module associated with TGF-β signalling (TGF-β module, p = 4.0e-4, Student’s *t*-test; Fig. 2n and Supplementary Fig. 3), were more modestly decreased during ageing. Finally, modules containing phagocytosis and lysosomal genes (phagolysosomal module; Fig. 3d and Supplementary Fig. 3), or associated with the CPM cluster (CPM-associated module), were not significantly altered during ageing (p > 0.05, Student’s *t*-test).

**Figure 3.**
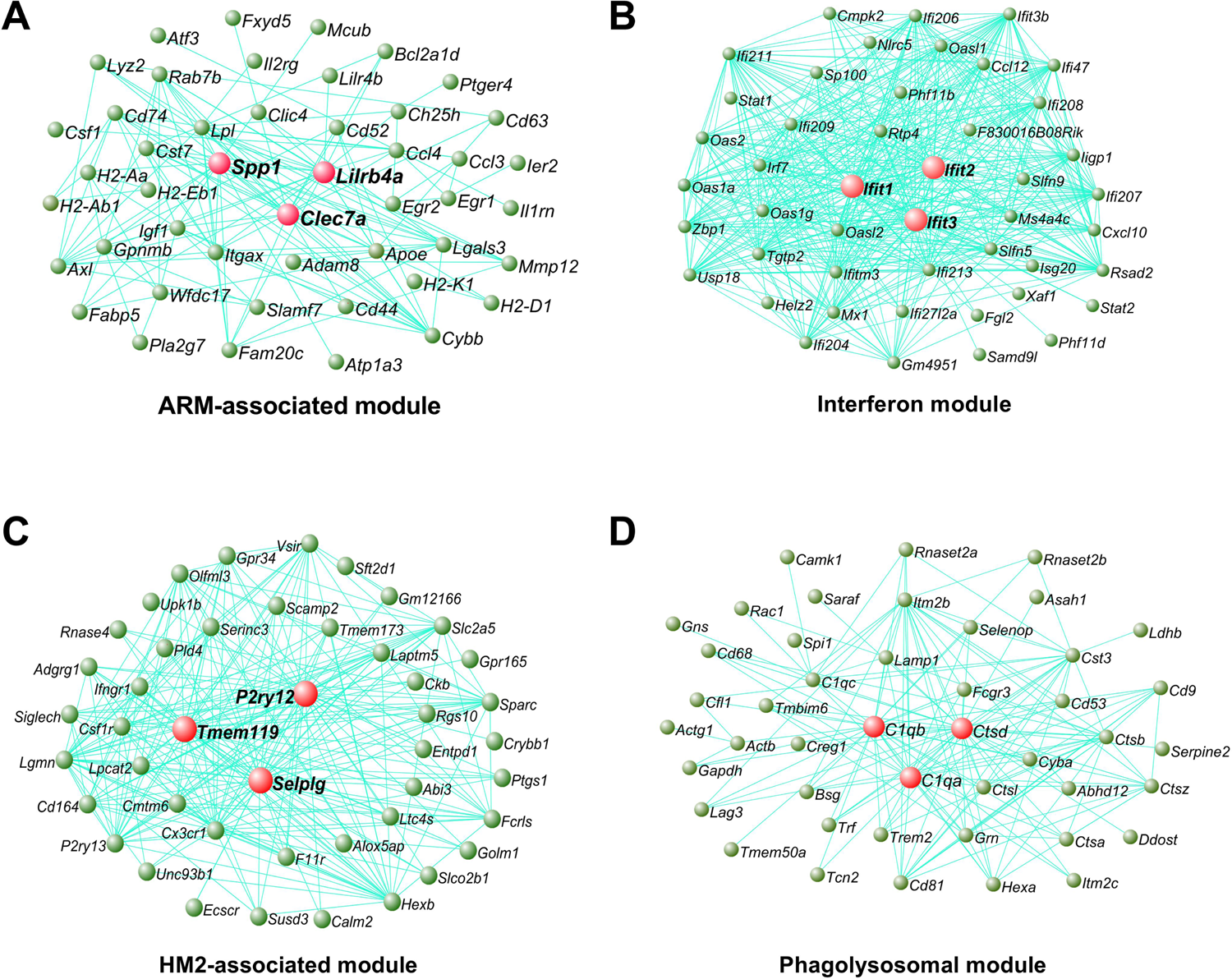
Gene co-expression networks from microglia analysed by scRNA-seq associated with ageing. The genetic networks from microglial cells isolated from wild-type mice at 3, 6, 12 and 21 months-of-age analysed by scRNA-seq^34^. Network plot of the 45 most connected genes in the ARM-associated (A), interferon (B), HM2-associated (C), and phagolysosomal (D), modules. Green nodes represent genes, edge lines represent co-expression connections, and the central large red nodes are the hub genes. Full networks given in Supplementary Table 12, 13, 14 and 15.

To gain further insights into how these modules relate to microglial activation, we used semi-supervised Pseudotime analysis to order cells by their expression of genes which nominally significantly differ (p < 0.05) between subpopulations (Fig. 2b), using Monocle 2^37, 38^. We saw that the HM1 microglial subpopulation could be considered as the root state, and then the ARM and IRM populations acted as the endpoint of distinct activation trajectories, with HM2 and TRM as intermediate states. We saw that the ARM trajectory was defined by increased expression of the ARM-associated module of genes, although some cells in the IRM trajectory also expressed this gene module (Fig. 2c). The ribosomal module was also upregulated along the ARM trajectory (Fig. 2g). While the IRM trajectory was defined by increased expression of the interferon module (Fig. 2e). In contrast, expression of the HM1-/HM2-associated, and TGF-β-associated modules was highest in the root homeostatic state and decreased along both activation trajectories (Fig. 2i, k, m). Collectively these data show that microglial transcription programs associated with states considered to be activated are increased with age in the mouse hippocampus, and transcriptional signatures associated with homeostatic functions are decreased with age.

### Single-cell RNA-seq signatures of microglia associated with ageing resemble the changes seen with demyelination

Due to the enrichment of oligodendrocytic hub genes in our bulk microglial module, we then sought to gain further insight into whether the microglial responses during ageing seen in our single-cell analysis may be influenced by interactions with oligodendrocytes. Demyelination is a major feature of oligodendrocyte ageing in the mouse hippocampus^39^. Therefore, we examined how these age-dependent scRNA-seq modules were affected when microglia were exposed to demyelination in young mice, following feeding with cuprizone (data from Ref^40^). Interestingly, we found that the co-expression of genes within our age-associated microglial modules (Fig. 2 and 3) were strongly preserved in young mice fed a demyelinating diet (Fig. 4a), indicating that these genes were also co-expressed in these mice exhibiting demyelination. Looking again at our age-dependent modules of interest (Fig. 2 and 3), we found that the average expression of their 100 most connected genes showed the same change with demyelination as they showed with ageing. The ARM-associated module (ARM population) was increased after both 5-weeks (p = 6.7e-5, Dunnett’s test), and 12-weeks (p = 9.3e-6, Dunnett’s test) of a demyelinating cuprizone diet, mirroring the changes seen with ageing (Fig. 4b). The average expression of core interferon-response module genes was increased following 5-weeks (p = 4.0e-3, Dunnett’s test) of a demyelinating cuprizone diet, as we saw during ageing, although this subsided after 12-weeks of the cuprizone diet (p = 2.4e-1, Dunnett’s test; Fig. 4c). While the ribosomal module was more modestly elevated following 5- and 12-weeks on the cuprizone diet (5-weeks: p = 2.9e-3; 12-weeks: p = 2.6e-2; Dunnett’s tests; Fig. 4d). In contrast, the average expression of core genes in the HM2-associated, HM1-associated, and TGF-β modules were decreased after both 5-weeks (HM2, p = 1.3e-2; HM1, p = 3.4e-2; TGF-β, p = 1.2e-3; Dunnett’s tests) and 12-weeks (HM2, p = 2.9e-4; HM1, p = 7.8e-3; TGF-β, p = 6.0e-4; Dunnett’s tests) (Fig. 4e, f, g) of a demyelinating cuprizone diet, mirroring the repression of these modules during ageing (Fig. 2). Overall, these data provide evidence that the transcriptional programmes we identified were similarly changed in microglia during ageing and in response to the breakdown of myelin in young mice.

**Figure 4.**
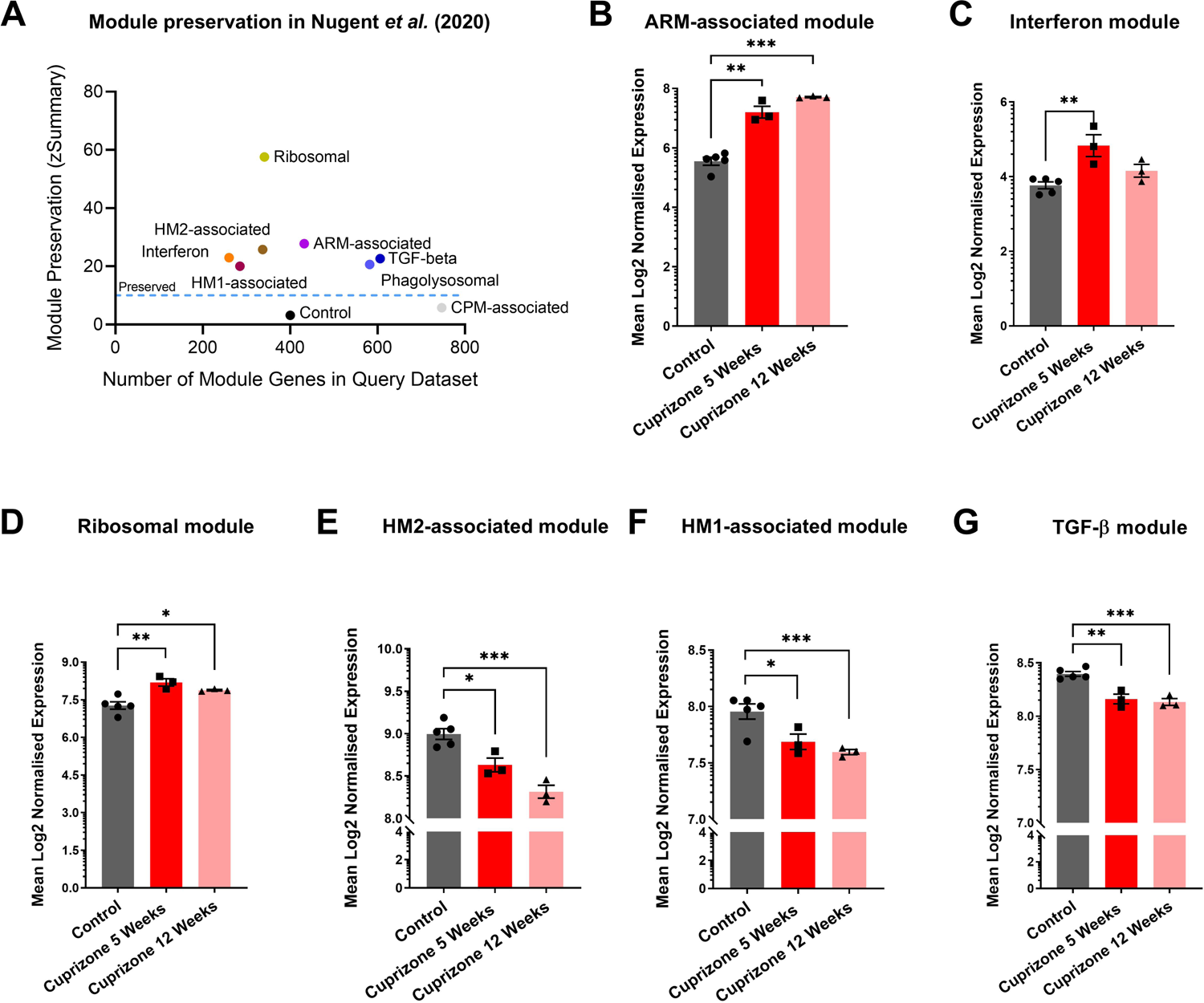
Gene co-expression network changes in microglia isolated from mice treated with cuprizone and analysed using scRNA-seq. A) Preservation analysis of age-related modules in a dataset generated by scRNA-seq of microglia isolated from the corpus callosum of 9-11 month old WT mice fed a control or demyelinating cuprizone diet for 5- or 12-weeks^40^. Control module represents results for randomly chosen genes of a similar module size. B-F) Comparison of expression of the 100 most central genes (as a proxy of module expression) from the ARM-associated (B), interferon (C), ribosomal (D), HM2-associated (E), HM1-associated (F), and TGF-β (G) modules in microglia isolated from the corpus callosum fed a control diet or a demyelinating cuprizone diet for 5- or 12-weeks and profiled by bulk RNA-seq^40^.

### Transcriptional networks of oligodendrocytic and microglial genes are associated with age in the human hippocampus

As ageing in mice may not reflect ageing in humans^41, 42^, we then performed a comparable analysis of bulk RNA-seq data from the non-diseased hippocampi of 196 cognitively normal individuals from the GTEx consortium^43^. This analysis produced several co-expression modules significantly correlated to age (Fig. 5a). Strikingly, the module most correlated to age was enriched for microglial genes (R^2^ = 0.73, p < 0.001; Hu_Microglial module), as was the case in mice, with genes such as *TYROBP*, *SYK* and *CD53* presenting as hub genes (Fig. 5a, b). Additionally, expression of an oligodendrocytic gene enriched module (R^2^ = 0.58, p < 0.001; Hu_Oligodendrocyte module) was also significantly positively associated with age, with genes such as *ENPP2*, *CNTN2*, *PSEN1*, *SLC44A1* and *CYP27A1* presenting as hub genes (Fig. 5a, c). However, despite this broad inter-species similarity, there were differences in the gene constituents of these modules compared to their mouse counterparts, as the human and mouse microglial, and oligodendrocytic co-expression modules showed only moderate preservation of their gene co-expression patterns in the mouse data (Fig. 5d). Correspondingly, 42 genes in the human microglial module have orthologues in the mouse microglial module (p = 3.4e-26 overlap, Fisher’s Exact Test), and 90 genes in the human oligodendrocyte module have orthologues in the mouse oligodendrocyte module (p = 1.6e-75 overlap, Fisher’s Exact Test). Genes unique to the mouse age-dependent microglial network included *B2m*, *Cst7*, *Pik3cg* and *Ccl4*, and also *Il33* which is expressed mostly by oligodendrocytes and is part of the age-dependent module because it shows expression changes very similar to the microglial genes. In contrast, genes unique to the human age-dependent microglial network included *ABI3*, *ALOX5AP*, *C1QA-C*, *CD68*, *CD74*, *CSF1R*, *FCER1G*, *ITGAM*, *MRC1*, *MS4A6A*, *P2RY12*, *RUNX1*, *SAMSN1*, *SPI1*, *STAT6*, *SYK* and *TYROBP*. To gain further insights into how the age-dependent microglial response is different between humans and mice, we investigated biological annotations associated with the age-dependent genes unique to either mouse or human using the Gprofiler2 R package^44^. The age-dependent genes unique to mouse were associated with antigen presentation involving the major histocompatibility complex (MHC)(Supplementary Fig. 4a). Whereas age-dependent genes unique to humans were associated with leukocyte activation, cytokine production and T cell activation (Supplementary Fig. 4b). Therefore, the human and mouse age-related gene networks are overlapping, but each organism has its own distinct components of the network and so some of the human age-related processes which modulate AD risk and longevity in humans are not seen in mice.

**Figure 5.**
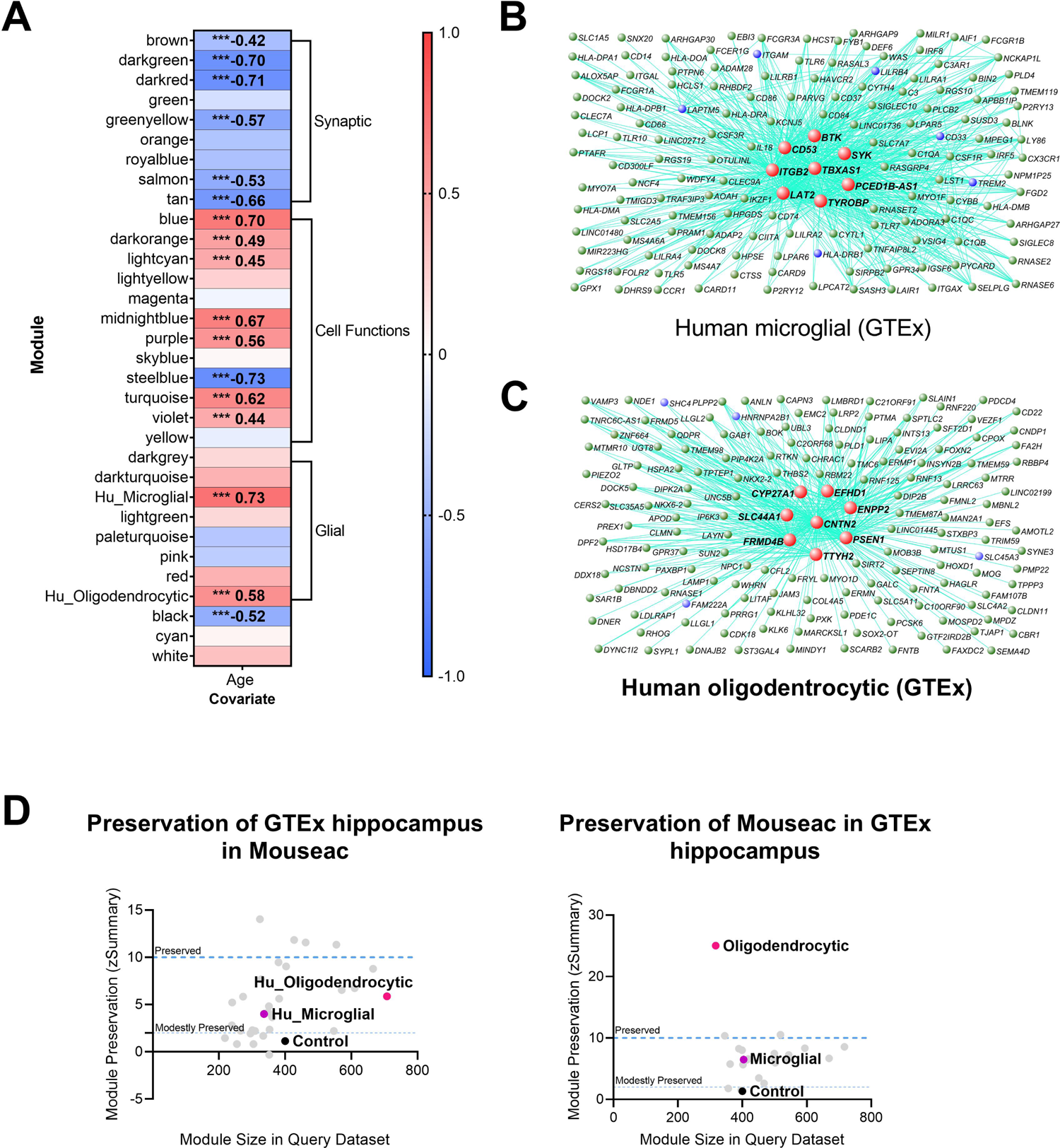
Co-expression analysis of the human hippocampus reveals aspects of ageing preserved in mice and aspects unique to humans. A) Co-expression analysis produced seventeen gene modules whose expression significantly correlates to age in the human hippocampus (correlation>0.4), from the GTEx data^43, 49^. *** p<0.001. B) Network diagram of the most central 152 genes from the human microglial network. C) Network diagram of the most central 150 genes from the human oligodendrocytic network. D) Mouse co-expression modules are generally moderately preserved in humans. Therefore, it appears only subsections of the mouse gene networks are preserved in humans. Human GTEx co-expression module preservation in mouse data (left), and mouse co-expression module preservation in human GTEx RNA-seq data (right). z.summary is an amalgamation of other preservation statistics (z.density, mean connection strength per gene, and z.connectivity, sum of all connections), found to depict module preservation better than these statistics alone or simple gene overlap measures^110^. Human data was compared to our Mouseac data^24^. Full networks given in Supplementary Table 16 and 17.

### Microglial and oligodendrocytic gene networks are enriched for AD risk genes

Our previous study testing for enrichment of common human gene variants in a co-expression network associated with amyloid plaques demonstrated that genetic risk for AD is substantially encoded by genes expressed during the response of microglia to these plaques^24^. Therefore, to investigate whether our age-dependent co-expression networks were connected to AD, we tested the mouse and human age-dependent bulk RNA-seq co-expression networks for enrichment of genes, or mouse orthologues of human genes, significantly associated with AD (top genes associated with AD are given in Supplementary Table 2, P_FDR_ < 0.01). Enrichment analyses were performed by comparing the number of overlapping genes in our gene-sets with randomly sampled gene-sets matched for LD, gene size and the number of genes in a set. We saw no significant enrichment of common human AD-associated genes in the age-dependent mouse networks from bulk RNA-seq (Fig. 1 and Table 1). In contrast, we saw a significant enrichment of overlapping genes associated with AD in both the age-dependent human microglial network (enrichment p = 1e-05, bootstrap-based test) and oligodendrocytic network (enrichment p = 0.033, bootstrap-based test) (Fig. 5b, c, and Table 1). The genes driving the enrichment (gene-based p-values < 0.01) of the microglial module are known AD-associated risk genes (*CD33, GAL3ST4, HLA-DRB1, INPP5D, MS4A4A, SPI1, TREM2*), and putative risk genes (*ITGAM, LAPTM5* and *LILRB4*)^24^ (Fig. 5 and Supplementary Table 3). We also observed genes with gene-based p-values < 0.01 not before linked to AD, which are likely to contribute to AD risk when combined with their co-expression with other genes enriched for AD risk, namely, *APOC2, ARHGAP45, ATP8B4, CMTM7, COX7A1, DOCK3, MARCO, MLPH*, *NOP2, PCED1B* and *TMC8* (Table 1; Supplementary Table 3). Therefore, the age-dependent microglial genetic network identified here both highlights a role for microglial-associated age-related changes in determining AD risk, and predicts new putative risk genes for AD.

**Table 1.** Test of enrichment of age-dependent gene modules with genes variants associated with Alzheimer’s disease from GWAS of Kunkle *et al.* (2019)^21^. Gene modules from mouse hippocampus bulk RNA-seq (Fig. 1), mouse hippocampus scRNA-seq (Fig. 2, 3), and human hippocampus bulk RNA-seq (Fig. 5). ARM, activated response microglia. HM2, homeostatic microglial subcluster 2.

| MAGMA analysis |  |  |  | Enrichment compared to random gene set by bootstrapping for genes p<0.01 association |  |
| --- | --- | --- | --- | --- | --- |
| Module | Genes | Competitive p value | Self P value | Genes | P value |
| Human bulk Microglial | 434 | 0.023 | 3.11e-6 | 26 | 1e-5 |
| Human bulk Oligodendrocytic | 780 | 0.406 | 0.018 | 29 | 0.033 |
| Mouse bulk Microglial | 240 | 0.064 | 0.002 | 9 | 0.160 |
| Mouse bulk Ribosomal | 333 | 0.429 | 0.007 | 9 | 0.515 |
| Mouse bulk Mitochondrial | 396 | 0.711 | 0.108 | 5 | 0.975 |
| Mouse bulk Oligodendrocytic | 236 | 0.989 | 0.836 | 3 | 0.943 |
| Mouse single-cell Interferon | 308 | 0.045 | 0.003 | 10 | 0.28 |
| Mouse single-cell ARM | 499 | 0.503 | 0.001 | 17 | 0.067 |
| Mouse single-cell Phagolysosomal | 1337 | 0.608 | 0.002 | 45 | 0.054 |
| Mouse single-cell HM2 Homeostatic | 964 | 0.533 | 0.003 | 28 | 0.329 |

**Table 2.** Test of enrichment of age-dependent gene modules with genes variants associated with ageing from GWAS of Timmers *et al.* (2020)^10^. Gene modules from mouse hippocampus bulk RNA-seq (Fig. 1), mouse hippocampus scRNA-seq (Fig. 2, 3), and human hippocampus bulk RNA-seq (Fig. 5). ARM, activated response microglia. HM2, homeostatic microglial subcluster 2.

| MAGMA analysis |  |  |  | Enrichment compared to random gene set by bootstrapping for genes p<0.01 association |  |
| --- | --- | --- | --- | --- | --- |
| Module | Genes | Competitive p value | Self P value | Genes | P value |
| Human bulk Microglial | 423 | 0.982 | 0.539 | 12 | 0.806 |
| Human bulk Oligodendrocytic | 764 | 0.871 | 0.106 | 21 | 0.804 |
| Mouse bulk Microglial | 238 | 0.507 | 0.107 | 7 | 0.671 |
| Mouse bulk Ribosomal | 321 | 0.510 | 0.002 | 12 | 0.472 |
| Mouse bulk Mitochondrial | 389 | 0.163 | 3.00e-4 | 11 | 0.801 |
| Mouse bulk Oligodendrocytic | 235 | 0.776 | 0.103 | 7 | 0.638 |
| Mouse single-cell Interferon | 305 | 0.284 | 0.056 | 10 | 0.553 |
| Mouse single-cell ARM | 487 | 0.721 | 0.478 | 9 | 0.969 |
| Mouse single-cell Phagolysosomal | 1310 | 0.759 | 2.88e-5 | 49 | 0.146 |
| Mouse single-cell HM2 Homeostatic | 936 | 0.734 | 0.037 | 39 | 0.032 |

None of the genes within the human oligodendrocytic module had previously been identified as GWAS hits for AD risk. Nevertheless, *CLASRP* meets genome-wide significance in our analysis (P_FDR_ = 1.50e-10), highlighting the benefit of our gene-based association analysis, and indicating a potential oligodendrocytic role for this gene in the human hippocampus. *CLASRP* is in the same region as *APOE*, but is included here as a gene of interest because it shows age-dependent expression changes. Moreover, the large number of co-expressed genes (29 genes) containing genetic variation associated with AD below our threshold (gene-based p < 0.01; Table 1; Supplementary Table 4), indicates that genetic variation which influences myelination during ageing may play a role in AD pathogenesis.

### A homeostatic microglial gene network is enriched with common gene variants associated with longevity

To determine which specific microglial transcriptional programs are particularly associated with AD and longevity, we repeated our assessment of enrichment for common genetic variation associated with these traits, using modules from our scRNA-seq analysis (Fig. 3). Comparing the number of overlapping genes from our modules to bootstrapped modules containing the same number of genes randomly selected from those reliably detected from this dataset (to account for the relative enrichment of microglia-expressed genes with these traits), we observed a trend for enrichment of genes associated with AD within the genes present in the ARM- and phagolysosomal-associated modules (Fig. 3 and Table 1; Supplementary Tables 5 and 6). Furthermore, our analysis suggests that the HM2-associated module is significantly, but modestly, enriched for genetic variation associated with longevity (enrichment p = 0.032, bootstrap-based test) (Table 2; Supplementary Table 7). This module contains the mouse orthologue of *FOXO3* and *ATXN2*, which have previously been linked to longevity by GWAS studies, as well as cognitive decline (*FOXO3*), and late-onset Parkinson’s disease and Spinocerebellar Ataxia 2^10, 45, 46^. This homeostatic microglial module also includes the mouse orthologue of *CASP8*, which acts as a molecular switch to control cell death via apoptosis, necroptosis and pyroptosis^47^. Importantly, the longevity-associated role of these genes has not previously been linked to microglia.

### Transcriptome-wide association study highlights the new genes associated with AD risk and genes expressed by homeostatic microglia that are associated with longevity

To investigate genetic variation associated with longevity using an independent approach, we performed a TWAS, to identify genes in which a change in expression is associated with genetic contribution to longevity. Data are not available to perform this analysis with microglia, but since peripheral monocytes differentiate into microglia-like macrophages within the CNS, we used expression data from naïve (CD14) and induced monocytes (LPS and IFN-gamma) from the Fairfax dataset^48^, as previously performed for AD^20^. In addition, we used expression data from the Genotype-Tissue Expression (GTEx) Project including the brain and hippocampus^43, 49^, whole blood from the young Finns study (YFS)^50^, and the Netherlands twin register peripheral blood (NTR)^51^. These datasets include sorted CD14+ve myeloid cells, whole blood, and hippocampus, which can shed light on myeloid cell transcription. Using the AD TWAS, this approach confirmed the importance of known AD risk genes such as *APOE*, *BIN1*, *CD33*, *CR1*, *MS4A4A*, *OAS1* and *SPI1* (extracted from the Supplementary data of Harwood *et al.* (2021)^20^, and summarised in Supplementary Table 8). Furthermore, this approach also identified new risk genes not previously associated with AD, such as *GEMIN7* (frontal cortex, p = 2.17e-11, Z = 6.69) and *MTCH2* (frontal cortex, p = 2.44e-10, Z = −6.33) (extracted from the Supplementary data of Harwood *et al.* (2021)^20^ and summarised in Supplementary Table 8). *GEMIN7* is in the same region as *APOE*, but is included here as a gene of interest because it shows transcription-dependent association. Using the longevity GWAS, we noted known genes showing variation associated with longevity including *FOXO3*, where reduced *FOXO3* expression was significantly associated with longevity in whole blood (Fig. 6 and Supplementary Fig. 5). Additionally, we identified several genes associated with altered expression and longevity, such as *ADD1*, a cytoskeletal protein which achieved genome-wide significance in the CD14-positive monocyte cell dataset, with increased expression being linked to longevity (Fig. 6 and Supplementary Fig. 5). *CASP8* also achieved genome-wide significance in cortical tissue (frontal cortex, p = 3.71e-08, Z = −5.50; Fig. 6 and Supplementary Table 9), and blood, with decreased expression linked to longevity (Supplementary Fig. 5). Cathepsin S (*CTSS*) was also significant in the putamen, with increased expression linked to longevity (Supplementary Fig. 5 and Supplementary Table 9). This approach has highlighted new genes associated with longevity (p < 0.01 and Z > 3.0 or < −3.0), of which around half are thought to interact via Ingenuity pathway analysis including *APOE, CASP8, CTSB, CTSH, CTSS, LIPA, LPL, NPC1* and *TUFM* (Fig. 7 and Supplementary Table 9). Given that *FOXO3*, *ADD1* and *CASP8* are expressed by homeostatic microglia, and that genetic variation associated with longevity is enriched in genes expressed by homeostatic microglia, our collective data suggest a model whereby genes associated with longevity are involved in the homeostatic functions of microglia, and perhaps other innate immune cells. If these innate immune cells escape this state of homeostasis by activating stimuli such as amyloid pathology and age-dependent myelin fragmentation, then genes associated with AD determine how microglia are activated and how they respond to these large disturbances in homeostasis (Fig. 8).

**Figure 6.**
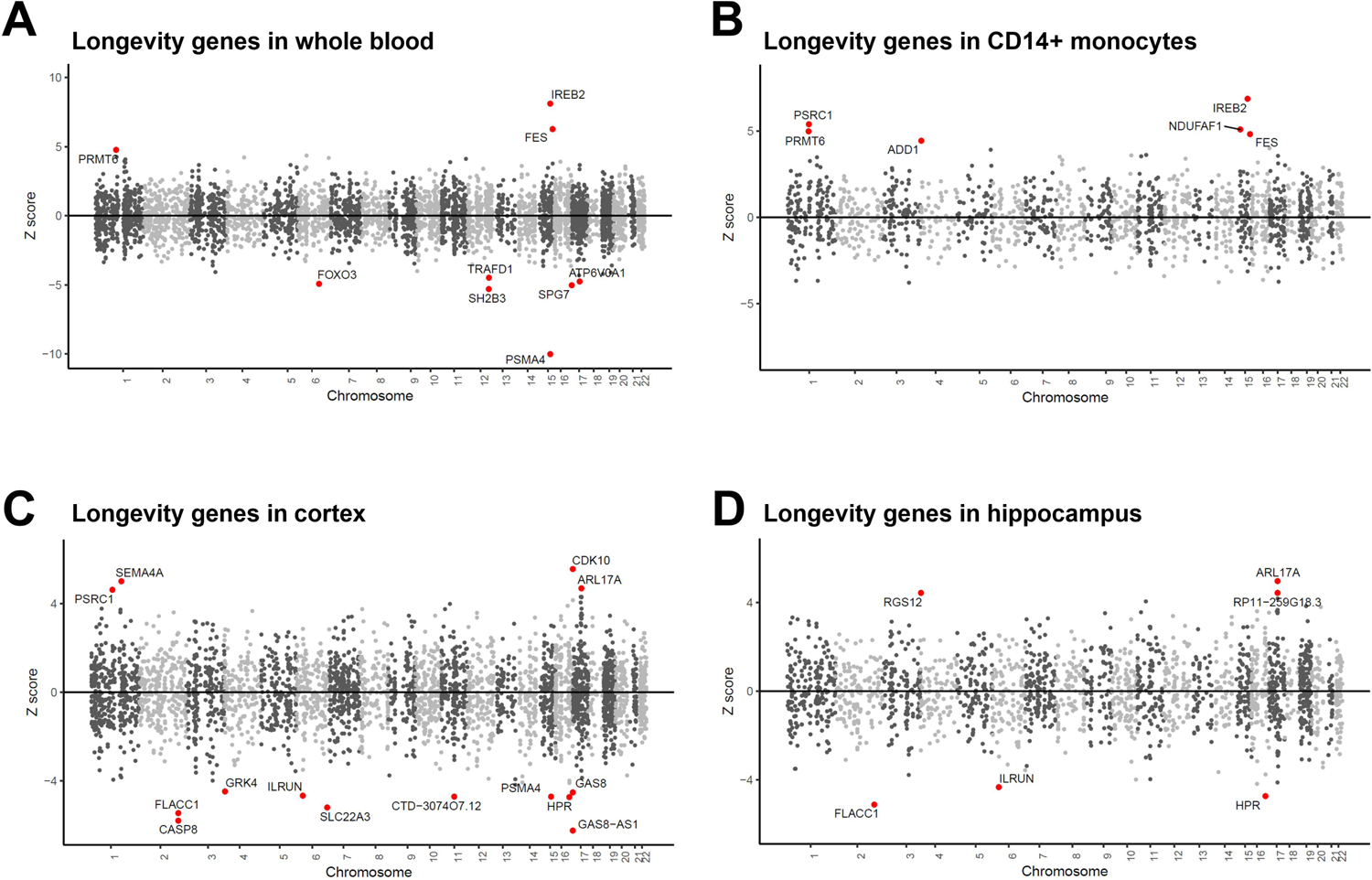
Manhattan plots of TWAS results identifying genes whose expression is associated with longevity in whole blood, CD14+ve myeloid cells, cortex and hippocampus. The y axis is the Z-score of the association between gene expression and longevity in whole blood from YFS study^50^ (A), brain cortex from the GTEx Project^43, 49^ (B), naïve (CD14) monocytes from the Fairfax dataset^48^ (C), and hippocampus from the GTEx Project^43, 49^ (D). Genes that showed significant association following Bonferroni correction for multiple testing are shown with red (ageing risk genes).

**Figure 7.**
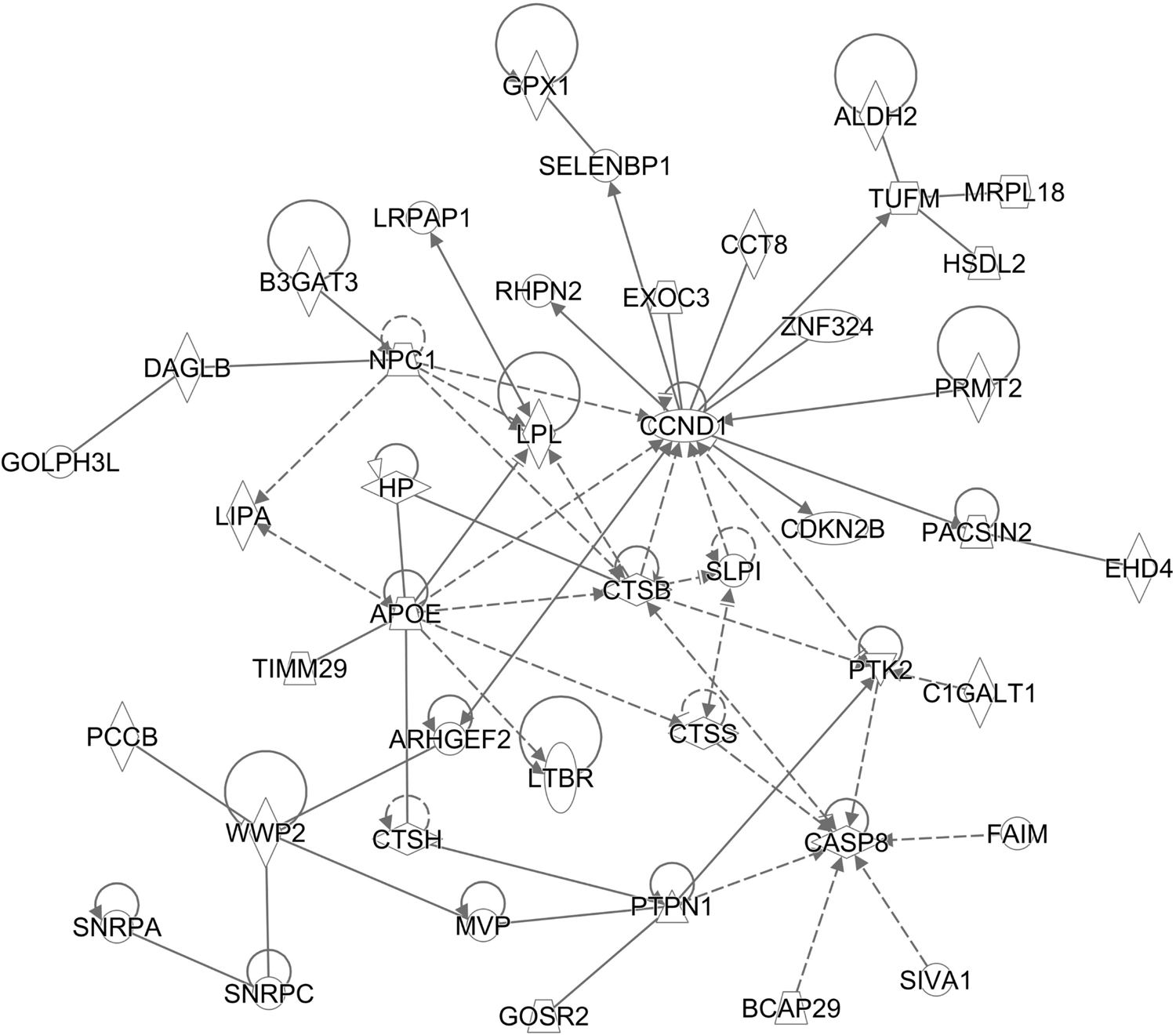
Ingenuity plot showing the TWAS hits associated with longevity are linked in a network containing *APOE*. Solid lines indicate direct interactions (*e.g.* protein-protein interactions, or phosphorylation), and broken lines indirect interactions, between pairs of genes across all mammalian species for tissues and cell types curated in Ingenuity (QIAGEN). Longevity-associated genes from TWAS used as input (p < 0.01 and Z > 2.5 or < −2.5; Supplementary Table 9). Default settings used.

**Figure 8.**
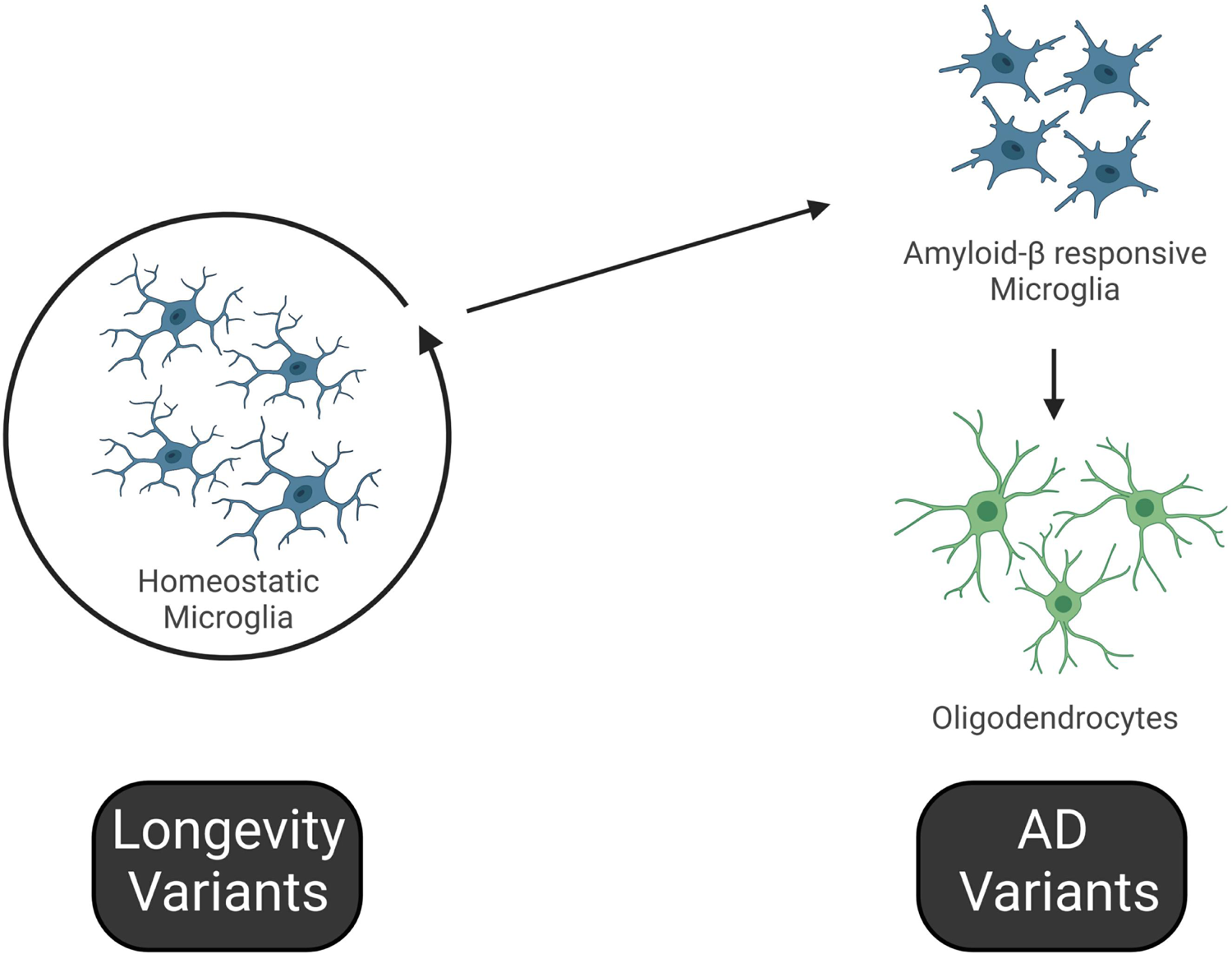
Hypothesis for regulation of homeostatic microglia by genes associated with longevity, and on the other side regulation of microglial activity and oligodendrocyte function by genes associated with AD-risk. Our data suggest a model whereby genes associated with longevity are involved in the homeostatic functions of microglia, and perhaps other innate immune cells, and that if these immune cells escape this state of homeostasis by activating stimuli such as amyloid pathology and age-dependent myelin fragmentation, then genes associated with AD determine how microglia are activated and how microglia interact with remyelinating oligodendrocytes. The age-associated genes we identified in this study that putatively drive the genetic networks associated with innate immune homeostatic processes and activation may underlie “inflammageing.”

## Discussion

### Human common gene-level variation associated with AD is enriched in age-dependent oligodendrocytic and microglial co-expression networks

Human ageing and AD are so entwined due to numerous interconnected co-morbidities, such as vascular disease, and the long pre-symptomatic period of AD highlights the challenge of studying each phenomenon independently. A clear clue that links ageing and AD is that traditional GWAS have repeatedly demonstrated that *APOE* is the top genetic risk factor contributing to both late-onset AD^21^ and longevity^10, 11^, with the ɛ4 isoform being detrimental in both contexts, showing the highest risk for AD (with earlier age of onset among the late-onset AD cases)^52^, and the lowest odds for longevity. Additionally, longevity shows a significant negative genetic correlation with AD^10^. However, the molecular mechanisms linking AD and ageing are poorly understood. To gain new insights into mechanisms linking ageing and AD, we have combined genetic association in human GWAS data with transcriptomic analyses of ageing in the human and mouse hippocampus, aiming to identify biological pathways and cell types which are both altered during ageing, and associated with longevity and/or AD-risk. We show that the expression of separate gene modules associated with microglia and oligodendrocytes are strongly positively correlated to age in both species. These age-dependent microglial and oligodendrocytic co-expression networks in mice show modest preservation in humans. Notably, only the human modules were significantly enriched for genetic variation associated with AD risk. This implies that microglial and oligodendrocytic ageing may contribute to AD risk, where genetic variants which alter the response of these cell-types to ageing in the human hippocampus may modulate the onset of AD pathogenesis. While the influence of microglia on AD risk is well known^53^, the relationship between oligodendrocytes and myelination on AD risk is less well understood. Although it has been previously suggested that individuals with white matter abnormalities show a higher risk of dementia^54^, and disruption of the expression and epigenetic marks of oligodendrocytic genes is seen in AD and ageing^15, 55–59^.

We highlight new genes not previously reported to be associated with AD, and show that they may have a role in age-related responses. Although, we cannot discount the possibility that these modules are associated with AD due to the same genes being involved in the response to both Aβ-associated cellular damage during the AD process, and in the cellular response to ageing. The hub genes for these modules, and transcription factors whose targets are enriched within these modules, are putative drivers, and potential targets to influence these AD-related modules.

We identified gene expression networks that were significantly associated with age from different bulk and scRNA-seq datasets representing distinct cell-types and biological processes using wild-type mice to reduce the confounds of co-morbidities associated with human tissue. We find that common genetic variation associated with longevity is modestly enriched in genes expressed by homeostatic microglia. Collectively, our bulk and single-cell analyses suggest a new hypothesis whereby genes associated with longevity in the human population may be most active in homeostatic microglial processes, or may control how readily innate immune cells exit the homeostatic state, while genes associated with AD are involved in responses to substantial disturbances of CNS homeostasis, such as age-related myelin breakdown and amyloid pathology (Fig. 8). The microglial-expressed genes may govern whether microglia can resolve the myelin turnover and facilitate healthy ageing, or whether microglia fail to repair myelin damage, leading to CNS dyshomeostasis and potentially then to AD. This two-hit hypothesis involving control of microglial homeostatic functions, and then activation of microglia to support oligodendrocyte function may underlie how ageing and AD are linked. Discovering these genes showing variation associated with age may allow us to predict ‘physiological-age’ versus ‘chronological age’ in the brain, and thus to better predict AD and other age-related diseases. These insights may also lead to intelligent design of new biomarkers to track disease progression and new mechanistic insights for drug development.

### Differences between human and mice age-related microglial responses

Many studies have reported differences in microglial responses to AD-associated features between mice and humans, with characterisation of human AD microglia (HAM), and other related work^41, 60^. Similarly, our preservation analysis showed only modest preservation of age-dependent microglial networks between human and mice. ARM/DAM are thought to be protective in mouse models of AD, thus the lack of the ARM/DAM profile at the end stages of disease could be a distinguishing factor for human AD. In our analysis we see a number of key genes are present in both the human and mouse age-dependent microglial network including *AIF1, C3, CD33, CLEC7A, CTSS, CX3CR1, IRF8, LAPTM5, P2RY13, TLR3* and *TREM2*. Genes unique to the mouse age-dependent microglial network included *B2m, Cst7, Pik3cg* and *Ccl4,* and also *Il33* which is expressed mostly by oligodendrocytes. IL-33 has been shown to have beneficial effects on synaptic plasticity, microglial clearance and AD-associated pathology in rodent models^61–63^. In contrast, genes unique to the human age-dependent microglial network included some familiar genes known to be involved or implicated in AD, including *ABI3, C1QA-C, CSF1R, ITGAM, MRC1, MS4A6A, RUNX1, SAMSN1, SPI1* and *TYROBP*. An intriguing possibility is the stronger age-dependency of some of these genes in humans contribute to progression of AD. Our analysis of biological annotations associated with the age-dependent genes unique to either mouse or human, revealed that age-dependent genes unique to mouse were associated with antigen presentation involving MHC. In contrast, age-dependent genes unique to humans were associated with leukocyte activation, cytokine production and T cell activation. Recent studies have indicated that T cells expand in the brain during AD and ageing, and may contribute to disease progression^64–68^. Thus, microglial genes especially age-regulated in humans may contribute to ageing and AD, in part by recruitment and/or activation of other immune cells, including T cells.

### Human common gene-level variation associated with longevity is enriched in an age-dependent single-cell homeostatic microglial co-expression network

While there are clearly differences between human and mouse microglial responses to ageing, the separation of genes into microglial functions afforded by our co-expression analysis of single-cell mouse microglial transcriptomes, combined with our analysis of longevity associated genes, highlighted that homeostatic functions of innate immune cells may be a significant contributor to longevity. Potentially, the loss of these functions during ageing, possibly triggered in the hippocampus by microglial activation to respond to age-related demyelination, may disrupt brain homeostasis, ultimately leading to less healthy ageing and an earlier death. This is a novel suggestion, but is not totally surprising, as markers of neuronal damage are positively associated with mortality in elderly individuals^69^, and microglia’s homeostatic functions maintain neuronal function and neurogenesis^70–72^. Additionally, by conducting this analysis and grouping co-expressed genes into modules enriched for associations to specific biological annotations and populations of microglia, we provide a resource for researchers to gain a preliminary idea of a potential function of their gene-of-interest, simply by looking up in which module it is expressed. As this approach was no doubt hampered by the sparse nature of scRNA-seq data, we foresee it to become even more capable to distinguish groups of co-expressed genes involved in similar cellular processes as technology improves.

We particularly highlight a potential role for microglia in mediating at least some of *ATXN2* and *FOXO3*’s impact on longevity, which requires further exploration. *FOXO3* has previously been reported to augment microglial proliferation, activation, and apoptotic injury^73^. Therefore, lower *FOXO3* levels could lead to longevity, as indicated by our TWAS of whole-blood samples, by reducing loss of homeostatic myeloid cells during ageing. To our knowledge, *ATXN2* function has not been investigated in microglia, although it is expressed in both mouse (as shown by this study) and human microglia (Myeloid Landscape dataset)^41^. Reduction of *Atxn2* expression extended the lifespan and decreased the pathology of a mouse model of neurodegeneration^74^.

CASP8 is a protease with a well-known critical role in cell death mechanisms such as inflammation, necroptosis and apoptosis^75–77^. *CASP8* expression has been associated with several neurodegenerative disorders, in particular AD, Parkinson’s disease, and autoimmune disorders^75, 76, 78, 79^. Several studies demonstrated that LPS primed microglia promoted microglial proliferation, survival and pro-inflammation in a CASP8-dependent manner, associated with neurotoxicity towards neurons^75, 76, 80^. *CASP8* has been reported to mediate Aβ-induced neuronal apoptosis, leading to the characteristic neuronal loss in AD^78^. In a recent study, CASP8 was found to be active in multiple sclerosis (MS) lesions^79^. Surprisingly, while microglial *Casp8* ablation in experimental autoimmune encephalomyelitis mice didn’t affect the course of disease, myeloid *Casp8* ablation worsened the autoimmune demyelination, as CASP8 acts as a negative regulator of the RIPK1/RIPK3/IL-1β pathway. Furthermore, a study of skin fibroblasts showed CASP8 cleaved ATG3 and p63, controlling autophagy and nutrient sensing, which are pathways known to be dysregulated in ageing^81^. This CASP8/AG3/p63 pathway has not been explored in other cell types, which would be beneficial for understanding the role of CASP8 in ageing. Finally, a decline in T cell numbers and impaired antibody production in B cells were observed in mice in the absence of *Casp8*^82^, emphasising its importance in immunity. These findings suggest a cell/tissue-specific role of *CASP8* in the body. Our TWAS data indicate lower levels of *CASP8* in the cortex and cerebellum are associated with longevity which may act by attenuating chronic neuroinflammation and cell death in the brain and thus, promote longevity. While we also saw in our TWAS data with whole blood the opposite effect to that in the cortex, where higher *CASP8* levels were associated with longevity, suggesting that *CASP8* may be beneficial for maintaining the innate and adaptive immune function of peripheral blood cells. It may be that the opposing effects seen for *CASP8* in cortex versus whole blood could be a direct effect of *CASP8* in longevity promotion in one cell type, followed by a secondary, indirect compensatory response in the other cell type.

### Oligodendrocyte function in ageing and AD

Interestingly, changes in the expression of microglial co-expression modules showed remarkable similarity between ageing and responses to demyelination in young mice. Combined with the predominant oligodendrocytic expression of the bulk mouse microglial module’s hub genes, this indicates that microglial responses during ageing could be driven by age-related demyelination. Therefore, it may be the microglial response to age-related demyelination that disrupts their longevity-associated homeostatic networks. Similarly, HAMs seen in human AD patients, have similar transcriptional profiles to microglial populations detected in MS patients, suggesting the possibility that demyelination may be a common phenomenon in ageing, MS, and AD^41^.

Early changes in myelination and oligodendrocytes have been identified with AD^58, 59, 83–85^. Indeed, recent work shows that *APOE* status affects myelination and oligodendrocyte function^86, 87^. Myelin debris is lipid-rich and compacted, and so is difficult to digest by microglia. This may contribute to microglial dysfunction, brain ageing and neurodegeneration. In leukodystrophies (myelin degeneration), activated microglia collect in “nodules,” providing further evidence of the importance of microglia in responding to changes in myelin and axon integrity. White matter-associated microglia have distinct transcriptional profiles, depend on TREM2 signalling and upregulate ARM-associated genes involved in lipid metabolism and phagocytosis (such as *Apoe*, *Cst7*, *Bm2* and *Ctsb*)^13^. This is consistent with the TREM2 receptor binding to a series of ligands related to myelin debris uptake including apolipoproteins and anionic lipids. In our age-dependent human transcriptome networks *TREM2* was central in the microglial network, alongside downstream mediators *TYROBP* and *SYK* and this age-dependent network was enriched with genetic variation associated with AD. Individuals with homozygous loss-of-function of *TREM2* display Nasu-Hakola disease, where leukoencephalopathy is a prominent feature. Amongst our putative risk genes from the ageing-associated TWAS analysis include *EIF2B2*, *NPC1* and *TUFM* which show leukodystrophy as a feature in people inheriting mutations of these genes (Leukoencephalopathy with vanishing white matter, Niemann-Pick disease and combined oxidative phosphorylation deficiency 4, respectively). Thus, promoting myelin integrity and microglial activity to support remyelination is an emerging area of therapeutics for AD^17, 84, 88^.

### Validation by subsequent GWAS studies

A recent GWAS representing the largest association study for dementia identified *ANKH, GRN, PLEKHA1, SNX1* and *UNC5CL* as new genes of interest^89^, which validates our analysis here, as we also identify these genes as putative risk genes for AD, with a much smaller sample size than in the AD GWAS. We identified that the inorganic pyrophosphate transport regulator, *ANKH*, and *PLEKHA1* were expressed by DAM/ARM cells. ANKH is a transmembrane protein involved in pyrophosphate (PPi) and ATP efflux which inhibits excessive mineralization and calcification^90^, and is linked to altered mineralization of tissues, including calcification of the vasculature leading to atherosclerosis. Limited studies have investigated the role of *ANKH in vivo*, but one study addressing vascular calcification in a rat model reported that *ANKH* activity is suppressed by TNF-alpha-activated NF-κB in immune cells which promotes a pro-inflammatory signature, suggesting a role for this gene in regulating inflammation associated with disease^91^. Furthermore, study of a consanguineous family with a novel *ANKH* mutation showed mental retardation in combination with the expected ankylosis and soft tissues, and provided the first evidence of the pathological effect of *ANKH* in the CNS^92^. Recent work has also shown strong association of *ANKH* genetic variation with cognitive change in people with normal ageing and probable AD using fluorodeoxyglucose-PET^93^. *GRN* was confirmed to be a risk gene for dementia^89^, and more specifically frontotemporal degeneration^94^. *GRN* was expressed by microglia transitioning to the DAM/ARM state in our analysis. *GRN* is of interest to the microglial community because loss-of-function mutations have been shown to cause excessive activation of microglia, showing generally opposing effects to TREM2 loss-of-function mutations^95^. *SNX1* is expressed at the highest level by oligodendrocytes. SNX1 is a member of the sorting nexin family (SNX) which is involved in endosomal protein sorting and its dysfunction has been linked to neurodegenerative diseases including AD^96^. *UNC5CL* was also part of the age-dependent oligodendrocyte network but is close to the *TREM2* locus and so may be linked to *TREM2* SNPs^89^.

### Canonical genes involved in neurodegeneration are associated with longevity

Throughout these investigations we saw that many genes associated with different neurodegenerative conditions either showed genetic variation associated with lifespan, or were part of age-dependent co-expression modules, suggesting that these neurodegenerative genes may exert part of their effects in regulating disease in response to specific cellular processes during ageing. We saw that *MAPT* had genetic association with longevity at the gene-level assessed by GWAS SNPs (gene-based p = 0.04), and was significant in our TWAS analysis showing that lower expression of *MAPT* in the cerebellum was associated with longevity (Supplementary Fig. 5). *PSEN1* is a hub gene in the human age-dependent oligodendrocyte network (Fig. 5). *Mobp*, whose human orthologue is a risk gene for tauopathy progressive supranuclear palsy, corticobasal degeneration and amyotrophic lateral sclerosis^97–99^, was a hub gene in the mouse age-dependent oligodendrocyte network (Fig. 1). *HTT*, which shows repeat expansions causing Huntington’s disease, showed genome-wide significance in our TWAS analysis, with lower expression of *HTT* in the whole blood associated with longevity (Supplementary Fig. 5). Similarly, expansion of CAG trinucleotide repeats in the *ATXN2* gene are associated with increased risk of amyotrophic lateral sclerosis^100^, and *ATXN2* was a significant TWAS hit in our study. These findings suggest that common genetic variation in these genes, or their co-expression networks, may also modify lifespan through changes in gene expression between human individuals, in contrast to the familial mutations seen in genes such as *MAPT* and *PSEN1* which alter the amino acid sequence and lead to the protein deposition that is seen in early onset neurodegeneration.

### General immune system changes may govern whole organism ageing

The putative gene variants associated with longevity in this study, although identified by enrichment in the homeostatic microglial network of the brain, are expressed in other immune cells including CD4^+^ T memory T_REG_ cells, natural killer cells, neutrophils, monocytes and macrophages^101–103^. A possibility remains that these identified genes may affect the ageing of the brain and progression of AD by interaction of CNS-resident microglia with other immune cells associated with the blood-brain barrier or throughout the body. T cell changes in the brain that may contribute to ageing and AD have been discussed above^64–67^. Putative ageing-associated genes include *CASP8*, *FES*, *ZKSCAN5*, *JAM3*, *STAT3* and *USP38*, which are expressed by multiple immune cells. It is also possible that the function of these genes in immune cells impact the ageing of the brain via intermediate tissues, such as the vasculature, which has recently been shown to contribute to brain ageing^104, 105^. The immediate next steps of this work are to investigate the array of homeostatic functions of these ageing-associated genes in microglia. Collectively the immune system changes described in our study are likely to contribute to the “Inflammageing” hypothesis of ageing, whereby chronic inflammatory processes promote the ageing of multiple tissues^68, 106^, including the brain.

In conclusion, we identify unbiased co-expression networks in the ageing brain that are enriched for genetic variation associated with AD or longevity to gain new insights into the cell types and cellular processes driving brain ageing and AD. We show that genetic risk for AD is enriched in age-dependent activated microglia and oligodendrocytes. In contrast, we show that genetic variation associated with longevity is enriched in homeostatic microglia. Thus genetic variants associated with longevity and AD both fall on microglia but controlling different and opposing states of their function (homeostatic versus activated). Understanding the function of these priority risk and hub genes may allow us to develop interventions to slow age-related brain decline and AD. These genetic networks driving ageing, AD and inflammageing may help to predict the physiological age of the brain and AD progression more accurately in individual people by generation of biomarker and genotyping assays.

## Methods

### Sample cohorts

*AD*: we used the publicly available GWAS summary statistics data from the International Genomics of Alzheimer’s Projects (IGAP) from the stage 1 GWAS meta-analysis of 21,982 AD cases and 41,944 cognitively normal controls^21^.

*Ageing*: we used the publicly available summary statistics data from European-ancestry GWAS data with overlapping ageing traits, incorporating three GWASs covering healthspan (300,477 individuals, of which 28.3% were not healthy), parental lifespan (1,012,240 parents, of which 60% were deceased), and longevity (defined by 11,262 individuals surviving to the 90th percentile of life, and 25,483 controls whose age at death corresponds to the 60th survival percentile)^10, 11^.

### Gene-based analysis

Gene-based testing was performed using MAGMA v1.08^107^ to obtain gene-based significance (p-values) using GWAS summary statistics. The gene-based approach we used summarizes the strength of the association of multiple adjacent SNPs restricted to individual gene boundaries (sequence between the first and last exons, including the introns), and so accounts for common and complex DNA variants associated with a particular trait, e.g. if longevity is conferred by several (semi) independent SNPs within a locus, each with moderate effect sizes^107, 108^. SNPs were assigned to genes based on the location obtained from the NCBI (no annotation window was added around the genes), and Phase 3 of 1,000 Genome build 37 was used as a reference panel. All required files can be downloaded from the software’s website (https://ctg.cncr.nl/software/magma). Finally, the SNP-wise mean model was used for the analysis. We report the gene-based p-values both before and after the FDR correction.

### RNA-seq data pre-processing

Transcripts per million (TPM) normalised bulk RNA-seq datasets generated from wild-type C57BL/6J mice (8-, 16-, 32-, and 72-weeks-of-age; data available from Mouseac.org)^24^, and non-diseased human hippocampi (from 196 individuals; v8 TPM counts downloaded from gtexportal.org/home/datasets)^43^, were filtered to remove genes with <5% variation between samples, or mean expression levels below 0.5 TPM in all experimental groups. As large differences in gene expression distributions between samples were detected in GTEx data, this data was quantile-quantile normalised using preprocessCore R package’s *normalize.quantiles* function. These bulk RNA-seq datasets were then transformed by log2(normalised counts+1). Batch effects were analysed in bulk RNA-seq datasets by multidimensional scaling using the limma R package’s *plotMDS* function to visualise sample clustering due to sequencing batch. Additionally, correlation of batch covariates to principle components of variation between samples, were identified using the swamp R package’s *prince* function and the CoExpNets R package’s *princePlot* function^23^.

Bulk RNA-seq datasets generated from microglia isolated from the hippocampus of aged and adult wild-type mice^35^ (data available from publication), or the corpus callosum of mice treated for zero, 5-, or 12-weeks with a demyelinating cuprizone diet^40^ (data downloaded from GSO: GSE130627), were normalised using DeSeq2’s estimatesizefactors function^109^. Genes with an average detection level <1.5 normalised counts were removed.

Normalised bulk RNA-seq datasets generated from the hippocampi of wild-type mice of different ages^31–33^, used to assess module preservation, were downloaded from <u>Synapse accession syn20808171</u>, GSO: GSE110741, and GSO: GSE61918, respectively. Genes with an average detection level <0.5 normalised counts or between sample variation <5%, and samples detected as outliers by having p<0.05 result from the outliers R package’s *grubbs.test* function were removed.

Raw counts generated by plate-based scRNA-seq of microglia isolated from the hippocampi of 3-, 6-, 12-, and 21-month-old wild-type mice (downloaded from GSE127893; non-hippocampal and *APP^NL-G-F^* samples were removed)^34^, were filtered to remove cells which appeared either unhealthy (>5% of reads aligned to mitochondrial genes or <1500 transcripts or <1000 genes sequenced) or potential doublets (>3500 genes sequenced). Counts were then transformed by log2(normalised counts+1), and genes not detected in >98.5% of cells in all predefined clusters, or with between sample variation <4%, were removed. Batch effects were assessed by running downstream analysis and determining correlation of generated modules to sequencing plate.

As all datasets demonstrated batch effects, these were removed using limma’s *removeBatchEffect* function. The effects of trait(s)-of-interest were preserved using this function’s design argument. This function’s covariate argument was also used to remove the effects of confounding covariates, extraction batch, sequencing batch, post-mortem interval, sex, cause of death, and centre of origin, from the GTEx dataset.

### Co-expression network analysis

#### Module Construction

Co-expression analysis was performed on pre-processed datasets using CoExpNets’ *getDownstreamNetwork* function. CoExpNets is an optimisation of the popular WGCNA package^22^, which uses an additional k-means clustering step to reassign genes to more appropriate modules, producing more biologically relevant and reproducible modules^23^. Module eigengenes (ME), identified by CoExpNets were correlated (Pearson’s product moment correlation) to numeric traits of interest using CoExpNets’ *corWithNumTraits* function, or to categorical traits using the *corWithCatTraits* function.

#### Biological annotation

Modules formed by co-expression analysis were assessed for enrichment of cell-type specific genes using CoExpNets’ *genAnnotationCellType* function. Module enrichment for biological annotations was assessed using the Gprofiler2 R package’s *gost* function^44^. The genes assigned to the module (ordered by correlation with the module eigengene), were input as the query argument and all genes expressed above the expression threshold (TPM = 0.5 for bulk RNA-seq datasets, and detection in >2.5% of cells in any cell cluster for scRNA-seq datasets), were used as the custom background. Predicted annotations were excluded, and p-values were Bonferroni corrected. Module expression by age was assessed by two-tailed Student’s *t*-test of the mean expression of the module’s 100 most central genes (genes with highest correlation with the module eigengene) in the processed O’Neil *et al.* (2018) dataset^35^, while module expression in cuprizone treatment groups was assessed by one-way ANOVA followed by the pairwise comparison of cuprizone diet timepoints to control diet using Dunnett’s test if the ANOVA indicated a significant difference between treatment groups (p < 0.05), in the processed Nugent *et al.* (2020) dataset^40^.

#### Module preservation

Module preservation between our networks was calculated in pre-processed datasets described above, using CoExpNets’ *preservationoneway* function. Control module size was set to 400. This returned preservation statistics calculated by the WGCNA *modulePreservation* function^110^, of which z.summary (summary of other preservation statistics), was reported unless the two sub-measures of preservation (z.connectivity and z.density) differed significantly. Comparison between human and mouse datasets was preceded by conversion of gene symbols to orthologues using the biomaRt R package^111^. Module eigengenes were detected in query datasets using CoExpNets *getNetworkEigengenes* function.

### Pseudotime analysis

We used the Monocle 2 R package to infer single-cell transcriptional trajectories^38, 112^, based on the expression of genes significantly differentially expressed (qval < 0.025) between cell clusters identified in Ref^34^.

### Enrichment analyses of identified overlapping genes

Enrichment analyses were performed by comparing the number of overlapping genes showing genetic association with AD or longevity derived from summary statistics from Kunkle *et al.* (2019)^21^, and Timmers *et al.* (2020)^10^, respectively, to our transcriptome module gene-sets with gene-sets randomly bootstrapped (Ninterations=100,000), from genes reliably expressed in the bulk RNA-seq (>0.5 TPM in any experimental group), and scRNA-seq datasets (expressed in >2.5% cells from any cluster). The random gene sets were matched to the gene sets of interest by: a) the number of genes in a set, and b) the numbers of independent SNPs per gene. The latter was estimated by MAGMA software as the number of principal components of the SNP data matrix of the gene, pruning away principal components with very small eigenvalues ensuring that only 0.1% of the variance in the SNP data matrix is pruned away. The enrichment analyses p-values (bootstrap p-values in the text) do not require correction for multiple testing because the different age-associated gene modules were pre-selected from different transcriptome datasets (*i.e.* mouse versus human, bulk versus single-cell RNA-seq).

### Transcriptome-wise association study (TWAS)

Expression weights were used in TWAS for autosomal chromosomes and excluding the MHC region, with the longevity summary statistics^10^, using the R script FUSION.assoc_test.R from the FUSION software^113^. The TWAS weights were downloaded from the FUSION website (GTEX7 Brain (13 tissues), GTEX7 Whole blood)^43, 49^, YFS blood^50^, NTR blood^51^, and for all monocytes in the dataset produced by Fairfax *et al.* (2014)^48^ (samples: CD14, LPS2, LPS24 and IFN-gamma), downloaded from https://github.com/janetcharwood/MONOCYTE_TWAS (see Harwood *et al.* (2021)^20^ for details). Bonferroni correction for the number of genes analysed in each tissue was used to determine significance of TWAS associations. Plink v1.9 was used for SNP quality-control analysis, and MAGMA (v1.08) to obtain gene-wide p-values, and Logistic regression was performed in R.

## Data Availability

The R code for performing the data analysis described in this manuscript will be made freely available via GitHub, and all data produced in the present work are contained in the manuscript.

## Acknowledgements

This work was funded by the UK DRI, which receives its funding from the DRI Ltd, funded by the UK Medical Research Council, Alzheimer’s Society and Alzheimer’s Research UK (ARUK). DAS also received funding from the ARUK pump priming scheme via the UCL network. JH is supported by the Dolby Foundation, and by the National Institute for Health Research University College London Hospitals Biomedical Research Centre. The Genotype-Tissue Expression (GTEx) Project was supported by the Common Fund of the Office of the Director of the National Institutes of Health, and by NCI, NHGRI, NHLBI, NIDA, NIMH, and NINDS. The data used for the analyses described in this manuscript were obtained from the GTEx Portal.

## Code and data availability

The R code for performing the data analysis described in this manuscript will be made freely available via GitHub. The data used for this study are publicly available for non-commercial research purposes, accession numbers are given in Methods.

## References

1. Wahl, D. et al. Nutritional strategies to optimise cognitive function in the aging brain. Ageing Res. Rev. 31, 80–92 (2016).

2. Murman, D. L. The Impact of Age on Cognition. Semin. Hear. 36, 111–121 (2015).

3. Fan, X., Wheatley, E. G. & Villeda, S. A. Mechanisms of Hippocampal Aging and the Potential for Rejuvenation. Annu. Rev. Neurosci. 40, 251–272 (2017).

4. Volianskis, A. et al. Long-term potentiation and the role of N-methyl-D-aspartate receptors. Brain Res. 1621, 5–16 (2015).

5. Partridge, L., Fuentealba, M. & Kennedy, B. K. The quest to slow ageing through drug discovery. Nature Reviews Drug Discovery 19, 513–532 (2020).

6. Middeldorp, J. et al. Preclinical assessment of young blood plasma for Alzheimer disease. JAMA Neurol. 73, 1325–1333 (2016).

7. Salih, D. A. & Brunet, A. FoxO transcription factors in the maintenance of cellular homeostasis during aging. Current Opinion in Cell Biology 20, 126– 136 (2008).

8. Askew, K. et al. Coupled Proliferation and Apoptosis Maintain the Rapid Turnover of Microglia in the Adult Brain. Cell Rep. 18, 391–405 (2017).

9. Udeochu, J. C., Shea, J. M. & Villeda, S. A. Microglia communication: Parallels between aging and Alzheimer’s disease. Clin. Exp. Neuroimmunol. 7, 114–125 (2016).

10. Timmers, P. R. H. J., Wilson, J. F., Joshi, P. K. & Deelen, J. Multivariate genomic scan implicates novel loci and haem metabolism in human ageing. Nat. Commun. 11, 1–10 (2020).

11. Deelen, J. et al. A meta-analysis of genome-wide association studies identifies multiple longevity genes. Nat. Commun. 10, 1–14 (2019).

12. Safaiyan, S. et al. Age-related myelin degradation burdens the clearance function of microglia during aging. Nat. Neurosci. 19, 995–998 (2016).

13. Safaiyan, S. et al. White matter aging drives microglial diversity. Neuron 109, 1100–1117.e10 (2021).

14. Mahmood, A. & Miron, V. E. Microglia as therapeutic targets for central nervous system remyelination. Curr. Opin. Pharmacol. 63, 102188 (2022).

15. White, C. W. 3rd, Pratt, K. & Villeda, S. A. OPCs on a Diet: A Youthful Serving of Remyelination. Cell Metab. 30, 1004–1006 (2019).

16. Lloyd, A. F. et al. Central nervous system regeneration is driven by microglia necroptosis and repopulation. Nat. Neurosci. 22, 1046–1052 (2019).

17. McNamara, N. B. et al. Microglia regulate central nervous system myelin growth and integrity. Nature 613, 120–129 (2023).

18. Depp, C. et al. Ageing-associated myelin dysfunction drives amyloid deposition in mouse models of Alzheimer’s disease. bioRxiv 2021.07.31.454562 (2021). doi:10.1101/2021.07.31.454562.

19. Yassa, M. A. et al. Pattern separation deficits associated with increased hippocampal CA3 and dentate gyrus activity in nondemented older adults. Hippocampus 21, 968–979 (2011).

20. Harwood, J. C. et al. Defining functional variants associated with Alzheimer’s disease in the induced immune response. Brain Commun. 3, fcab083 (2021).

21. Kunkle, B. W. et al. Genetic meta-analysis of diagnosed Alzheimer’s disease identifies new risk loci and implicates Aβ, tau, immunity and lipid processing. Nat. Genet. 51, 414–430 (2019).

22. Langfelder, P. & Horvath, S. WGCNA: an R package for weighted correlation network analysis. BMC Bioinformatics 9, 559 (2008).

23. Botía, J. A. et al. An additional k-means clustering step improves the biological features of WGCNA gene co-expression networks. BMC Syst. Biol. 11, 47 (2017).

24. Salih, D. A. et al. Genetic variability in response to amyloid beta deposition influences Alzheimer’s disease risk. Brain Commun. 1, fcz022 (2019).

25. Xue, Z. et al. Genetic programs in human and mouse early embryos revealed by single-cell RNA sequencing. Nature 500, 593–597 (2013).

26. van Dam, S., Võsa, U., van der Graaf, A., Franke, L. & de Magalhães, J. P. Gene co-expression analysis for functional classification and gene–disease predictions. Brief. Bioinform. 19, 575–592 (2018).

27. Ogrodnik, M. et al. Whole-body senescent cell clearance alleviates age-related brain inflammation and cognitive impairment in mice. Aging Cell 20, e13296 (2021).

28. Butler, C. A. et al. Microglial phagocytosis of neurons in neurodegeneration, and its regulation. J. Neurochem. 158, 621–639 (2021).

29. He, D. et al. Disruption of the IL-33-ST2-AKT signaling axis impairs neurodevelopment by inhibiting microglial metabolic adaptation and phagocytic function. Immunity 55, 159–173.e9 (2022).

30. Nguyen, P. T. et al. Microglial Remodeling of the Extracellular Matrix Promotes Synapse Plasticity. Cell 182, 388–403.e15 (2020).

31. Zhao, N. et al. Alzheimer’s Risk Factors Age, APOE Genotype, and Sex Drive Distinct Molecular Pathways. Neuron 106, 727–742.e6 (2020).

32. Sierksma, A., et al. Novel Alzheimer risk genes determine the microglia response to amyloid-β but not to TAU pathology. EMBO Mol. Med. 12, e10606 (2020).

33. Stilling, R. M. et al. De-regulation of gene expression and alternative splicing affects distinct cellular pathways in the aging hippocampus. Front. Cell. Neurosci. 8, 373 (2014).

34. Sala Frigerio, C., et al. The Major Risk Factors for Alzheimer’s Disease: Age, Sex, and Genes Modulate the Microglia Response to Aβ Plaques. Cell Rep. 27, 1293–1306.e6 (2019).

35. O’Neil, S. M., Witcher, K. G., McKim, D. B. & Godbout, J. P. Forced turnover of aged microglia induces an intermediate phenotype but does not rebalance CNS environmental cues driving priming to immune challenge. Acta Neuropathol. Commun. 6, 129 (2018).

36. Keren-Shaul, H. et al. A Unique Microglia Type Associated with Restricting Development of Alzheimer’s Disease. Cell 169, 1276–1290.e17 (2017).

37. Qiu, X. et al. Reversed graph embedding resolves complex single-cell trajectories. Nat. Methods 14, 979–982 (2017).

38. Trapnell, C. et al. The dynamics and regulators of cell fate decisions are revealed by pseudotemporal ordering of single cells. Nat. Biotechnol. 32, 381– 386 (2014).

39. Hill, R. A., Li, A. M. & Grutzendler, J. Lifelong cortical myelin plasticity and age-related degeneration in the live mammalian brain. Nat. Neurosci. 21, 683– 695 (2018).

40. Nugent, A. A. et al. TREM2 Regulates Microglial Cholesterol Metabolism upon Chronic Phagocytic Challenge. Neuron 105, 837–854.e9 (2020).

41. Srinivasan, K. et al. Alzheimer’s Patient Microglia Exhibit Enhanced Aging and Unique Transcriptional Activation. Cell Rep. 31, 107843 (2020).

42. Galatro, T. F. et al. Transcriptomic analysis of purified human cortical microglia reveals age-associated changes. Nat. Neurosci. 20, 1162–1171 (2017).

43. Human genomics. The Genotype-Tissue Expression (GTEx) pilot analysis: multitissue gene regulation in humans. Science 348, 648–660 (2015).

44. Kolberg, L., Raudvere, U., Kuzmin, I., Vilo, J. & Peterson, H. gprofiler2 -- an R package for gene list functional enrichment analysis and namespace conversion toolset g:Profiler [version 2; peer review: 2 approved]. F1000Research 9(ELIXIR), 709 (2020).

45. Sanese, P., Forte, G., Disciglio, V., Grossi, V. & Simone, C. FOXO3 on the Road to Longevity: Lessons From SNPs and Chromatin Hubs. Comput. Struct. Biotechnol. J. 17, 737–745 (2019).

46. Imbert, G. et al. Cloning of the gene for spinocerebellar ataxia 2 reveals a locus with high sensitivity to expanded CAG/glutamine repeats. Nat. Genet. 14, 285– 291 (1996).

47. Fritsch, M. et al. Caspase-8 is the molecular switch for apoptosis, necroptosis and pyroptosis. Nature 575, 683–687 (2019).

48. Fairfax, B. P. et al. Innate immune activity conditions the effect of regulatory variants upon monocyte gene expression. Science 343, 1246949 (2014).

49. Aguet, F., et al. The GTEx Consortium atlas of genetic regulatory effects across human tissues. *bioRxiv* 787903 (2019). doi:10.1101/787903.

50. Raitakari, O. T. et al. Cohort profile: the cardiovascular risk in Young Finns Study. Int. J. Epidemiol. 37, 1220–1226 (2008).

51. Willemsen, G. et al. The Netherlands Twin Register biobank: a resource for genetic epidemiological studies. Twin Res. Hum. Genet. Off. J. Int. Soc. Twin Stud. 13, 231–245 (2010).

52. Liu, C.-C., Liu, C.-C., Kanekiyo, T., Xu, H. & Bu, G. Apolipoprotein E and Alzheimer disease: risk, mechanisms and therapy. Nature reviews. Neurology 9, 106–118 (2013).

53. Sierksma, A., Escott-Price, V. & De Strooper, B. Translating genetic risk of Alzheimer’s disease into mechanistic insight and drug targets. Science 370, 61–66 (2020).

54. Prins, N. D. & Scheltens, P. White matter hyperintensities, cognitive impairment and dementia: An update. Nature Reviews Neurology 11, 157–165 (2015).

55. Murphy, K. B., Nott, A. & Marzi, S. J. CHAS, a deconvolution tool, infers cell type-specific signatures in bulk brain histone acetylation studies of brain disorders. *bioRxiv* 2021.09.06.459142 (2021). doi:10.1101/2021.09.06.459142.

56. Saito, E. R. et al. Alzheimer’s disease alters oligodendrocytic glycolytic and ketolytic gene expression. Alzheimers. Dement. 17, 1474–1486 (2021).

57. Murthy, M. et al. Epigenetic age acceleration is associated with oligodendrocyte proportions in MSA and control brain tissue. Neuropathol. Appl. Neurobiol. 49, e12872 (2023).

58. Vanzulli, I. et al. Disruption of oligodendrocyte progenitor cells is an early sign of pathology in the triple transgenic mouse model of Alzheimer’s disease. Neurobiol. Aging 94, 130–139 (2020).

59. Chacon-De-La-Rocha, I. et al. Accelerated Dystrophy and Decay of Oligodendrocyte Precursor Cells in the APP/PS1 Model of Alzheimer’s-Like Pathology. Front. Cell. Neurosci. 14, 575082 (2020).

60. Gosselin, D. et al. An environment-dependent transcriptional network specifies human microglia identity. Science 356, (2017).

61. Wang, Y. et al. Astrocyte-secreted IL-33 mediates homeostatic synaptic plasticity in the adult hippocampus. Proc. Natl. Acad. Sci. U. S. A. 118, e2020810118 (2021).

62. Lau, S.-F. et al. IL-33-PU.1 Transcriptome Reprogramming Drives Functional State Transition and Clearance Activity of Microglia in Alzheimer’s Disease. Cell Rep. 31, 107530 (2020).

63. Fu, A. K. Y. et al. IL-33 ameliorates Alzheimer’s disease-like pathology and cognitive decline. Proc. Natl. Acad. Sci. U. S. A. 113, E2705–13 (2016).

64. Gate, D. et al. Clonally expanded CD8 T cells patrol the cerebrospinal fluid in Alzheimer’s disease. Nature 577, 399–404 (2020).

65. Lemaitre, P., et al. Molecular and cognitive signatures of ageing partially restored through synthetic delivery of IL2 to the brain. *bioRxiv* 2022.03.01.482519 (2022) doi:10.1101/2022.03.01.482519.

66. Yshii, L., et al. The AppNL-G-F mouse model of Alzheimer’s disease is refractory to regulatory T cell treatment. *bioRxiv* 2022.03.11.483903 (2022). doi:10.1101/2022.03.11.483903.

67. Kaya, T. et al. CD8+ T cells induce interferon-responsive oligodendrocytes and microglia in white matter aging. Nat. Neurosci. 25, 1446–1457 (2022).

68. Borgoni, S., Kudryashova, K. S., Burka, K. & de Magalhães, J. P. Targeting immune dysfunction in aging. Ageing Res. Rev. 70, 101410 (2021).

69. Rübsamen, N. et al. Serum neurofilament light and tau as prognostic markers for all-cause mortality in the elderly general population—an analysis from the MEMO study. BMC Med. 19, 38 (2021).

70. Rogers, J. T. et al. CX3CR1 deficiency leads to impairment of hippocampal cognitive function and synaptic plasticity. J. Neurosci. 31, 16241–16250 (2011).

71. Costello, D. A. et al. Long term potentiation is impaired in membrane glycoprotein CD200-deficient mice: A role for toll-like receptor activation. J. Biol. Chem. 286, 34722–34732 (2011).

72. Bachstetter, A. D. et al. Fractalkine and CX 3CR1 regulate hippocampal neurogenesis in adult and aged rats. Neurobiol. Aging 32, 2030–2044 (2011).

73. Shang, Y., Chong, Z., Hou, J. & Maiese, K. The Forkhead Transcription Factor FOXO3a Controls Microglial Inflammatory Activation and Eventual Apoptotic Injury through Caspase 3. Curr. Neurovasc. Res. 6, 20–31 (2009).

74. Becker, L. A. et al. Therapeutic reduction of ataxin-2 extends lifespan and reduces pathology in TDP-43 mice. Nature 544, 367–371 (2017).

75. Burguillos, M. A. et al. Caspase signalling controls microglia activation and neurotoxicity. Nature 472, 319–324 (2011).

76. Zhang, C.-J. et al. TLR-stimulated IRAKM activates caspase-8 inflammasome in microglia and promotes neuroinflammation. J. Clin. Invest. 128, 5399–5412 (2018).

77. Orning, P. & Lien, E. Multiple roles of caspase-8 in cell death, inflammation, and innate immunity. J. Leukoc. Biol. 109, 121–141 (2021).

78. Wei, W., Norton, D. D., Wang, X. & Kusiak, J. W. Abeta 17-42 in Alzheimer’s disease activates JNK and caspase-8 leading to neuronal apoptosis. Brain 125, 2036–2043 (2002).

79. Kim, S., Lu, H. C., Steelman, A. J. & Li, J. Myeloid caspase-8 restricts RIPK3-dependent proinflammatory IL-1β production and CD4 T cell activation in autoimmune demyelination. Proc. Natl. Acad. Sci. U. S. A. 119, e2117636119 (2022).

80. Xu, S., Zhang, H., Yang, X., Qian, Y. & Xiao, Q. Inhibition of cathepsin L alleviates the microglia-mediated neuroinflammatory responses through caspase-8 and NF-κB pathways. Neurobiol. Aging 62, 159–167 (2018).

81. Sanchez-Garrido, J., Sancho-Shimizu, V. & Shenoy, A. R. Regulated proteolysis of p62/SQSTM1 enables differential control of autophagy and nutrient sensing. Sci. Signal. 11, eaat6903 (2018).

82. Lemmers, B. et al. Essential role for caspase-8 in Toll-like receptors and NFkappaB signaling. J. Biol. Chem. 282, 7416–7423 (2007).

83. Bouhrara, M. et al. Evidence of demyelination in mild cognitive impairment and dementia using a direct and specific magnetic resonance imaging measure of myelin content. Alzheimer’s Dement. 14, 998–1004 (2018).

84. Chen, J.-F. et al. Enhancing myelin renewal reverses cognitive dysfunction in a murine model of Alzheimer’s disease. Neuron 109, 2292–2307.e5 (2021).

85. Sadick, J. S. et al. Astrocytes and oligodendrocytes undergo subtype-specific transcriptional changes in Alzheimer’s disease. Neuron 110, 1788–1805.e10 (2022).

86. Cheng, G. W.-Y. et al. Apolipoprotein E ε4 Mediates Myelin Breakdown by Targeting Oligodendrocytes in Sporadic Alzheimer Disease. J. Neuropathol. Exp. Neurol. 81, 717–730 (2022).

87. Blanchard, J. W. et al. APOE4 impairs myelination via cholesterol dysregulation in oligodendrocytes. Nature 611, 769–779 (2022).

88. Nasrabady, S. E., Rizvi, B., Goldman, J. E. & Brickman, A. M. White matter changes in Alzheimer’s disease: a focus on myelin and oligodendrocytes. Acta Neuropathol. Commun. 6, 22 (2018).

89. Bellenguez, C. et al. New insights into the genetic etiology of Alzheimer’s disease and related dementias. Nat. Genet. 54, 412–436 (2022).

90. Ho, A. M., Johnson, M. D. & Kingsley, D. M. Role of the mouse ank gene in control of tissue calcification and arthritis. Science 289, 265–270 (2000).

91. Zhao, G. et al. Activation of nuclear factor-kappa B accelerates vascular calcification by inhibiting ankylosis protein homolog expression. Kidney Int. 82, 34–44 (2012).

92. Morava, E. et al. Autosomal recessive mental retardation, deafness, ankylosis, and mild hypophosphatemia associated with a novel ANKH mutation in a consanguineous family. J. Clin. Endocrinol. Metab. 96, E189—98 (2011).

93. Nugent, S., Potvin, O., Cunnane, S. C., Chen, T.-H. & Duchesne, S. Associating Type 2 Diabetes Risk Factor Genes and FDG-PET Brain Metabolism in Normal Aging and Alzheimer’s Disease. Front. Aging Neurosci. 12, 580633 (2020).

94. Baker, M. et al. Mutations in progranulin cause tau-negative frontotemporal dementia linked to chromosome 17. Nature 442, 916–919 (2006).

95. Götzl, J. K., et al. Opposite microglial activation stages upon loss of PGRN or TREM2 result in reduced cerebral glucose metabolism. EMBO Mol. Med. 11, e9711 (2019).

96. Zhang, H. et al. The Retromer Complex and Sorting Nexins in Neurodegenerative Diseases. Front. Aging Neurosci. 10, 79 (2018).

97. Höglinger, G. U. et al. Identification of common variants influencing risk of the tauopathy progressive supranuclear palsy. Nat. Genet. 43, 699–705 (2011).

98. Kouri, N. et al. Genome-wide association study of corticobasal degeneration identifies risk variants shared with progressive supranuclear palsy. Nat. Commun. 6, 7247 (2015).

99. van Rheenen, W. et al. Genome-wide association analyses identify new risk variants and the genetic architecture of amyotrophic lateral sclerosis. Nat. Genet. 48, 1043–1048 (2016).

100. Sproviero, W. et al. ATXN2 trinucleotide repeat length correlates with risk of ALS. Neurobiol. Aging 51, 178.e1–178.e9 (2017).

101. Schmiedel, B. J. et al. Impact of Genetic Polymorphisms on Human Immune Cell Gene Expression. Cell 175, 1701–1715.e16 (2018).

102. Schmiedel, B. J. et al. COVID-19 genetic risk variants are associated with expression of multiple genes in diverse immune cell types. Nat. Commun. 12, 6760 (2021).

103. Lavin, Y. et al. Tissue-resident macrophage enhancer landscapes are shaped by the local microenvironment. Cell 159, 1312–1326 (2014).

104. Francis, C. M. et al. Genome-wide associations of aortic distensibility suggest causality for aortic aneurysms and brain white matter hyperintensities. Nat. Commun. 13, 4505 (2022).

105. Wagen, A. Z. et al. Life course, genetic, and neuropathological associations with brain age in the 1946 British Birth Cohort: a population-based study. Lancet Heal. Longev. 3, e607–e616 (2022). doi:https://doi.org/10.1016/S2666-7568(22)00167-2.

106. de Magalhães, J. P. & Passos, J. F. Stress, cell senescence and organismal ageing. Mech. Ageing Dev. 170, 2–9 (2018).

107. de Leeuw, C. A., Mooij, J. M., Heskes, T. & Posthuma, D. MAGMA: generalized gene-set analysis of GWAS data. PLoS Comput. Biol. 11, e1004219 (2015).

108. Escott-Price, V. et al. Gene-Wide Analysis Detects Two New Susceptibility Genes for Alzheimer’s Disease. PLoS One 9, e94661 (2014).

109. Love, M. I., Huber, W. & Anders, S. Moderated estimation of fold change and dispersion for RNA-seq data with DESeq2. Genome Biol. 15, 550 (2014).

110. Langfelder, P., Luo, R., Oldham, M. C. & Horvath, S. Is my network module preserved and reproducible? PLoS Comput. Biol. 7, e1001057 (2011).

111. Durinck, S., Spellman, P. T., Birney, E. & Huber, W. Mapping identifiers for the integration of genomic datasets with the R/Bioconductor package biomaRt. Nat. Protoc. 4, 1184–1191 (2009).

112. Qiu, X. et al. Single-cell mRNA quantification and differential analysis with Census. Nat. Methods 14, 309–315 (2017).

113. Gusev, A. et al. Integrative approaches for large-scale transcriptome-wide association studies. Nat. Genet. 48, 245–252 (2016).

