## Supplementary Figure for "Genetic variation associated with human longevity and Alzheimer’s disease risk act through microglia and oligodendrocyte cross-talk"

#### Supplementary Materials

**Supplementary Figure 1. Genes within the mouse microglial module are expressed by oligodendrocytes.** Dot plot of the expression of the 14 most central genes within the mouse microglial bulk RNA-seq module (14 genes with highest correlation to the module eigengene) with respect to the different cell-types identified in scRNA-seq data of the wild-type mouse hippocampus at 3 and 24 months-of-age[1].

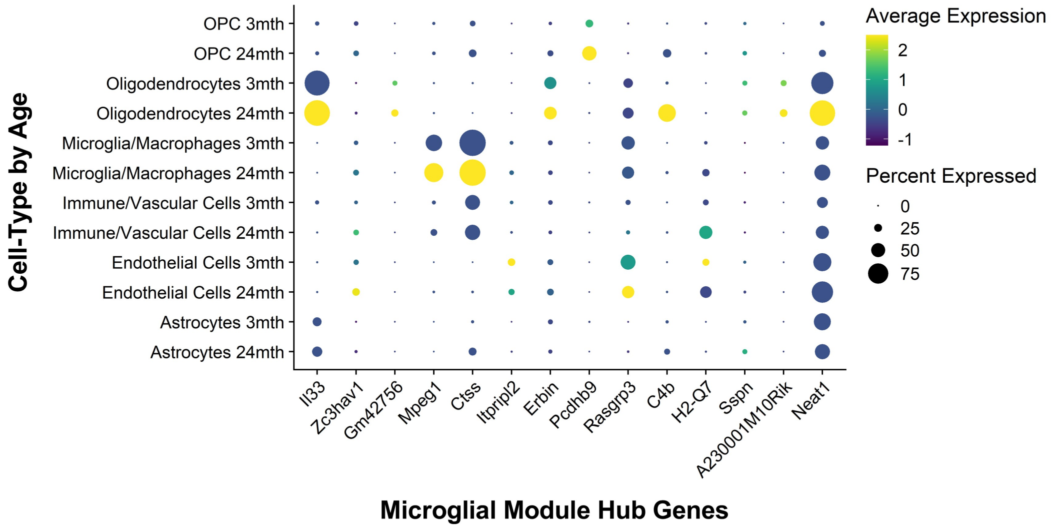


#### Supplementary Figure 1

**Supplementary Figure 2. Preservation of mouse microglial and oligodendrocytic bulk RNA-seq co-expression module's connectivity and correlation to age in publicly available datasets.** A) Preservation analysis determined that the mouse microglial and oligodendrocytic gene networks from our Mouseac data[2] both demonstrate strong preservation (z.summary > 10) of co-expression patterns between module genes in publicly available datasets generated by bulk RNA-seq of hippocampi from wild-type mice of different ages[3–5]. B) Module eigengene (ME) value (first principal component of module expression), calculated for our mouse microglial bulk module from our Mouseac dataset (black line; Salih *et al.* 2019)[2], alongside other datasets which include older mice, which all show strong positive correlations with age; Zhao *et al.* (2020)[3] (Pearson's product-moment correlation = 0.94, $p < 2.2e^{-16}$), and Stilling *et al.* (2014)[5] (Pearson's product-moment correlation = 0.95, $p = 5.8e^{-09}$) datasets. Individual data points represent the ME value in a single sample/mouse from that dataset. C) MEs calculated for our mouse oligodendrocytic module and other datasets also shows strong positive correlation to age in the datasets from Zhao *et al.* (2020)[3] (Pearson's product-moment correlation = 0.86, $p = 7.7e^{-15}$), and Stilling *et al.* (2014)[5] (Pearson's product-moment correlation = 0.58, p = 0.01). Individual data points represent the ME value in a single sample/mouse from that dataset.

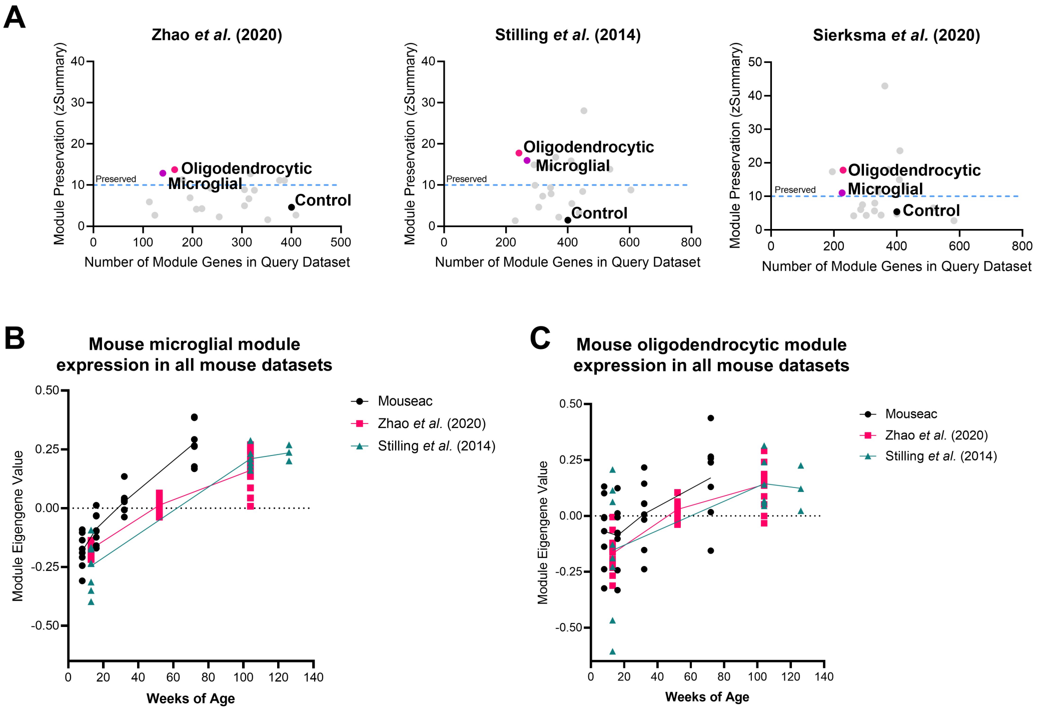


### Supplementary Figure 2

**Supplementary Figure 3. Enrichment of biological annotations representing different aspects of the immune system within the mouse age-related scRNA-seq co-expression modules.** A) ARM-associated module. B) Interferon module. C) HM2-associated module. D) HM1-associated module. E) Phagolysosomal module. F) TGF-β module. G) Ribosomal module.

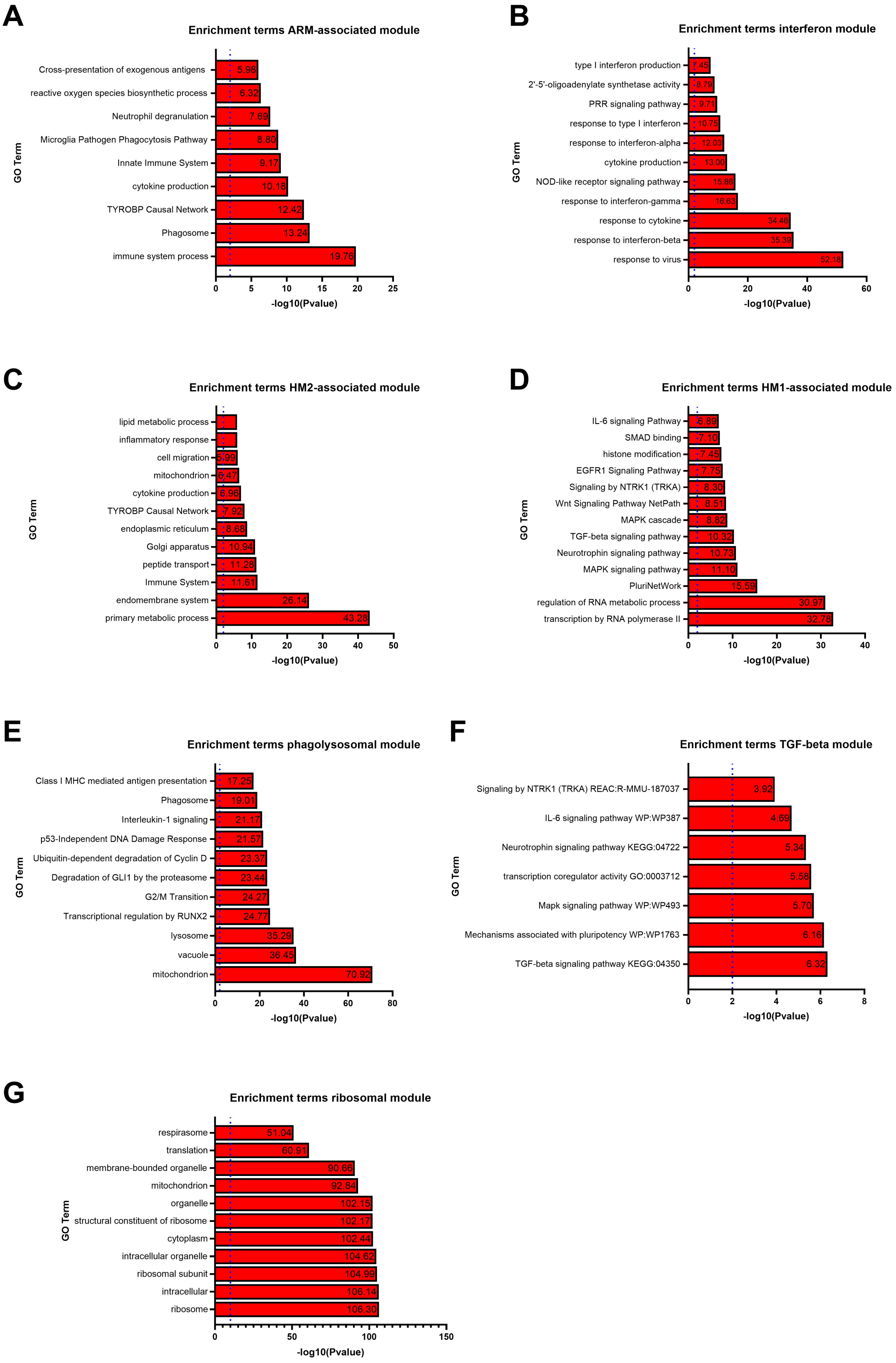


### Supplementary Figure 3

**Supplementary Figure 4. Enrichment of biological annotations associated with microglial age-dependent genetic module of genes expressed uniquely in mice are associated with antigen presentation, and genes expressed uniquely in humans are associated with leukocyte activation, cytokine production and T cell activation.** A) Enrichment terms associated with genes unique to the mouse microglial age-dependent module. B) Enrichment terms associated with genes unique to the human microglial age-dependent module.

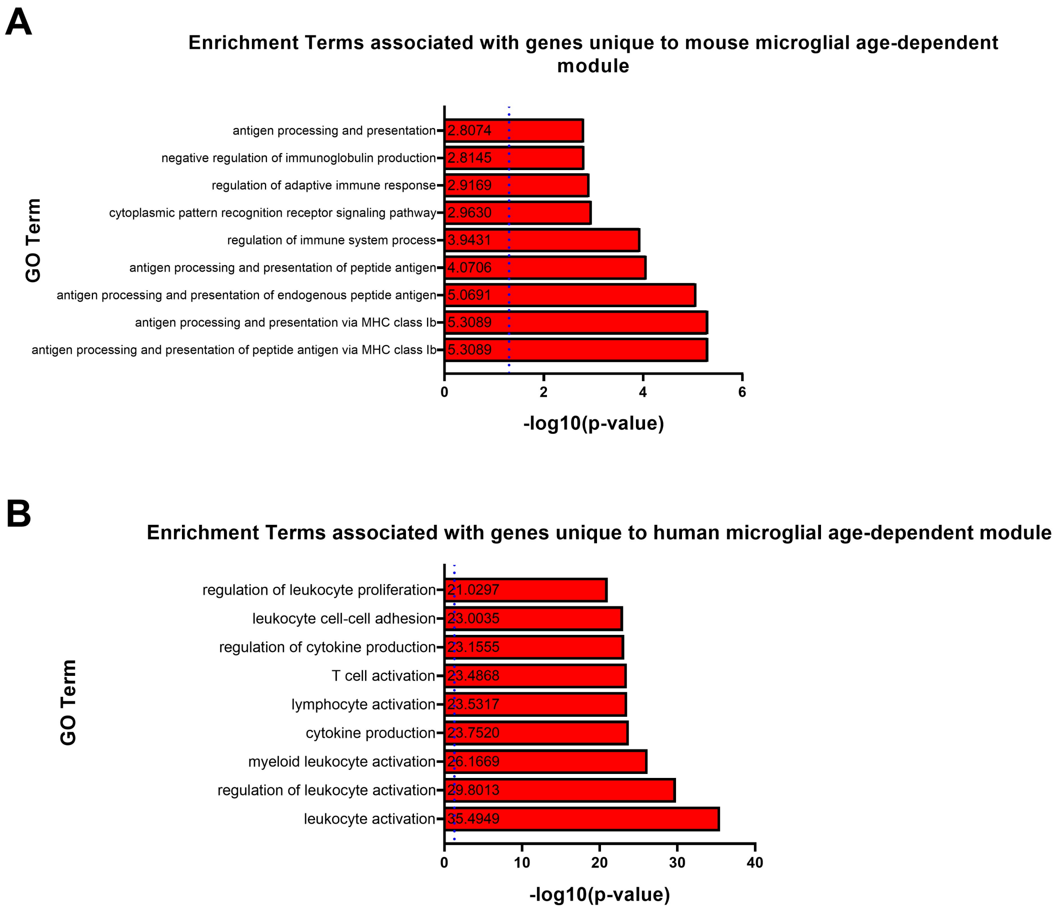


#### Supplementary Figure 4

**Supplementary Figure 5. Manhattan plots of TWAS results identifying genes whose expression is associated with longevity in the GTEX7 brain tissues, GTEX7 whole blood[6,7], YFS blood[8], NTR blood[9], and for all monocytes in the dataset from Fairfax and colleagues (2014)[10] (samples: CD14, LPS2, LPS24 and IFN-gamma).** The y axis is the Z-score of the association between gene expression and longevity in the samples named in top left quarter of plot. Genes that showed significant association following Bonferroni correction for multiple testing are shown with red (ageing risk genes).

**References**


1. Ogrodnik, M. *et al.* Whole-body senescent cell clearance alleviates age-related brain inflammation and cognitive impairment in mice. *Aging Cell* **20**, e13296 (2021). doi:10.1111/acel.13296.
2. Salih, D. A. *et al.* Genetic variability in response to amyloid beta deposition influences Alzheimer's disease risk. *Brain Commun.* **1**, fcz022 (2019).
3. Zhao, N. *et al.* Alzheimer's Risk Factors Age , APOE Genotype , and Sex Drive Distinct Molecular Pathways. *Neuron* **106**, 727-742.e6 (2020).
4. Sierksma, A. *et al.* Novel Alzheimer risk genes determine the microglia response to amyloid-β but not to TAU pathology. *EMBO Mol. Med.* **12**, e10606 (2020).
5. Stilling, R. M. *et al.* De-regulation of gene expression and alternative splicing affects distinct cellular pathways in the aging hippocampus. *Front. Cell. Neurosci.* **8**, 373 (2014).
6. Human genomics. The Genotype-Tissue Expression (GTEx) pilot analysis: multitissue gene regulation in humans. *Science* **348**, 648–660 (2015).
7. Aguet, F. *et al.* The GTEx Consortium atlas of genetic regulatory effects across human tissues. *bioRxiv* 787903 (2019) doi:10.1101/787903.
8. Raitakari, O. T. *et al.* Cohort profile: the cardiovascular risk in Young Finns Study. *Int. J. Epidemiol.* **37**, 1220–1226 (2008).
9. Willemsen, G. *et al.* The Netherlands Twin Register biobank: a resource for genetic epidemiological studies. *Twin Res. Hum. Genet. Off. J. Int. Soc. Twin Stud.* **13**, 231–245 (2010).
10. Fairfax, B. P. *et al.* Innate immune activity conditions the effect of regulatory variants upon monocyte gene expression. *Science* **343**, (2014).
