## Supplementary Fig. 5 for "Genetic variation associated with human longevity and Alzheimer’s disease risk act through microglia and oligodendrocyte cross-talk"

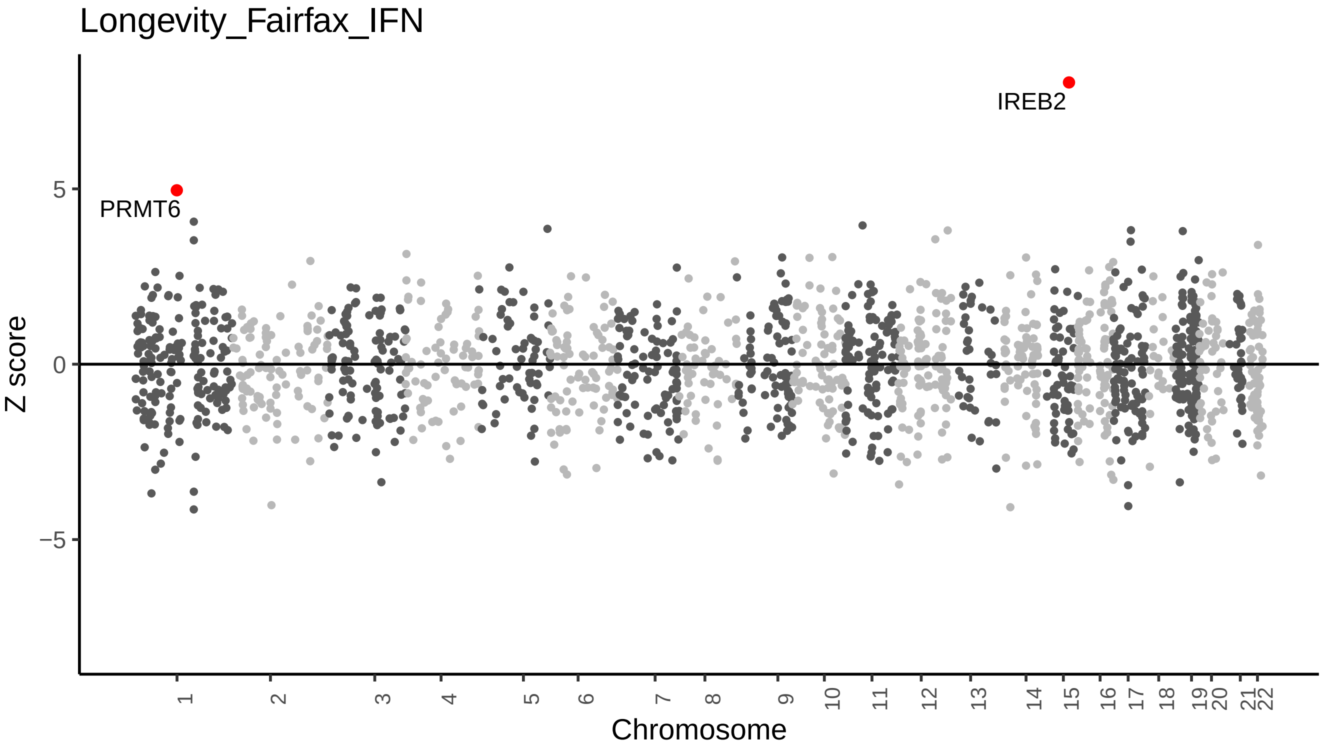

Longevity_Fairfax_IFN
Z score
5
0
−5
PRMT6
IREB2
Chromosome

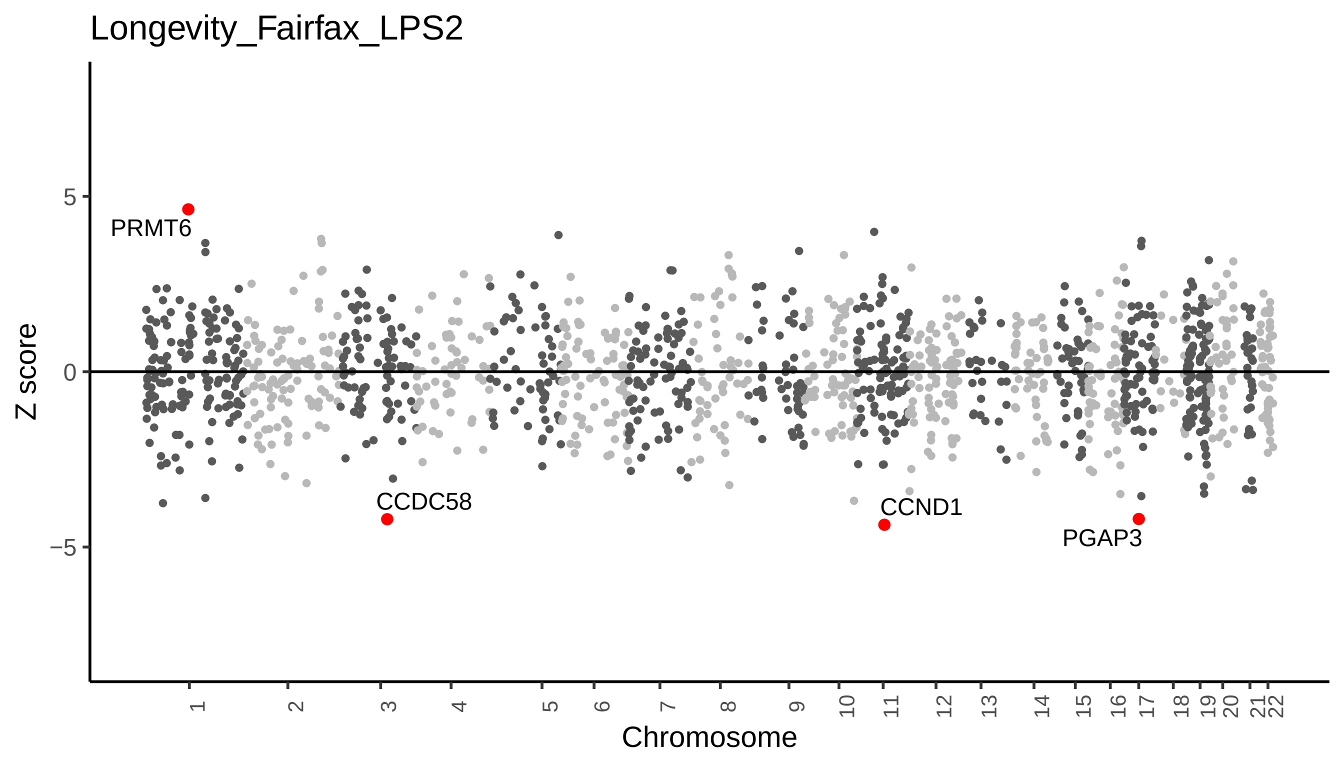

Longevity_Fairfax_LPS2
Z score
5
0
−5
PRMT6
CCDC58
CCND1
PGAP3
Chromosome

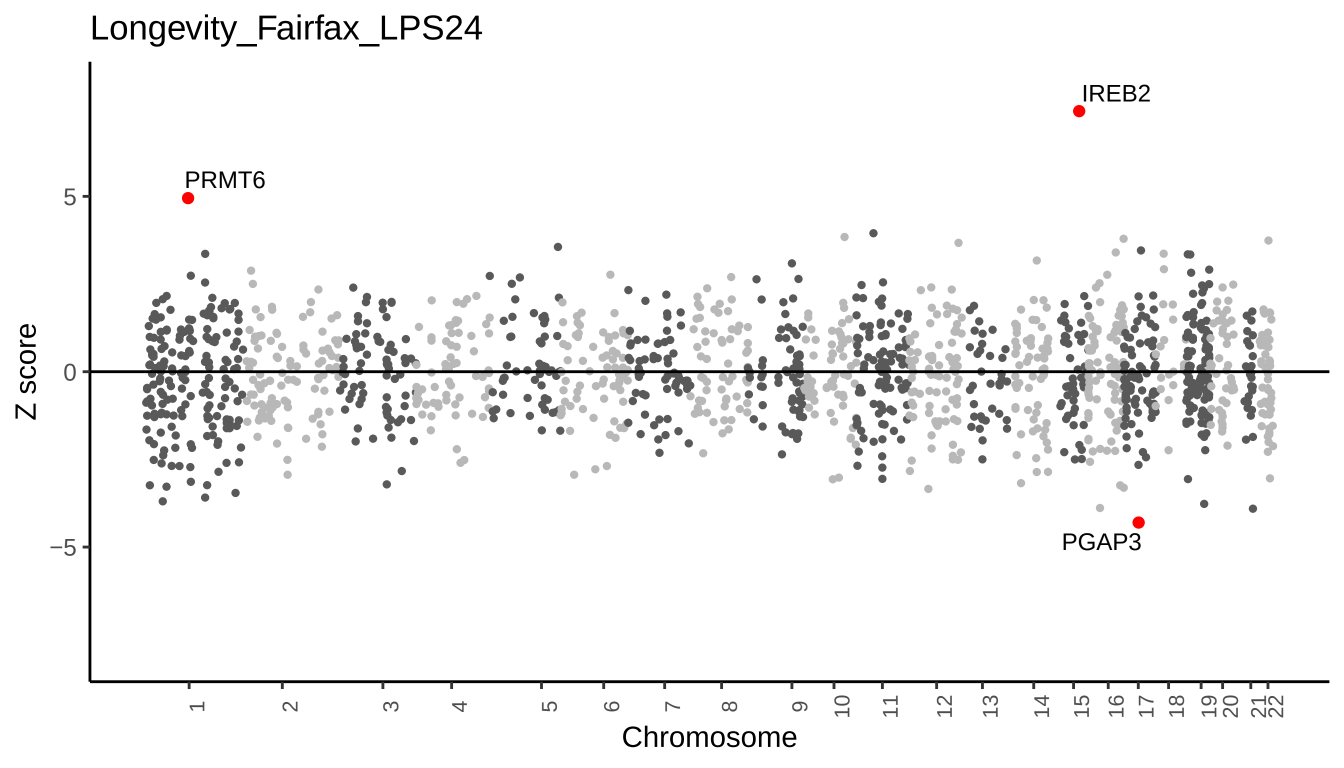

Longevity_Fairfax_LPS24
IREB2
PRMT6
PGAP3
5
0
−5
Z score
Chromosome

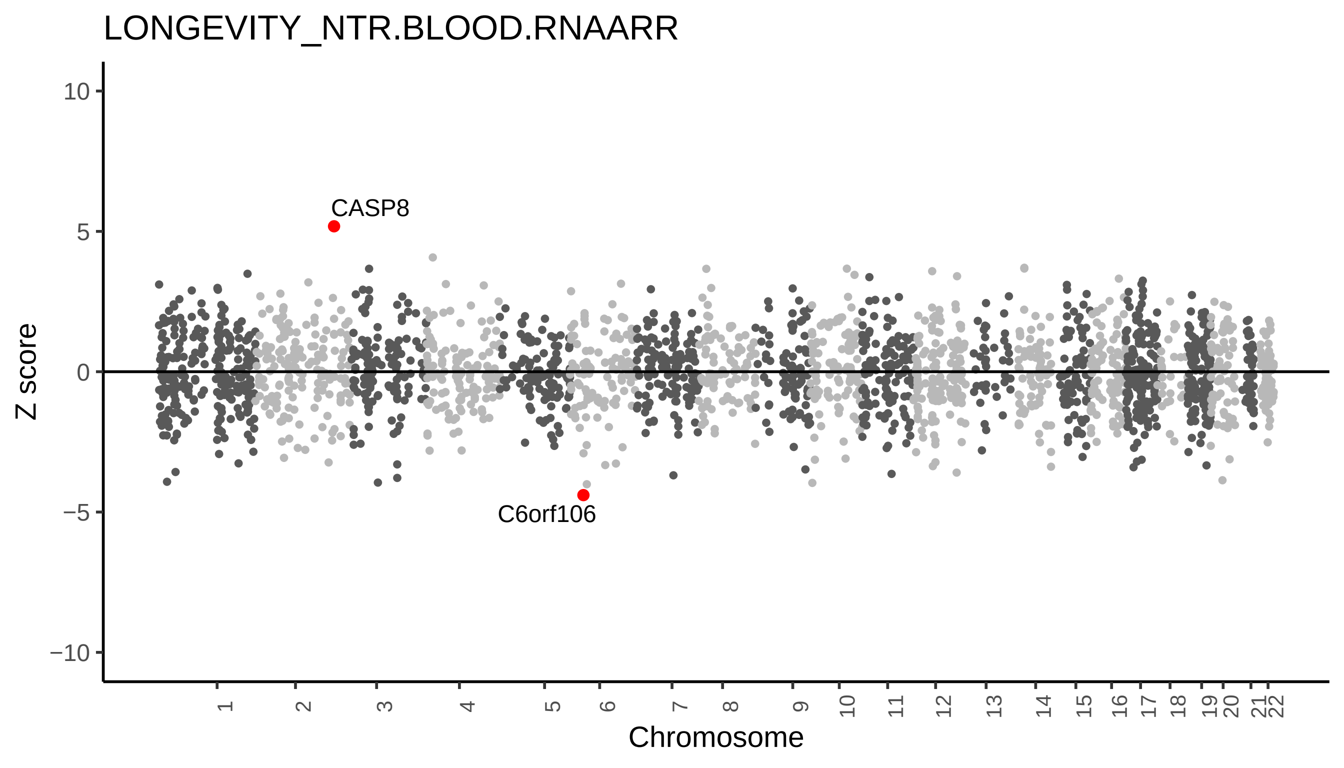

LONGEVITY_NTR.BLOOD.RNAARR
Z score
10
5
0
−5
−10
CASP8
C6orf106
Chromosome

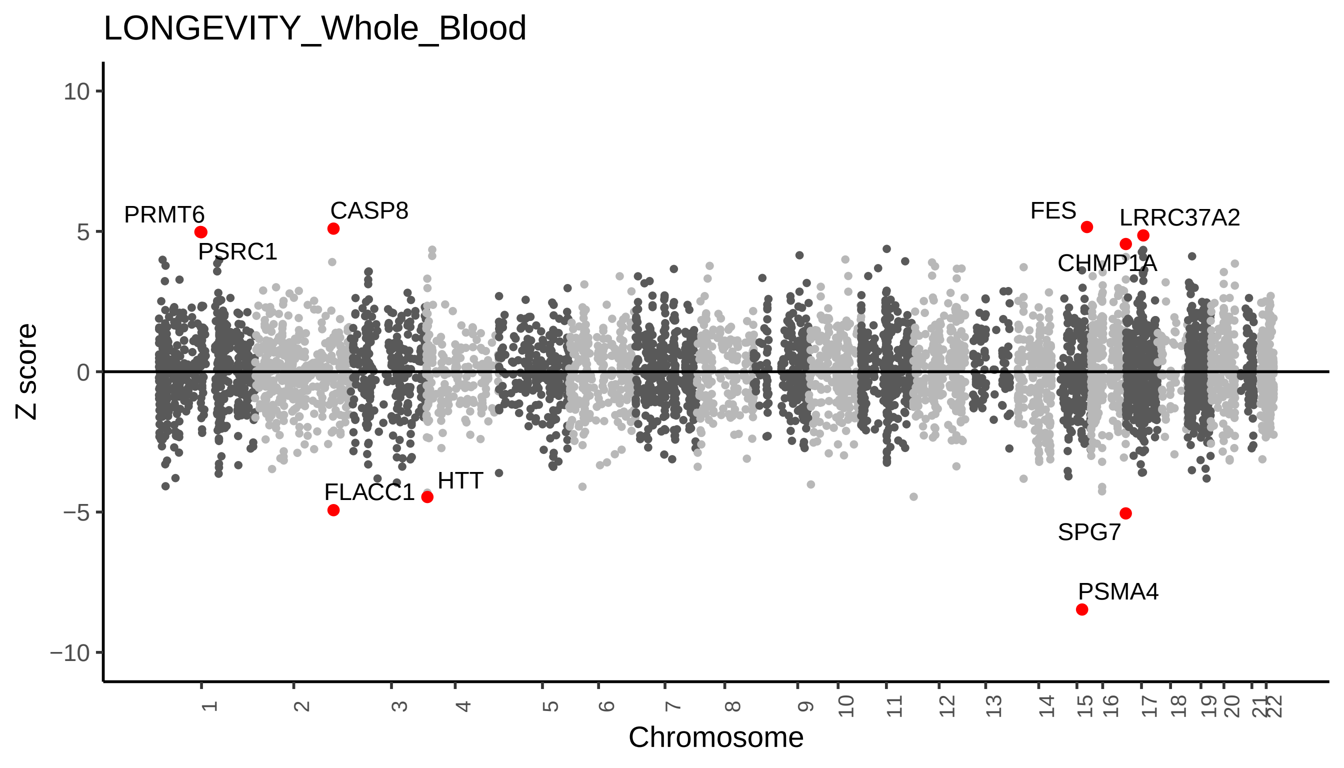
LONGEVITY_Whole_Blood
Z score
Chromosome
10
5
0
−5
−10
PRMT6
PSRC1
CASP8
FES
LRRC37A2
CHMP1A
FLACC1
HTT
SPG7
PSMA4

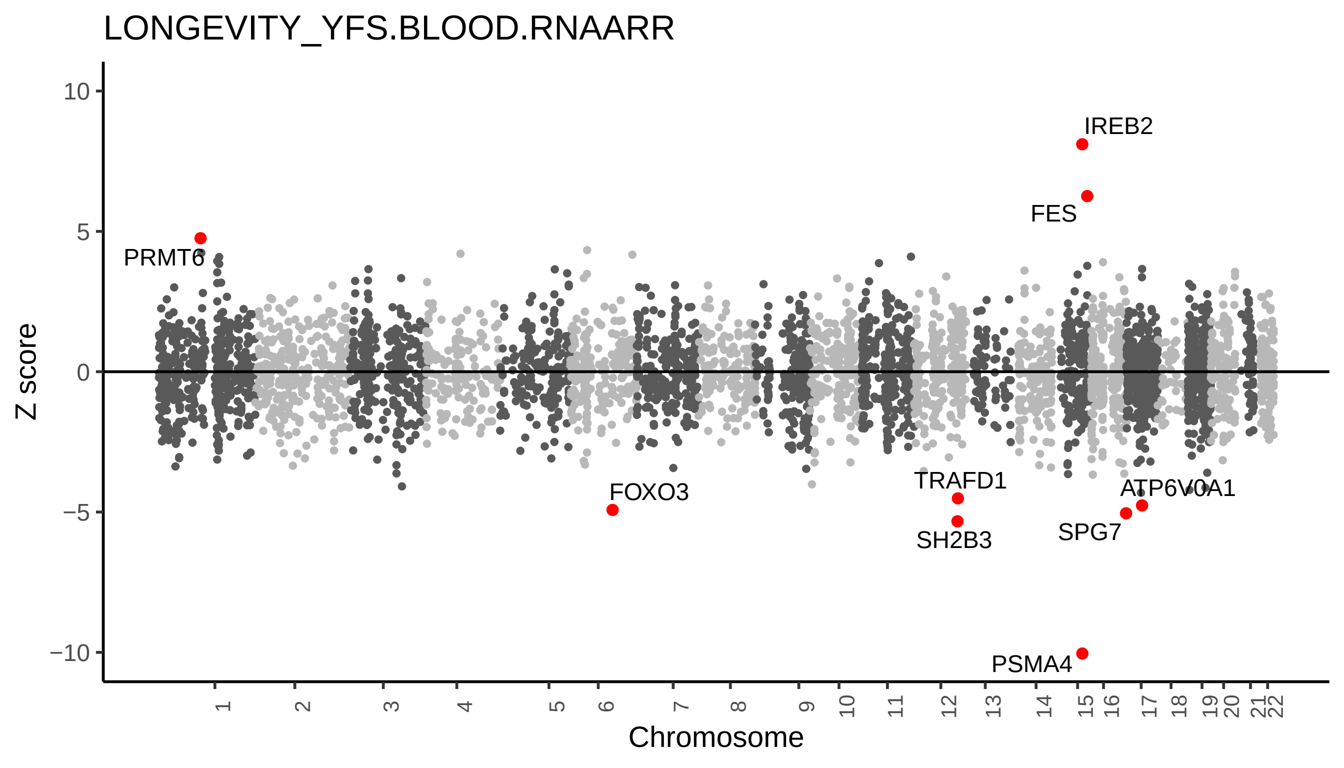

LONGEVITY_YFS.BLOOD.RNAARR
Z score
10
5
0
−5
−10
Chromosome
IREB2
FES
PRMT6
FOXO3
TRAFD1
SH2B3
ATP6V0A1
SPG7
PSMA4

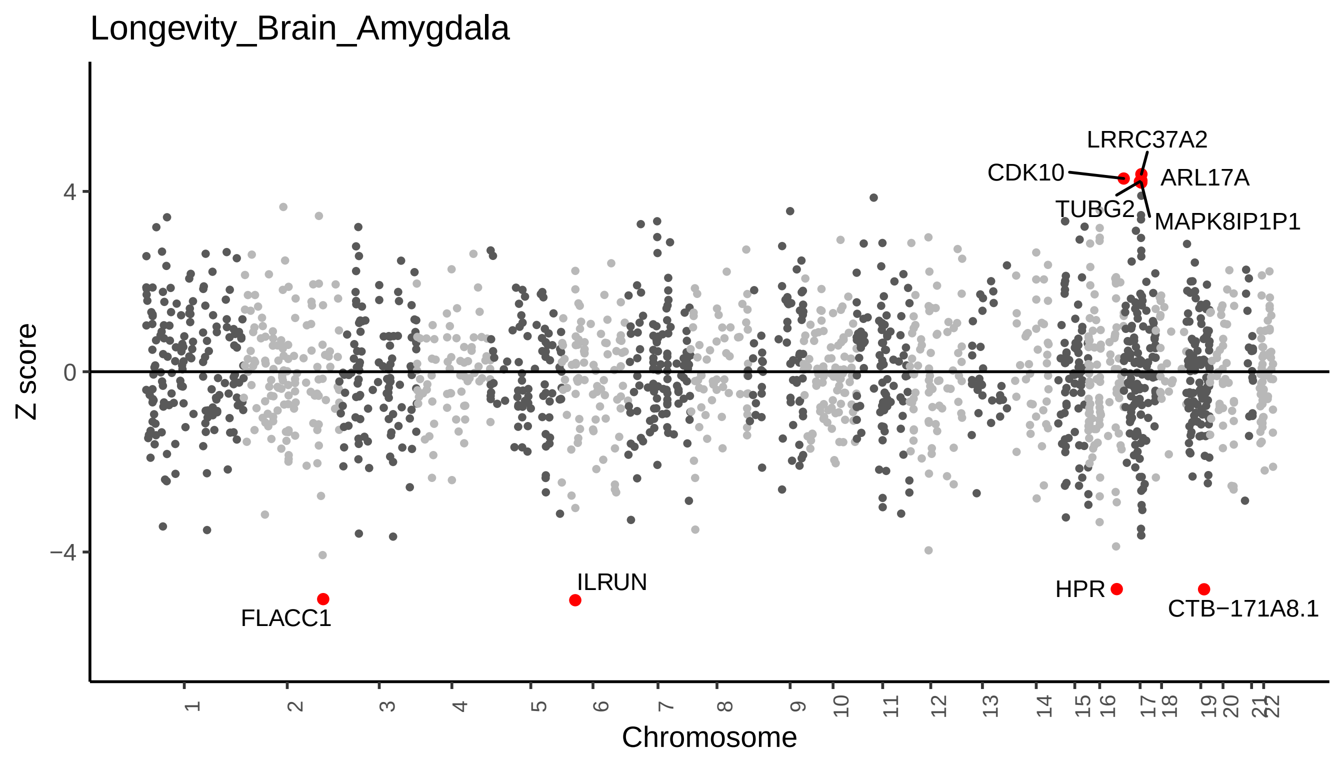
Longevity_Brain_Amygdala
Z score
Chromosome
4
0
−4
LRRC37A2
CDK10
ARL17A
TUBG2
MAPK8IP1P1
ILRUN
FLACC1
HPR
CTB−171A8.1

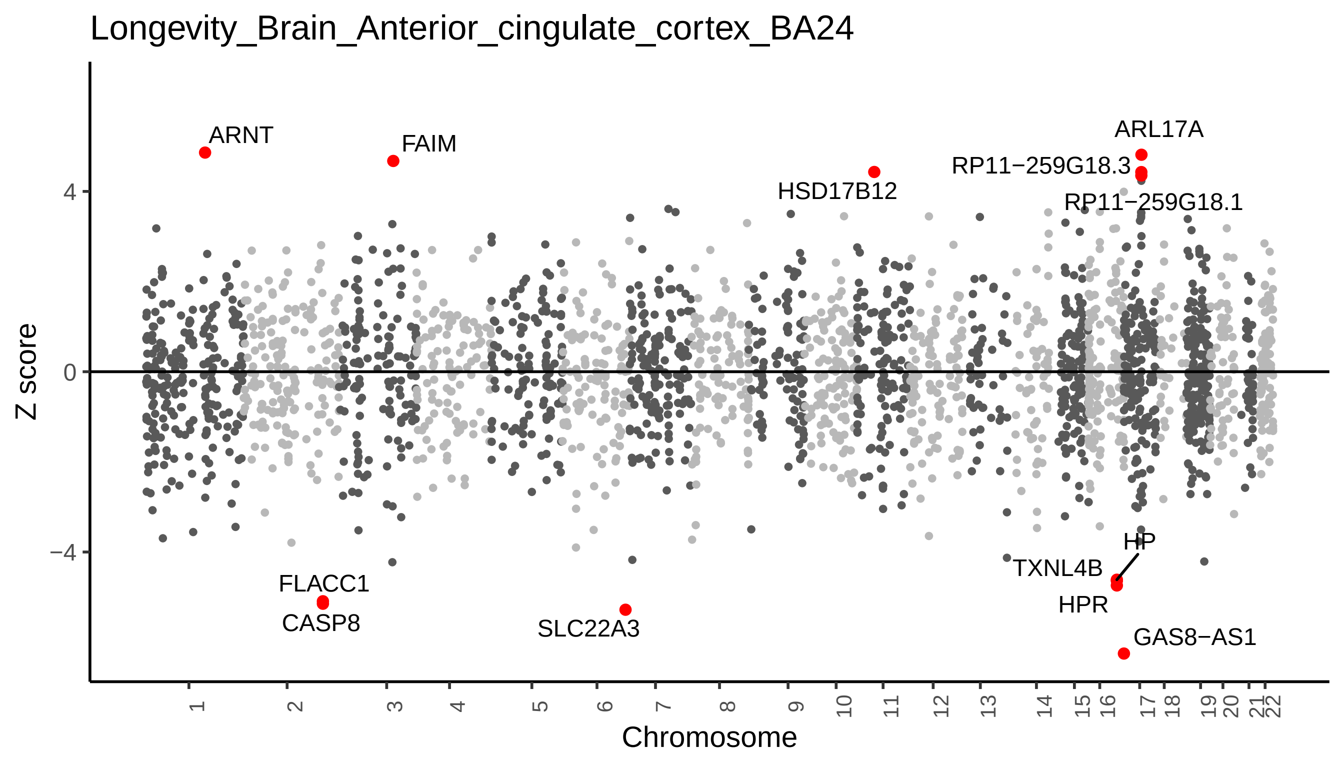

Longevity_Brain_Anterior_cingulate_cortex_BA24
Z score
4
0
−4
ARNT
FAIM
HSD17B12
ARL17A
RP11−259G18.3
RP11−259G18.1
FLACC1
CASP8
SLC22A3
HP
TXNL4B
HPR
GAS8−AS1
Chromosome

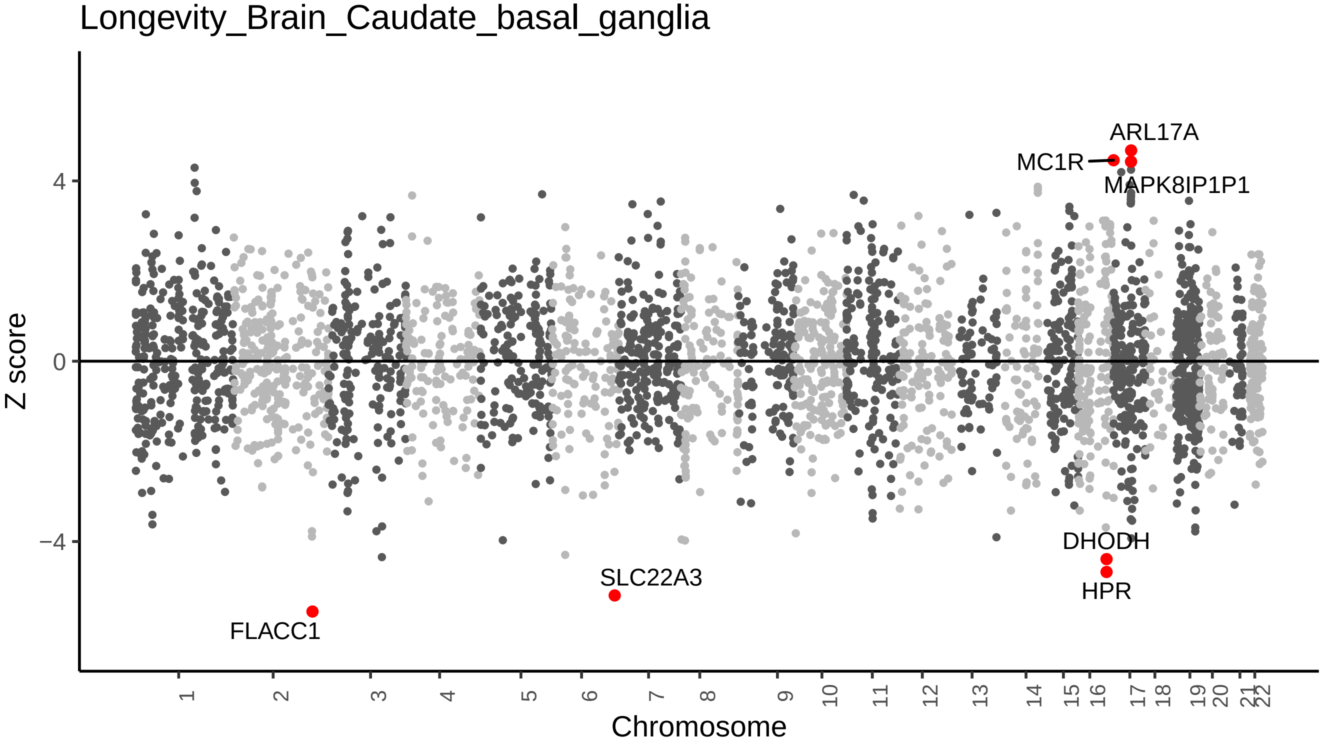

Longevity_Brain_Caudate_basal_ganglia
Z score
4
0
−4
ARL17A
MC1R
MAPK8IP1P1
DHODH
HPR
SLC22A3
FLACC1
Chromosome

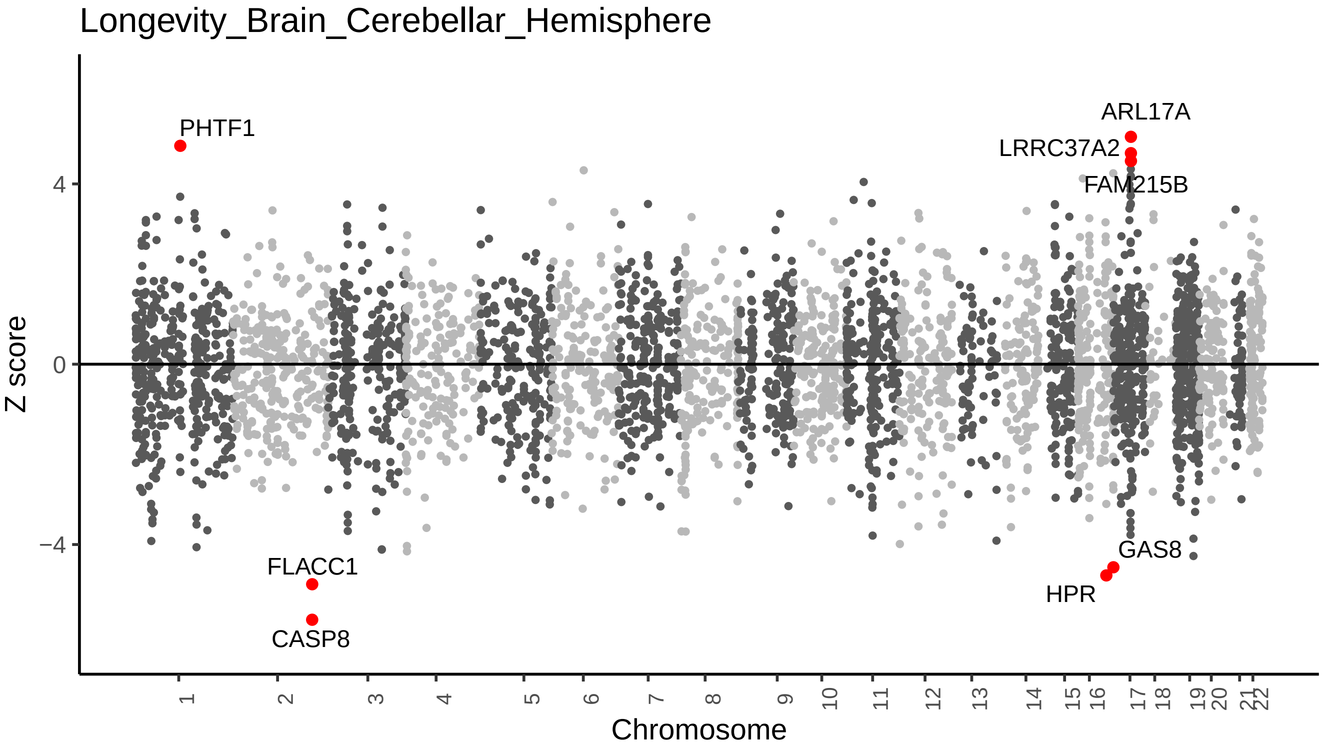
Longevity_Brain_Cerebellar_Hemisphere
Z score
Chromosome
4
0
−4
PHTF1
FLACC1
CASP8
ARL17A
LRRC37A2
FAM215B
GAS8
HPR

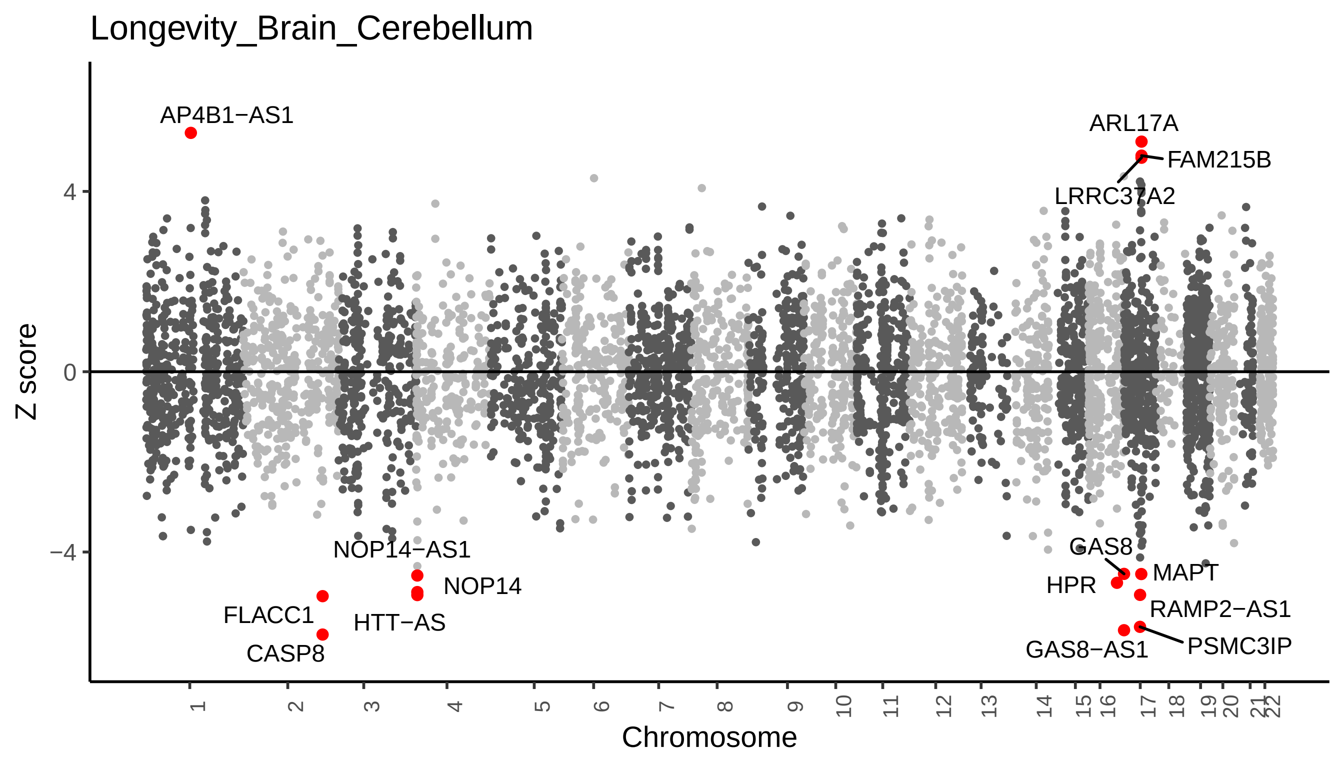
Longevity_Brain_Cerebellum
Z score
4
0
−4
Chromosome
AP4B1−AS1
ARL17A
FAM215B
LRRC37A2
NOP14−AS1
NOP14
HTT−AS
FLACC1
CASP8
GAS8
MAPT
HPR
RAMP2−AS1
PSMC3IP
GAS8−AS1

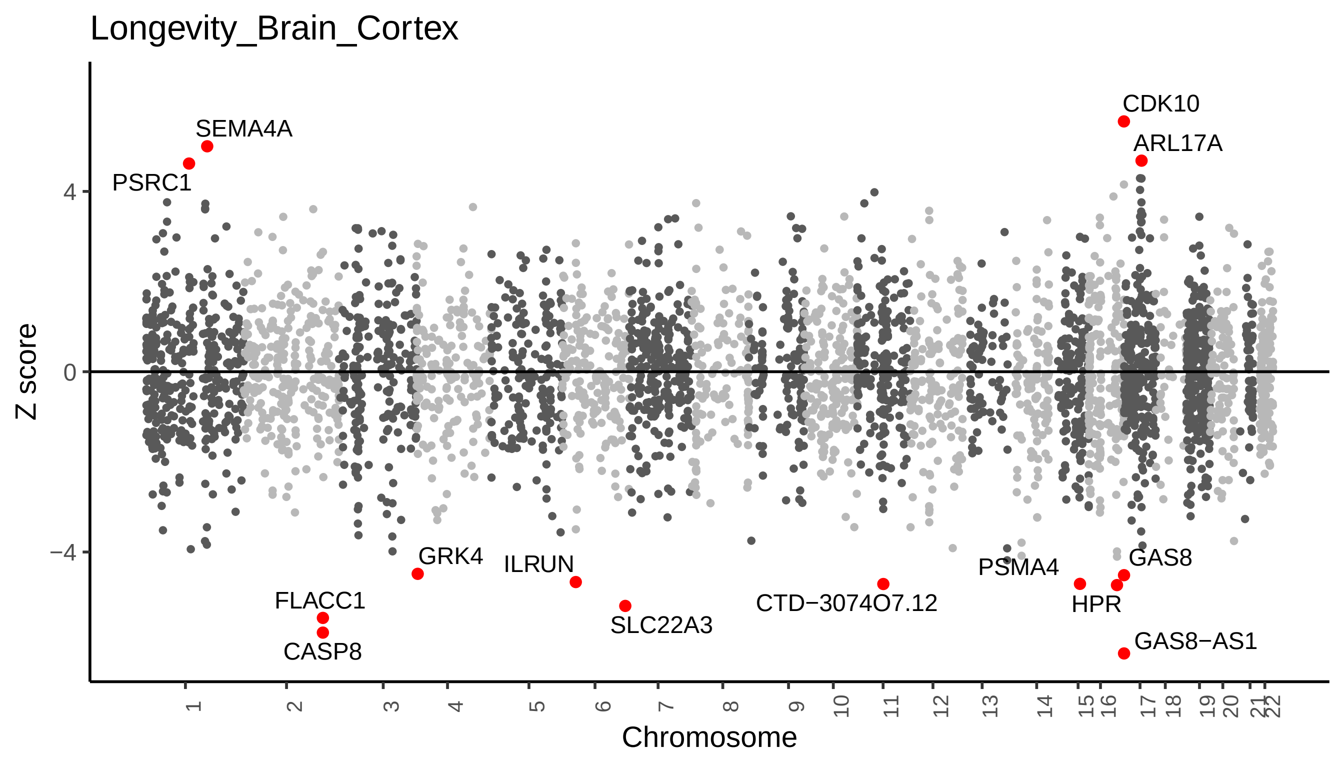

Longevity_Brain_Cortex
CDK10
SEMA4A
ARL17A
PSRC1
4
0
−4
Z score
GRK4
ILRUN
PSMA4
GAS8
FLACC1
CTD−3074O7.12
HPR
SLC22A3
CASP8
GAS8−AS1
Chromosome

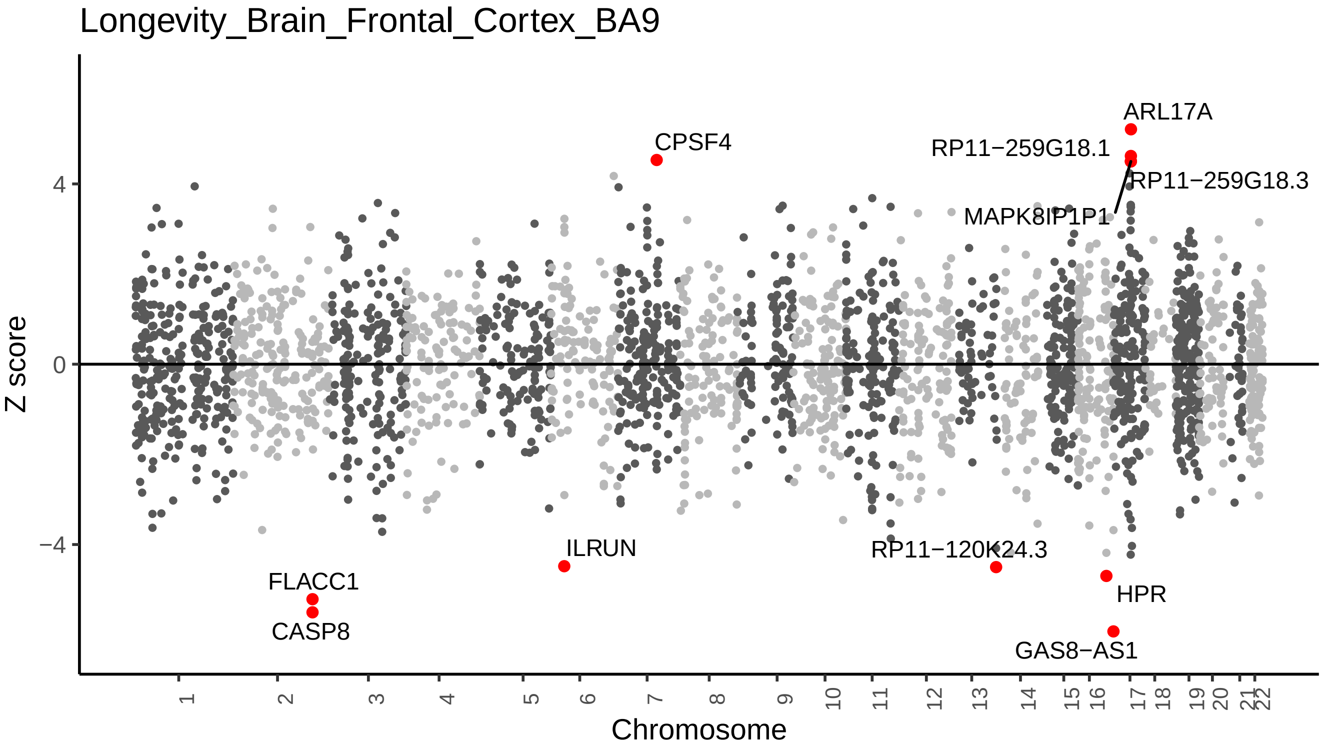

Longevity_Brain_Frontal_Cortex_BA9
Z score
4
0
−4
Chromosome
ARL17A
CPSF4
RP11−259G18.1
RP11−259G18.3
MAPK8IP1P1
ILRUN
RP11−120K24.3
FLACC1
CASP8
HPR
GAS8−AS1

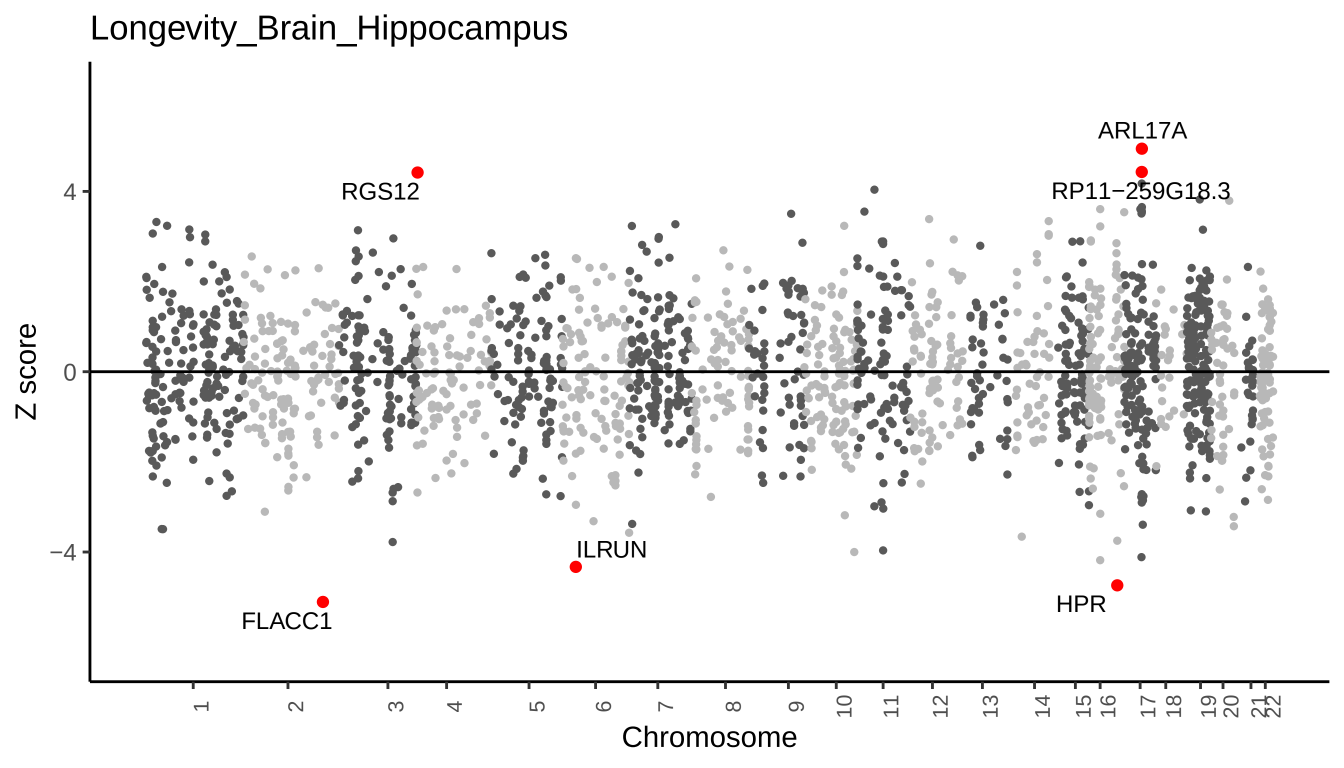

Longevity_Brain_Hippocampus
Z score
4
0
−4
RGS12
ARL17A
RP11−259G18.3
ILRUN
FLACC1
HPR
Chromosome

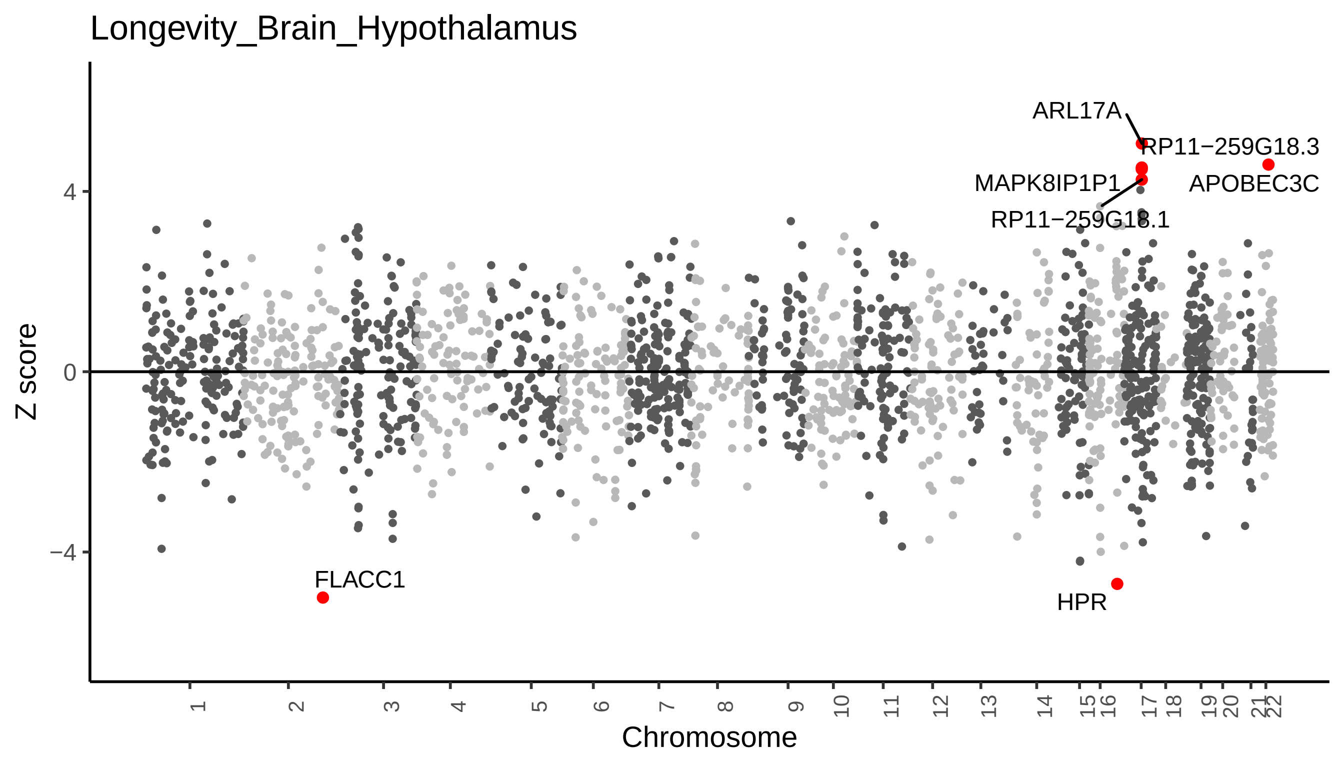

Longevity_Brain_Hypothalamus
Z score
4
0
−4
Chromosome
ARL17A
RP11−259G18.3
MAPK8IP1P1
APOBEC3C
RP11−259G18.1
FLACC1
HPR

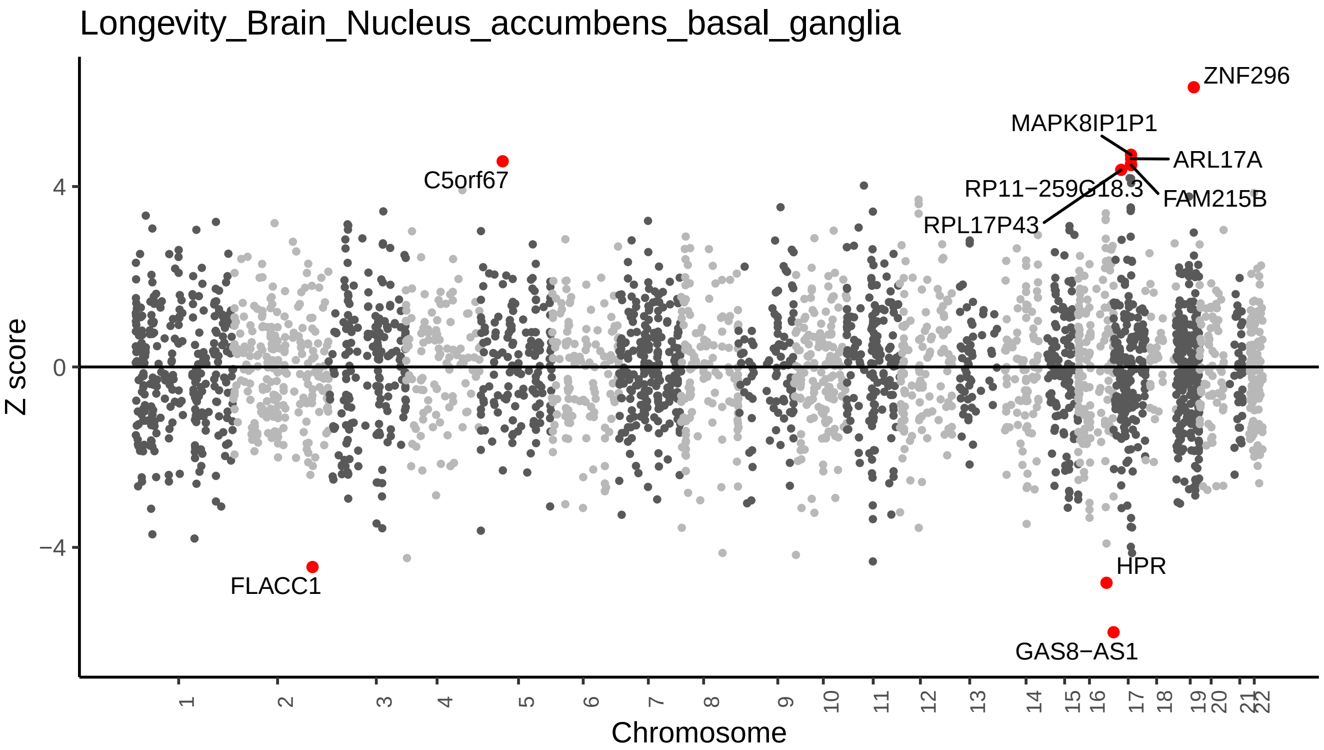
Longevity_Brain_Nucleus_accumbens_basal_ganglia
Z score
4
0
−4
ZNF296
MAPK8IP1P1
ARL17A
C5orf67
RP11−259G18.3
FAM215B
RPL17P43
FLACC1
HPR
GAS8−AS1
Chromosome

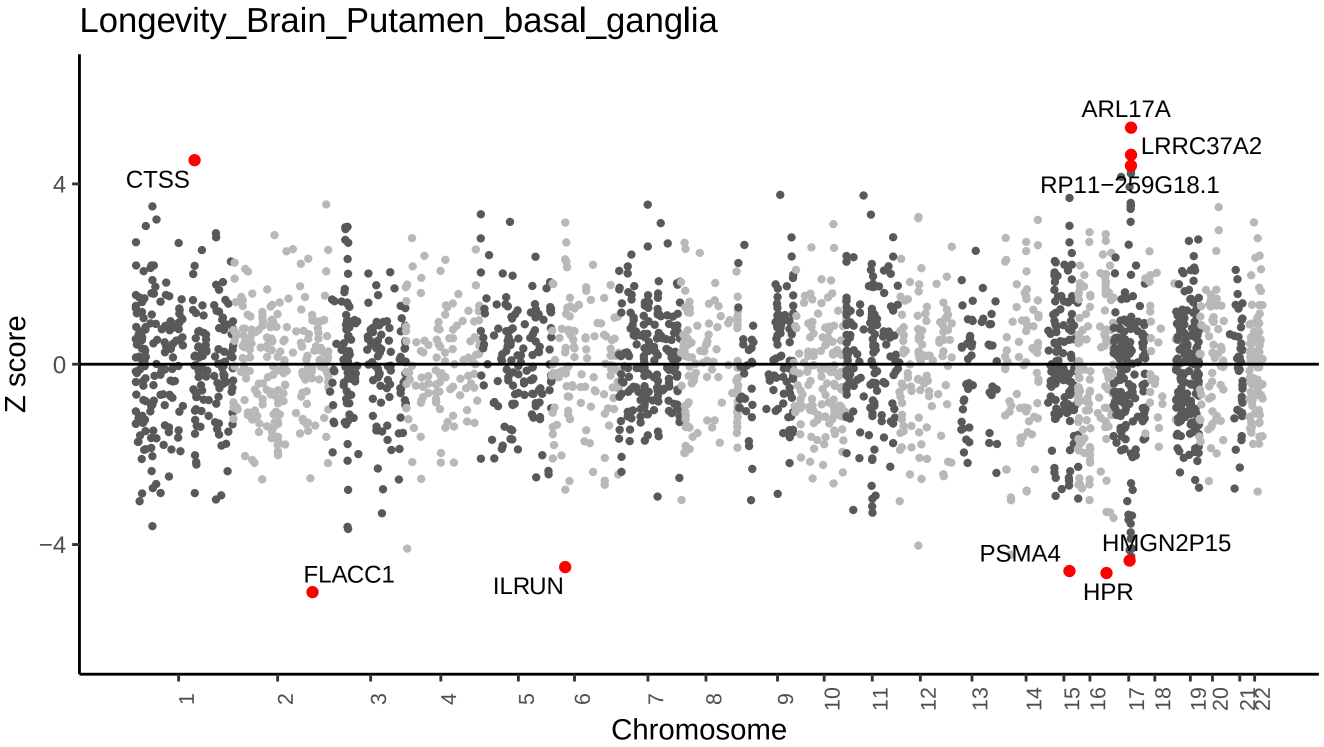

Longevity_Brain_Putamen_basal_ganglia
ARL17A
LRRC37A2
RP11−259G18.1
CTSS
FLACC1
ILRUN
PSMA4
HMGN2P15
HPR
4
0
−4
Z score
Chromosome

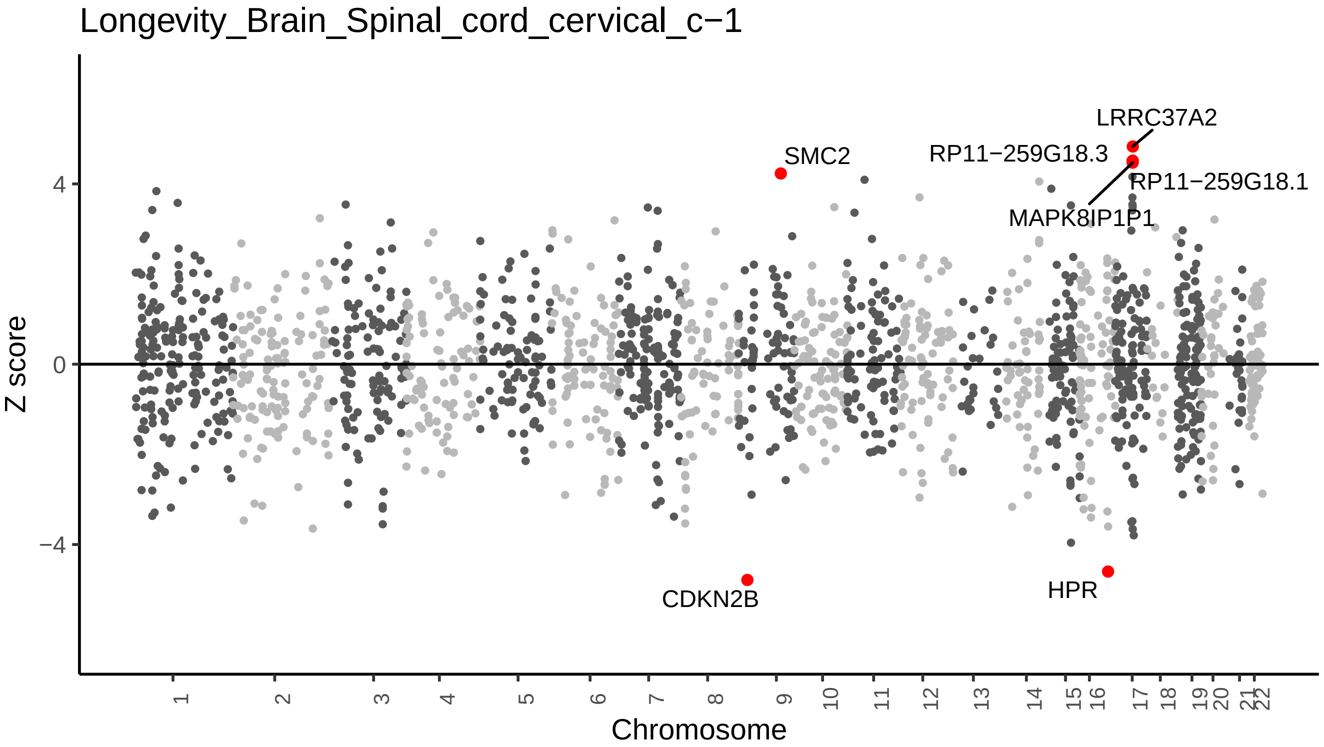

Longevity_Brain_Spinal_cord_cervical_c−1
Z score
4
0
−4
Chromosome
LRRC37A2
SMC2
RP11−259G18.3
RP11−259G18.1
MAPK8IP1P1
CDKN2B
HPR

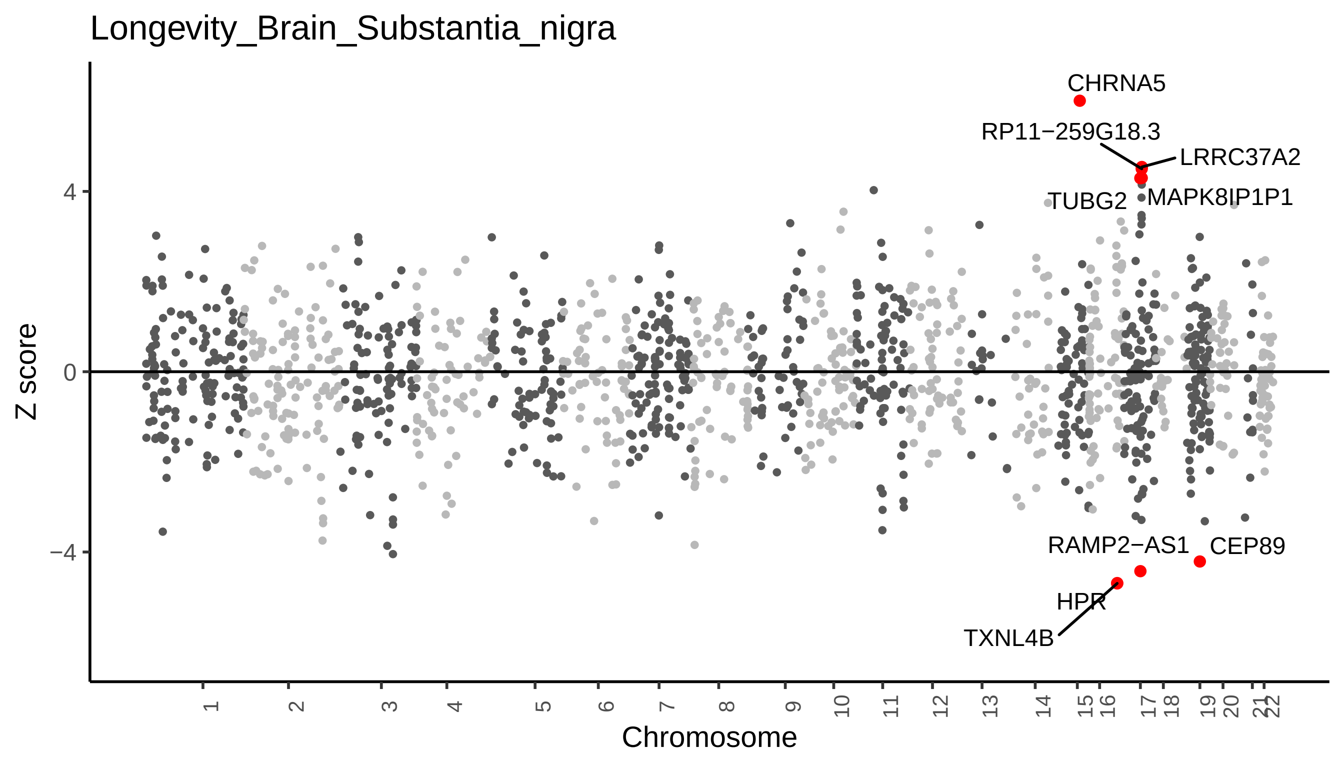

Longevity_Brain_Substantia_nigra
CHRNA5
RP11−259G18.3
LRRC37A2
TUBG2
MAPK8IP1P1
RAMP2−AS1
CEP89
HPR
TXNL4B
4
0
−4
Z score
Chromosome

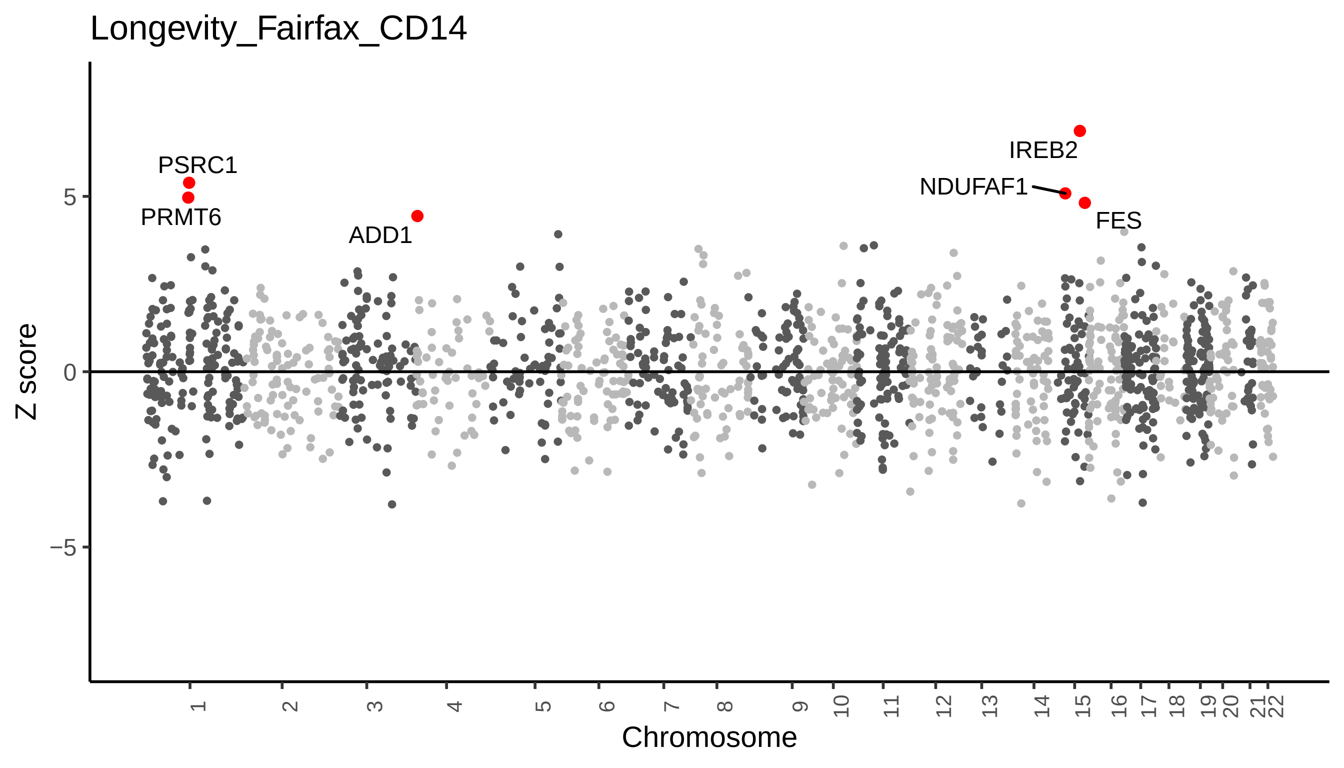

Longevity_Fairfax_CD14
Z score
5
0
−5
Chromosome
PSRC1
PRMT6
ADD1
IREB2
NDUFAF1
FES
