## Supplementary Table for "Genetic variation associated with human longevity and Alzheimer’s disease risk act through microglia and oligodendrocyte cross-talk"

**Supplementary Table 1. Gene-based analysis results (p ≤ 0.01) for ageing-associated genes using GWAS summary statistics from Timmers *et al.* (2020)^10^.**

| **GENE** | **CHR** | **START** | **STOP** | **NSNPS** | **NPARAM** | **Z** | **P** | **P_FDR** |
| --- | --- | --- | --- | --- | --- | --- | --- | --- |
| *PRMT6* | 1 | 107599301 | 107610258 | 9 | 3 | 4.4443 | 4.41E-06 | 3.62E-03 |
| *PHTF1* | 1 | 114239453 | 114302111 | 98 | 12 | 4.5211 | 3.08E-06 | 2.78E-03 |
| *RSBN1* | 1 | 114304454 | 114355098 | 67 | 14 | 4.2724 | 9.67E-06 | 6.47E-03 |
| *DCLRE1B* | 1 | 114447763 | 114456708 | 12 | 5 | 4.5479 | 2.71E-06 | 2.78E-03 |
| *IGF2BP2* | 3 | 185361527 | 185542844 | 258 | 28 | 4.2995 | 8.56E-06 | 6.19E-03 |
| *IRF4* | 6 | 391739 | 411447 | 67 | 15 | 4.3072 | 8.27E-06 | 6.19E-03 |
| *SKIV2L* | 6 | 31926857 | 31937532 | 34 | 12 | 4.2394 | 1.12E-05 | 7.05E-03 |
| *STK19* | 6 | 31938868 | 31950598 | 25 | 10 | 4.5255 | 3.01E-06 | 2.78E-03 |
| *FOXO3* | 6 | 108881038 | 109005977 | 160 | 19 | 4.4992 | 3.41E-06 | 2.93E-03 |
| *SLC22A3* | 6 | 160769300 | 160873613 | 217 | 19 | 5.4427 | 2.62E-08 | 3.65E-05 |
| *LPA* | 6 | 160952515 | 161085291 | 222 | 20 | 6.0449 | 7.48E-10 | 1.35E-06 |
| *ZKSCAN5* | 7 | 99102274 | 99132323 | 40 | 8 | 4.2372 | 1.13E-05 | 7.05E-03 |
| *AL359922.1* | 9 | 21802635 | 22032985 | 423 | 52 | 4.2906 | 8.91E-06 | 6.19E-03 |
| *CDKN2B* | 9 | 22002902 | 22009362 | 7 | 3 | 4.4039 | 5.32E-06 | 4.18E-03 |
| *TCF7L2* | 10 | 114710009 | 114927437 | 453 | 70 | 10.244 | 6.29E-25 | 2.84E-21 |
| *FGFR2* | 10 | 123237848 | 123357972 | 211 | 46 | 5.4456 | 2.58E-08 | 3.65E-05 |
| *IREB2* | 15 | 78729773 | 78793798 | 126 | 13 | 9.3585 | 4.04E-21 | 1.22E-17 |
| *HYKK* | 15 | 78799906 | 78829714 | 69 | 6 | 7.1569 | 4.12E-13 | 8.28E-10 |
| *CHRNA5* | 15 | 78857862 | 78887611 | 65 | 9 | 9.9362 | 1.45E-23 | 5.24E-20 |
| *CHRNA3* | 15 | 78885394 | 78913637 | 64 | 8 | 9.2953 | 7.34E-21 | 1.90E-17 |
| *CHRNB4* | 15 | 78916461 | 79020096 | 225 | 28 | 5.7408 | 4.71E-09 | 7.74E-06 |
| *FURIN* | 15 | 91411822 | 91426688 | 27 | 6 | 4.7823 | 8.66E-07 | 9.79E-04 |
| *FES* | 15 | 91426925 | 91439006 | 31 | 6 | 5.1119 | 1.59E-07 | 2.06E-04 |
| *TOX3* | 16 | 52471917 | 52581714 | 220 | 36 | 4.8045 | 7.76E-07 | 9.35E-04 |
| *GAS8* | 16 | 90086037 | 90111383 | 112 | 16 | 4.1695 | 1.53E-05 | 9.19E-03 |
| *ATP6V0A1* | 17 | 40610862 | 40674629 | 64 | 14 | 4.5341 | 2.89E-06 | 2.78E-03 |
| *NECTIN2* | 19 | 45349432 | 45392485 | 160 | 24 | 7.6575 | 9.48E-15 | 2.14E-11 |
| *TOMM40* | 19 | 45393826 | 45406946 | 51 | 11 | 13.712 | 4.31E-43 | 2.60E-39 |
| *APOE* | 19 | 45409011 | 45412650 | 6 | 3 | 14.882 | 2.16E-50 | 1.95E-46 |
| *APOC1* | 19 | 45417504 | 45422606 | 8 | 4 | 17.331 | 1.38E-67 | 2.49E-63 |
| Gene-based analysis was perfomed using MAGMA v1.09a (default parameters). No annotation window was added around the genes. The p-value was corrected for multiple testing using FDR method (a≤0.05).  Key: CHR, Chromosome; NSNPS, Number of Single Nucleotide Polymorphisms; NPARAM, Number of Parameters; Z, z-score statistic; P, p-value; P_FDR, p-value after adjusting for multiple testing using the False Discovery Rate method. | | | | | | | | |

**Supplementary Table 2. Gene-based analysis results (p ≤ 0.01) for genes associated with Alzheimer’s disease using GWAS summary statistics from Kunkle *et al.* (2019)^21^.**

| **GENE** | **CHR** | **START** | **STOP** | **NSNPS** | **NPARAM** | **Z** | **P** | **P_FDR** |
| --- | --- | --- | --- | --- | --- | --- | --- | --- |
| *PCSK9* | 1 | 55505221 | 55530525 | 122 | 26 | 3.9737 | 3.54E-05 | 8.76E-03 |
| *INAVA* | 1 | 200860176 | 200884863 | 75 | 17 | 4.0663 | 2.39E-05 | 6.27E-03 |
| *KIF21B* | 1 | 200938518 | 200992828 | 199 | 28 | 4.2541 | 1.05E-05 | 3.05E-03 |
| *CR1* | 1 | 207669492 | 207815110 | 333 | 29 | 5.4331 | 2.77E-08 | 1.47E-05 |
| *CR1L* | 1 | 207818458 | 207911761 | 458 | 34 | 4.2064 | 1.30E-05 | 3.60E-03 |
| *BIN1* | 2 | 127805603 | 127864931 | 275 | 30 | 6.2961 | 1.53E-10 | 9.45E-08 |
| *FSIP2* | 2 | 186603622 | 186698017 | 297 | 17 | 4.0013 | 3.15E-05 | 7.91E-03 |
| *INPP5D* | 2 | 233924677 | 234116549 | 722 | 62 | 4.3107 | 8.14E-06 | 2.52E-03 |
| *TMPRSS11B* | 4 | 69092371 | 69111438 | 29 | 4 | 3.9627 | 3.71E-05 | 9.06E-03 |
| *PFDN1* | 5 | 139624624 | 139682706 | 120 | 16 | 3.9373 | 4.12E-05 | 9.94E-03 |
| *HBEGF* | 5 | 139712428 | 139726216 | 38 | 11 | 4.9412 | 3.88E-07 | 1.72E-04 |
| *C6orf10* | 6 | 32256303 | 32339689 | 940 | 35 | 4.2473 | 1.08E-05 | 3.09E-05 |
| *HLA-DRB5* | 6 | 32485120 | 32498064 | 461 | 24 | 4.7174 | 1.19E-06 | 4.72E-04 |
| *HLA-DRB1* | 6 | 32546546 | 32557625 | 291 | 20 | 4.9941 | 2.96E-07 | 1.41E-04 |
| *HLA-DQB1* | 6 | 32627244 | 32636160 | 549 | 25 | 4.0191 | 2.92E-05 | 7.43E-03 |
| *TREM2* | 6 | 41126244 | 41130924 | 5 | 3 | 5.5509 | 1.42E-08 | 7.76E-06 |
| *CD2AP* | 6 | 47445525 | 47594999 | 434 | 24 | 4.6093 | 2.02E-06 | 6.95E-04 |
| *EPHA1* | 7 | 143087382 | 143105985 | 47 | 15 | 4.7118 | 1.23E-06 | 4.75E-04 |
| *PTK2B* | 8 | 27168999 | 27316908 | 548 | 40 | 4.9568 | 3.58E-07 | 1.66E-04 |
| *CHRNA2* | 8 | 27317278 | 27337400 | 101 | 22 | 4.2762 | 9.50E-06 | 2.85E-03 |
| *CLU* | 8 | 27454434 | 27472548 | 45 | 9 | 7.3792 | 7.96E-14 | 8.70E-11 |
| *CRY2* | 11 | 45868669 | 45904799 | 82 | 19 | 4.1231 | 1.87E-05 | 5.03E-03 |
| *MYBPC3* | 11 | 47352948 | 47374253 | 53 | 17 | 4.8274 | 6.92E-07 | 2.99E-04 |
| *SPI1* | 11 | 47376411 | 47400127 | 87 | 14 | 6.7216 | 8.98E-12 | 6.68E-09 |
| *SLC39A13* | 11 | 47428683 | 47438052 | 27 | 8 | 6.4158 | 7.01E-11 | 4.65E-08 |
| *PSMC3* | 11 | 47440320 | 47448024 | 23 | 7 | 5.9752 | 1.15E-09 | 6.88E-07 |
| *C1QTNF4* | 11 | 47611216 | 47616211 | 6 | 4 | 4.2632 | 1.01E-05 | 2.97E-03 |
| *AGBL2* | 11 | 47681143 | 47736941 | 143 | 22 | 4.5995 | 2.12E-06 | 7.15E-04 |
| *FNBP4* | 11 | 47738069 | 47788995 | 132 | 19 | 4.6206 | 1.91E-06 | 6.71E-04 |
| *NUP160* | 11 | 47799639 | 47870107 | 186 | 24 | 4.7723 | 9.11E-07 | 3.84E-04 |
| *MS4A3* | 11 | 59824060 | 59838601 | 51 | 9 | 5.1727 | 1.15E-07 | 5.79E-05 |
| *MS4A2* | 11 | 59855734 | 59865939 | 33 | 7 | 6.6981 | 1.06E-11 | 7.54E-09 |
| *MS4A6A* | 11 | 59939487 | 59952139 | 33 | 7 | 6.9355 | 2.02E-12 | 1.72E-09 |
| *MS4A4A* | 11 | 59953175 | 60085417 | 451 | 34 | 7.6247 | 1.22E-14 | 1.62E-11 |
| *MS4A4E* | 11 | 59968726 | 60010561 | 155 | 19 | 6.9226 | 2.22E-12 | 1.79E-09 |
| *PICALM* | 11 | 85668727 | 85780924 | 453 | 38 | 6.9346 | 2.04E-12 | 1.72E-09 |
| *SORL1* | 11 | 121322912 | 121504402 | 420 | 64 | 4.0453 | 2.61E-05 | 6.74E-03 |
| *SLC24A4* | 14 | 92788925 | 92967827 | 899 | 72 | 4.6246 | 1.88E-06 | 6.70E-04 |
| *KNOP1* | 16 | 19713256 | 19729557 | 44 | 7 | 4.2323 | 1.16E-05 | 3.25E-03 |
| *IQCK* | 16 | 19727778 | 19869789 | 344 | 21 | 4.7649 | 9.45E-07 | 3.90E-04 |
| *MTSS1L* | 16 | 70695107 | 70719969 | 103 | 22 | 4.6476 | 1.68E-06 | 6.24E-04 |
| *CNN2* | 19 | 1026580 | 1039067 | 82 | 16 | 4.5477 | 2.71E-06 | 9.00E-04 |
| *ABCA7* | 19 | 1040100 | 1065571 | 121 | 22 | 5.7776 | 3.79E-09 | 2.13E-06 |
| *ARHGAP45* | 19 | 1065922 | 1086627 | 90 | 19 | 4.0655 | 2.40E-05 | 6.27E-03 |
| *ISYNA1* | 19 | 18545198 | 18549111 | 9 | 2 | 4.1995 | 1.34E-05 | 3.65E-03 |
| *IGSF23* | 19 | 45116940 | 45140081 | 75 | 14 | 5.9345 | 1.47E-09 | 8.56E-07 |
| *PVR* | 19 | 45147098 | 45166850 | 48 | 11 | 6.6252 | 1.73E-11 | 1.19E-08 |
| *CEACAM19* | 19 | 45165545 | 45187631 | 36 | 12 | 7.1541 | 4.21E-13 | 4.12E-10 |
| *CEACAM16* | 19 | 45202421 | 45213986 | 35 | 11 | 4.3919 | 5.62E-06 | 1.83E-03 |
| *BCL3* | 19 | 45250962 | 45263301 | 30 | 10 | 12.132 | 3.56E-34 | 1.32E-30 |
| *CBLC* | 19 | 45281126 | 45303909 | 63 | 12 | 9.7609 | 8.29E-23 | 1.71E-19 |
| *BCAM* | 19 | 45312316 | 45324678 | 45 | 9 | 8.8004 | 6.82E-19 | 1.27E-15 |
| *NECTIN2* | 19 | 45349432 | 45392485 | 188 | 28 | 23.065 | 5.24E-118 | 9.73E-114 |
| *TOMM40* | 19 | 45393826 | 45406946 | 50 | 11 | 22.38 | 3.07E-111 | 2.86E-107 |
| *APOE* | 19 | 45409011 | 45412650 | 6 | 3 | 19.036 | 4.29E-81 | 1.99E-77 |
| *APOC1* | 19 | 45417504 | 45422606 | 10 | 5 | 22.061 | 3.77E-108 | 2.33E-104 |
| *APOC4* | 19 | 45445495 | 45448753 | 9 | 4 | 10.157 | 1.55E-24 | 3.59E-21 |
| *APOC4-APOC2* | 19 | 45445495 | 45452822 | 27 | 5 | 7.5012 | 3.16E-14 | 3.67E-11 |
| *APOC2* | 19 | 45449239 | 45452822 | 16 | 3 | 6.3673 | 9.62E-11 | 6.16E-08 |
| *CLPTM1* | 19 | 45457842 | 45496599 | 116 | 16 | 8.5278 | 7.46E-18 | 1.26E-14 |
| *RELB* | 19 | 45504688 | 45541456 | 83 | 24 | 10.24 | 6.56E-25 | 1.74E-21 |
| *CLASRP* | 19 | 45542298 | 45574214 | 64 | 11 | 7.2985 | 1.45E-13 | 1.50E-10 |
| *GEMIN7* | 19 | 45582453 | 45594782 | 72 | 5 | 4.6276 | 1.85E-06 | 6.70E-04 |
| *MARK4* | 19 | 45582546 | 45808541 | 810 | 61 | 12.023 | 1.34E-33 | 4.15E-30 |
| *PPP1R37* | 19 | 45594654 | 45651335 | 198 | 19 | 7.6296 | 1.18E-14 | 1.62E-11 |
| *NKPD1* | 19 | 45653008 | 45661995 | 22 | 9 | 7.6996 | 6.83E-15 | 1.06E-11 |
| *TRAPPC6A* | 19 | 45666186 | 45681495 | 45 | 10 | 7.016 | 1.14E-12 | 1.06E-09 |
| *AC005779.2* | 19 | 45683080 | 45705702 | 96 | 19 | 7.5404 | 2.34E-14 | 2.90E-11 |
| *EXOC3L2* | 19 | 45715879 | 45748689 | 148 | 16 | 6.8647 | 3.33E-12 | 2.58E-09 |
| *FBXO46* | 19 | 46213887 | 46234162 | 31 | 14 | 5.2774 | 6.55E-08 | 3.38E-05 |
| *BHMG1* | 19 | 46236509 | 46267792 | 59 | 12 | 5.0252 | 2.51E-07 | 1.23E-04 |
| *SIX5* | 19 | 46268043 | 46272484 | 6 | 3 | 4.377 | 6.02E-06 | 1.89E-03 |
| *DMPK* | 19 | 46272975 | 46285810 | 21 | 7 | 4.3805 | 5.92E-06 | 1.89E-03 |
| *AC011530.1* | 19 | 46282695 | 46289231 | 12 | 5 | 4.661 | 1.57E-06 | 5.97E-04 |
| *DMWD* | 19 | 46286205 | 46296060 | 25 | 10 | 4.9494 | 3.72E-07 | 1.69E-04 |
| *RSPH6A* | 19 | 46298968 | 46318577 | 70 | 14 | 4.2853 | 9.13E-06 | 2.78E-03 |
| *CD33* | 19 | 51728320 | 51747115 | 34 | 12 | 4.7427 | 1.05E-06 | 4.26E-04 |
| Gene-based analysis was perfomed using MAGMA v1.09a (default parameters). No annotation window was added around the genes. The p-value was corrected for multiple testing using FDR method (a≤0.05).  Key: CHR, Chromosome; NSNPS, Number of Single Nucleotide Polymorphisms; NPARAM, Number of Parameters; Z, z-score statistic; P, p-value; P_FDR, p-value after adjusting for multiple testing using the False Discovery Rate method. | | | | | | | | |

**Supplementary Table 3. Human hippocampus bulk RNA-seq microglial module genes and their gene-based analysis p-value for Alzheimer’s disease.**

| **Mouse Symbol** | **Human Symbol** | **Human Chromosome** | **Start Location** | **End Location** | **AD Gene**  **P-value** |
| --- | --- | --- | --- | --- | --- |
| *Ms4a4a, Ms4a4b, Ms4a4c, Ms4a4d* | *MS4A4A* | 11 | 59953175 | 60085417 | 1.22E-14 |
| *Ms4a6c, Ms4a6b, Ms4a6d* | *MS4A6A* | 11 | 59939487 | 59952139 | 2.02E-12 |
| *Spi1* | *SPI1* | 11 | 47376411 | 47400127 | 8.98E-12 |
| *Apoc2, Gm44805* | *APOC2* | 19 | 45449239 | 45452822 | 9.62E-11 |
| *Trem2* | *TREM2* | 6 | 41126244 | 41130924 | 1.42E-08 |
| *H2-Eb2* | *HLA-DRB1* | 6 | 32546546 | 32557625 | 2.96E-07 |
| *-* | *CD33* | 19 | 51728320 | 51747115 | 1.05E-06 |
| *H2-Eb2* | *HLA-DRB5* | 6 | 32485120 | 32498064 | 1.19E-06 |
| *Inpp5d* | *INPP5D* | 2 | 233924677 | 234116549 | 8.14E-06 |
| *Arhgap45* | *ARHGAP45* | 19 | 1065922 | 1086627 | 2.40E-05 |
| *H2-Ab1* | *HLA-DQB1* | 6 | 32627244 | 32636160 | 2.92E-05 |
| *Laptm5* | *LAPTM5* | 1 | 31205316 | 31230667 | 6.64E-05 |
| *-* | *HLA-DQA1* | 6 | 32595956 | 32614839 | 3.96E-04 |
| *H2-Ea* | *HLA-DRA* | 6 | 32407619 | 32412823 | 9.98E-04 |
| *-* | *NOP2* | 12 | 6666029 | 6677857 | 1.32E-03 |
| *Atp8b4* | *ATP8B4* | 15 | 50150435 | 50475014 | 1.36E-03 |
| *Gpsm3* | *GPSM3* | 6 | 32158543 | 32163300 | 3.83E-03 |
| *Gal3st4* | *GAL3ST4* | 7 | 99756867 | 99766373 | 4.60E-03 |
| *Cmtm7* | *CMTM7* | 3 | 32433163 | 32524559 | 5.34E-03 |
| *Pced1b* | *PCED1B* | 12 | 47473386 | 47630445 | 5.61E-03 |
| *Itgam, Gm49368* | *ITGAM* | 16 | 31271288 | 31344213 | 5.77E-03 |
| *Dok3* | *DOK3* | 5 | 176928908 | 176938275 | 6.00E-03 |
| *Tmc8* | *TMC8* | 17 | 76126851 | 76139049 | 7.03E-03 |
| *Marco* | *MARCO* | 2 | 119699742 | 119752236 | 8.90E-03 |
| *Cox7a1* | *COX7A1* | 19 | 36641824 | 36643771 | 9.05E-03 |
| *Lilra5, Lilra6, Pira2, Pirb,*  *Gm14548, Gm15922,*  *Gm15922* | *LILRB4* | 19 | 55155340 | 55181810 | 9.11E-03 |
| p-value is not multiple testing corrected.  Key: AD, Alzheimer’s disease.  Full network given in Supplementary Table 16. | | | | | |

**Supplementary Table 4. Human hippocampus bulk RNA-seq oligodendrocytic module genes and their gene-based analysis p-value for Alzheimer’s disease.**

| **Mouse Symbol** | **Human Symbol** | **Human Chromosome** | **Start Location** | **End Location** | **AD Gene**  **P-value** |
| --- | --- | --- | --- | --- | --- |
| *Clasrp* | *CLASRP* | 19 | 45542298 | 45574214 | 1.45E-13 |
| *Hnrnpa2b1* | *HNRNPA2B1* | 7 | 26212677 | 26241149 | 1.24E-04 |
| *Tex22* | *TEX22* | 14 | 105864916 | 105916443 | 3.00E-04 |
| *Zkscan1* | *ZKSCAN1* | 7 | 99613195 | 99639312 | 5.63E-04 |
| *Shc4* | *SHC4* | 15 | 49115932 | 49255641 | 9.78E-04 |
| *-* | *BBIP1* | 10 | 112658488 | 112679032 | 1.45E-03 |
| *Folh1* | *FOLH1* | 11 | 49168187 | 49230222 | 1.51E-03 |
| *Slc45a3* | *SLC45A3* | 1 | 205626979 | 205649587 | 1.77E-03 |
| *Cdr2l* | *CDR2L* | 17 | 72983727 | 73001895 | 1.80E-03 |
| *Fam76b* | *FAM76B* | 11 | 95502106 | 95523573 | 1.91E-03 |
| *Zfpl1* | *ZFPL1* | 11 | 64851682 | 64855872 | 1.99E-03 |
| *Pdcd4* | *PDCD4* | 10 | 112631553 | 112659764 | 2.08E-03 |
| *Tmem37* | *TMEM37* | 2 | 120187477 | 120196096 | 2.40E-03 |
| *Slc20a2* | *SLC20A2* | 8 | 42273993 | 42397069 | 2.50E-03 |
| *Pvrig* | *PVRIG* | 7 | 99815864 | 99819113 | 3.58E-03 |
| *Cenpc1* | *CENPC* | 4 | 68334466 | 68411324 | 3.78E-03 |
| *Tmed7* | *TMED7* | 5 | 114949205 | 114968689 | 3.98E-03 |
| *Slc26a9* | *SLC26A9* | 1 | 205882176 | 205912588 | 4.40E-03 |
| *Stag3* | *STAG3* | 7 | 99775186 | 99819111 | 4.59E-03 |
| *Acp7* | *ACP7* | 19 | 39574553 | 39602133 | 4.85E-03 |
| *Snx1* | *SNX1* | 15 | 64386322 | 64438289 | 4.85E-03 |
| *Tmem42* | *TMEM42* | 3 | 44903361 | 44907162 | 5.31E-03 |
| *Tardbp* | *TARDBP* | 1 | 11072401 | 11086477 | 5.59E-03 |
| *Unc5cl* | *UNC5CL* | 6 | 40994650 | 41006956 | 6.39E-03 |
| *Slf2* | *SLF2* | 10 | 102672326 | 102724893 | 6.64E-03 |
| *Mettl14* | *METTL14* | 4 | 119606523 | 119636588 | 7.43E-03 |
| *Fam222a* | *FAM222A* | 12 | 110152033 | 110208312 | 7.52E-03 |
| *Slc4a9* | *SLC4A9* | 5 | 139739787 | 139754728 | 8.65E-03 |
| *Uba6* | *UBA6* | 4 | 68478370 | 68566897 | 9.57E-03 |
| p-value is not multiple testing corrected.  Key: AD, Alzheimer’s disease.  Full network given in Supplementary Table 17. | | | | | |

**Supplementary Table 5. Mouse hippocampus scRNA-seq activated response microglial, ARM, module genes and their gene-based analysis p-value for Alzheimer’s disease.**

| **Mouse Symbol** | **Human Symbol** | **Human Chromosome** | **Start Location** | **End Location** | **AD Gene**  **P-value** |
| --- | --- | --- | --- | --- | --- |
| Apoe | *APOE* | 19 | 45409011 | 45412650 | 4.29E-81 |
| Relb | *RELB* | 19 | 45504688 | 45541456 | 6.56E-25 |
| Ms4a6c | *MS4A6A* | 11 | 59939487 | 59952139 | 2.02E-12 |
| Pvr | *PVR* | 19 | 45147098 | 45166850 | 1.73E-11 |
| Ptk2b | *PTK2B* | 8 | 27168999 | 27316908 | 3.58E-07 |
| H2-Ab1 | *HLA-DQB1* | 6 | 32627244 | 32636160 | 2.92E-05 |
| Plekha1 | *PLEKHA1* | 10 | 124134212 | 124202118 | 6.88E-04 |
| Ydjc | *YDJC* | 22 | 21982378 | 21984353 | 9.94E-04 |
| Pirb | *LILRA5* | 19 | 54818353 | 54824409 | 1.30E-03 |
| Trim37 | *TRIM37* | 17 | 57059999 | 57184282 | 1.79E-03 |
| Nrp1 | *NRP1* | 10 | 33466420 | 33625190 | 2.65E-03 |
| Stbd1 | *STBD1* | 4 | 77227179 | 77232752 | 3.02E-03 |
| Alkbh2 | *ALKBH2* | 12 | 109525993 | 109531436 | 4.03E-03 |
| Ank | *ANKH* | 5 | 14704909 | 14871894 | 6.30E-03 |
| Tnip2 | *TNIP2* | 4 | 2743375 | 2758103 | 8.29E-03 |
| Wdr55 | *WDR55* | 5 | 140044261 | 140053709 | 8.80E-03 |
| Pirb | *LILRB4* | 19 | 55155340 | 55181810 | 9.11E-03 |
| p-value is not multiple testing corrected.  Key: AD, Alzheimer’s disease.  Full network given in Supplementary Table 12. | | | | | |

**Supplementary Table 6. Mouse hippocampus scRNA-seq phagolysosomal module genes and their gene-based analysis p-value for Alzheimer’s disease.**

| **Mouse Symbol** | **Human Symbol** | **Human Chromosome** | **Start Location** | **End Location** | **AD Gene**  **P-value** |
| --- | --- | --- | --- | --- | --- |
| Tomm40 | *TOMM40* | 19 | 45393826 | 45406946 | 3.07E-111 |
| Clptm1 | *CLPTM1* | 19 | 45457842 | 45496599 | 7.46E-18 |
| Ms4a6d | *MS4A6A* | 11 | 59939487 | 59952139 | 2.02E-12 |
| Spi1 | *SPI1* | 11 | 47376411 | 47400127 | 8.98E-12 |
| Psmc3 | *PSMC3* | 11 | 47440320 | 47448024 | 1.15E-09 |
| Trem2 | *TREM2* | 6 | 41126244 | 41130924 | 1.42E-08 |
| Fbxo46 | *FBXO46* | 19 | 46213887 | 46234162 | 6.55E-08 |
| Hbegf | *HBEGF* | 5 | 139712428 | 139726216 | 3.88E-07 |
| Gemin7 | *GEMIN7* | 19 | 45582453 | 45594782 | 1.85E-06 |
| Isyna1 | *ISYNA1* | 19 | 18545198 | 18549111 | 1.34E-05 |
| Pfdn1 | *PFDN1* | 5 | 139624624 | 139682706 | 4.12E-05 |
| Ssbp4 | *SSBP4* | 19 | 18529674 | 18545372 | 4.91E-05 |
| Mtch2 | *MTCH2* | 11 | 47638867 | 47664175 | 1.08E-04 |
| Cnpy4 | *CNPY4* | 7 | 99717236 | 99723134 | 1.71E-04 |
| Fcf1 | *FCF1* | 14 | 75179847 | 75205323 | 9.68E-04 |
| Ndufaf6 | *NDUFAF6* | 8 | 95907995 | 96128683 | 1.08E-03 |
| Lilra5 | *LILRA5* | 19 | 54818353 | 54824409 | 1.30E-03 |
| Dlst | *DLST* | 14 | 75348594 | 75370448 | 1.39E-03 |
| Actb | *ACTB* | 7 | 5566778 | 5603415 | 1.64E-03 |
| Csnk2b | *CSNK2B* | 6 | 31633013 | 31638120 | 1.76E-03 |
| Arpc1a | *ARPC1A* | 7 | 98923533 | 98963885 | 1.76E-03 |
| Zyx | *ZYX* | 7 | 143078173 | 143088204 | 2.00E-03 |
| Prkra | *PRKRA* | 2 | 179296141 | 179316239 | 2.10E-03 |
| Tmem37 | *TMEM37* | 2 | 120187477 | 120196096 | 2.40E-03 |
| Zfp655 | *ZNF655* | 7 | 99156029 | 99174076 | 2.80E-03 |
| Pkp4 | *PKP4* | 2 | 159313476 | 159539391 | 2.97E-03 |
| Psmc6 | *PSMC6* | 14 | 53173890 | 53195305 | 3.74E-03 |
| Fam220a | *FAM220A* | 7 | 6369040 | 6388612 | 4.78E-03 |
| Snx1 | *SNX1* | 15 | 64386322 | 64438289 | 4.85E-03 |
| Tmem42 | *TMEM42* | 3 | 44903361 | 44907162 | 5.31E-03 |
| Bnip3l | *BNIP3L* | 8 | 26240414 | 26363152 | 5.42E-03 |
| Ddah2 | *DDAH2* | 6 | 31694815 | 31698394 | 5.98E-03 |
| Mrpl43 | *MRPL43* | 10 | 102729215 | 102747272 | 6.03E-03 |
| Eif1b | *EIF1B* | 3 | 40351175 | 40353915 | 6.89E-03 |
| 2900026A02Rik | *KIAA1671* | 22 | 25348697 | 25593415 | 7.14E-03 |
| Zmat2 | *ZMAT2* | 5 | 140078265 | 140086261 | 7.57E-03 |
| Polr2e | *POLR2E* | 19 | 1086578 | 1095379 | 7.88E-03 |
| Ech1 | *ECH1* | 19 | 39306062 | 39322645 | 8.23E-03 |
| Ik | *IK* | 5 | 140026643 | 140042064 | 8.81E-03 |
| Lilra5 | *LILRB4* | 19 | 55155340 | 55181810 | 9.11E-03 |
| Oser1 | *OSER1* | 20 | 42825136 | 42839431 | 9.42E-03 |
| Etf1 | *ETF1* | 5 | 137841784 | 137878989 | 9.58E-03 |
| Skp1a | *SKP1* | 5 | 133484626 | 133512729 | 9.81E-03 |
| Mrpl58 | *MRPL58* | 17 | 73008765 | 73017356 | 9.92E-03 |
| Grn | *GRN* | 17 | 42422454 | 42430470 | 1.00E-02 |
| p-value is not multiple testing corrected.  Key: AD, Alzheimer’s disease.  Full network given in Supplementary Table 15. | | | | | |

**Supplementary Table 7. Mouse hippocampus scRNA-seq homeostatic microglial subcluster 2, HM2, module genes and their gene-based analysis p-value for longevity.**

| **Mouse Symbol** | **Human Symbol** | **Human Chromosome** | **Start Location** | **End Location** | **Longevity Gene P-value** |
| --- | --- | --- | --- | --- | --- |
| Fes | *FES* | 15 | 91426925 | 91439006 | 1.59E-07 |
| Zkscan5 | *ZKSCAN5* | 7 | 99102274 | 99132323 | 1.13E-05 |
| Ptcd1 | *PTCD1* | 7 | 99014362 | 99063786 | 3.28E-05 |
| Ptcd1 | *ATP5MF-PTCD1* | 7 | 99017372 | 99063820 | 3.59E-05 |
| Jam3 | *JAM3* | 11 | 133938820 | 134021896 | 1.77E-04 |
| Casp8 | *CASP8* | 2 | 202098166 | 202152434 | 2.80E-04 |
| Mafk | *MAFK* | 7 | 1570350 | 1582679 | 5.51E-04 |
| Mcrs1 | *MCRS1* | 12 | 49950327 | 49961936 | 9.35E-04 |
| Golph3l | *GOLPH3L* | 1 | 150618701 | 150669620 | 1.47E-03 |
| Rbm4 | *RBM4* | 11 | 66406088 | 66435845 | 1.56E-03 |
| Ppp2r3c | *PPP2R3C* | 14 | 35554673 | 35591723 | 1.62E-03 |
| Bcap29 | *BCAP29* | 7 | 107220422 | 107269615 | 1.66E-03 |
| Rit1 | *RIT1* | 1 | 155867599 | 155881195 | 1.86E-03 |
| Srp54b | *SRP54* | 14 | 35451163 | 35498773 | 1.92E-03 |
| 1700066M21Rik | *C2orf69* | 2 | 200775979 | 200820658 | 1.96E-03 |
| Cln8 | *CLN8* | 8 | 1703944 | 1749877 | 2.05E-03 |
| Plpp3 | *PLPP3* | 1 | 56960419 | 57110974 | 2.58E-03 |
| Maip1 | *MAIP1* | 2 | 200820040 | 200873263 | 3.01E-03 |
| Bud31 | *BUD31* | 7 | 99006264 | 99017239 | 3.22E-03 |
| Stat3 | *STAT3* | 17 | 40465342 | 40540586 | 3.25E-03 |
| Scamp5 | *SCAMP5* | 15 | 75249560 | 75313837 | 3.51E-03 |
| Klc2 | *KLC2* | 11 | 66024765 | 66035331 | 3.70E-03 |
| Ptpn1 | *PTPN1* | 20 | 49126858 | 49201778 | 4.24E-03 |
| Zkscan14 | *ZNF394* | 7 | 99084142 | 99097947 | 5.02E-03 |
| Zfp664 | *ZNF664* | 12 | 124456392 | 124499986 | 5.63E-03 |
| Exoc3 | *EXOC3* | 5 | 443273 | 472052 | 5.66E-03 |
| Dusp6 | *DUSP6* | 12 | 89741009 | 89747048 | 5.76E-03 |
| Slc44a2 | *SLC44A2* | 19 | 10713133 | 10755235 | 6.45E-03 |
| Xpc | *XPC* | 3 | 14186647 | 14220283 | 6.56E-03 |
| Tap2 | *TAP2* | 6 | 32789610 | 32806557 | 6.57E-03 |
| Ctsf | *CTSF* | 11 | 66330934 | 66336312 | 6.69E-03 |
| Csnk1a1 | *CSNK1A1* | 5 | 148871760 | 148931115 | 6.84E-03 |
| Gstz1 | *GSTZ1* | 14 | 77787227 | 77797940 | 7.66E-03 |
| Fads1 | *FADS1* | 11 | 61567097 | 61596790 | 7.85E-03 |
| Usp38 | *USP38* | 4 | 144106070 | 144144983 | 8.63E-03 |
| F11r | *F11R* | 1 | 160965001 | 160991138 | 9.06E-03 |
| Bsn | *BSN* | 3 | 49591922 | 49708978 | 9.08E-03 |
| Abhd16a | *ABHD16A* | 6 | 31654726 | 31671221 | 9.35E-03 |
| Zscan29 | *ZSCAN29* | 15 | 43650370 | 43663223 | 9.44E-03 |
| p-value is not multiple testing corrected.  Full network given in Supplementary Table 14. | | | | | |
